# Exome sequencing identifies rare damaging variants in *ATP8B4* and *ABCA1* as novel risk factors for Alzheimer’s Disease

**DOI:** 10.1101/2020.07.22.20159251

**Authors:** Henne Holstege, Marc Hulsman, Camille Charbonnier, Benjamin Grenier-Boley, Olivier Quenez, Detelina Grozeva, Jeroen G.J. van Rooij, Rebecca Sims, Shahzad Ahmad, Najaf Amin, Penny J. Norsworthy, Oriol Dols-Icardo, Holger Hummerich, Amit Kawalia, Alzheimer’s Disease Neuroimaging Initiative (ADNI) database, Philippe Amouyel, Gary W. Beecham, Claudine Berr, Joshua C. Bis, Anne Boland, Paola Bossù, Femke Bouwman, Jose Bras, Dominique Campion, J. Nicholas Cochran, Antonio Daniele, Jean-François Dartigues, Stéphanie Debette, Jean-François Deleuze, Nicola Denning, Anita L DeStefano, Lindsay A. Farrer, Maria Victoria Fernandez, Nick C. Fox, Daniela Galimberti, Emmanuelle Genin, Hans Gille, Yann Le Guen, Rita Guerreiro, Jonathan L. Haines, Clive Holmes, M. Arfan Ikram, M. Kamran Ikram, Iris E. Jansen, Robert Kraaij, Marc Lathrop, Afina W. Lemstra, Alberto Lleó, Lauren Luckcuck, Marcel M. A. M. Mannens, Rachel Marshall, Eden R Martin, Carlo Masullo, Richard Mayeux, Patrizia Mecocci, Alun Meggy, Merel O. Mol, Kevin Morgan, Richard M. Myers, Benedetta Nacmias, Adam C Naj, Valerio Napolioni, Florence Pasquier, Pau Pastor, Margaret A. Pericak-Vance, Rachel Raybould, Richard Redon, Marcel J.T. Reinders, Anne-Claire Richard, Steffi G Riedel-Heller, Fernando Rivadeneira, Stéphane Rousseau, Natalie S. Ryan, Salha Saad, Pascual Sanchez-Juan, Gerard D. Schellenberg, Philip Scheltens, Jonathan M. Schott, Davide Seripa, Sudha Seshadri, Daoud Sie, Erik Sistermans, Sandro Sorbi, Resie van Spaendonk, Gianfranco Spalletta, Niccólo Tesi, Betty Tijms, André G Uitterlinden, Sven J. van der Lee, Pieter Jelle de Visser, Michael Wagner, David Wallon, Li-San Wang, Aline Zarea, Jordi Clarimon, John C. van Swieten, Michael D. Greicius, Jennifer S. Yokoyama, Carlos Cruchaga, John Hardy, Alfredo Ramirez, Simon Mead, Wiesje M. van der Flier, Cornelia M van Duijn, Julie Williams, Gaël Nicolas, Céline Bellenguez, Jean-Charles Lambert

**Author notes:** To whom correspondence should be addressed –Henne Holstege –Marc Hulsman –Gael Nicolas –Jean-Charles Lambert. Authors contributed equally to this work (Henne Holstege, Marc Hulsman, Camille Charbonnier, Gaël Nicolas, Céline Bellenguez, Jean-Charles Lambert). ADNI consortium; see supplemental authors.

## Abstract

The genetic component of Alzheimer’s disease (AD) has been mainly assessed using Genome Wide Association Studies (GWAS), which do not capture the risk contributed by rare variants. Here, we compared the gene-based burden of rare damaging variants in exome sequencing data from 32,558 individuals —16,036 AD cases and 16,522 controls— in a two-stage analysis. Next to known genes *TREM2, SORL1* and *ABCA7*, we observed a significant association of rare, predicted damaging variants in *ATP8B4* and *ABCA1* with AD risk, and a suggestive signal in *ADAM10*. Next to these genes, the rare variant burden in *RIN3, CLU, ZCWPW1* and *ACE* highlighted these genes as potential driver genes in AD-GWAS loci. Rare damaging variants in these genes, and in particular loss-of-function variants, have a large effect on AD-risk, and they are enriched in early onset AD cases. The newly identified AD-associated genes provide additional evidence for a major role for APP-processing, Aβ-aggregation, lipid metabolism and microglial function in AD.

---

AD is the leading cause of dementia and its impact will grow with increasing life expectancy^1^. Beyond autosomal dominant early onset AD (<1% of all AD cases, onset ≤65 years), the common complex form of AD has an estimated heritability of ∼70%^2^. Using genome wide association studies (GWAS), 75 mostly common genetic risk factors/loci have been associated with AD-risk in populations with European ancestry, but individually these common variants have low effect-sizes^3^. Using DNA-sequencing strategies, rare (allele frequency <1%) damaging missense or loss-of-function (LOF) variants in the *TREM2, SORL1* and *ABCA7* genes were identified to also contribute to the heritability of AD, with substantially higher effect-sizes than individual GWAS-hits^4-8^.

In order to detect additional genes for which rare variants associate with AD-risk, it is necessary to compare genetic sequencing data from thousands of AD cases and controls. In a large collaborative effort, we harmonized sequencing data from studies from Europe and the United States (**Table S1**) and applied a multi-stage gene burden analysis (**Figure 1A**). We observed site-specific technical biases, since data were generated at multiple centers, using heterogeneous methods. To account for these batch effects, we designed and applied comprehensive quality control (QC) procedures (**Online methods, Tables S2-S3**).

**Fig 1.**
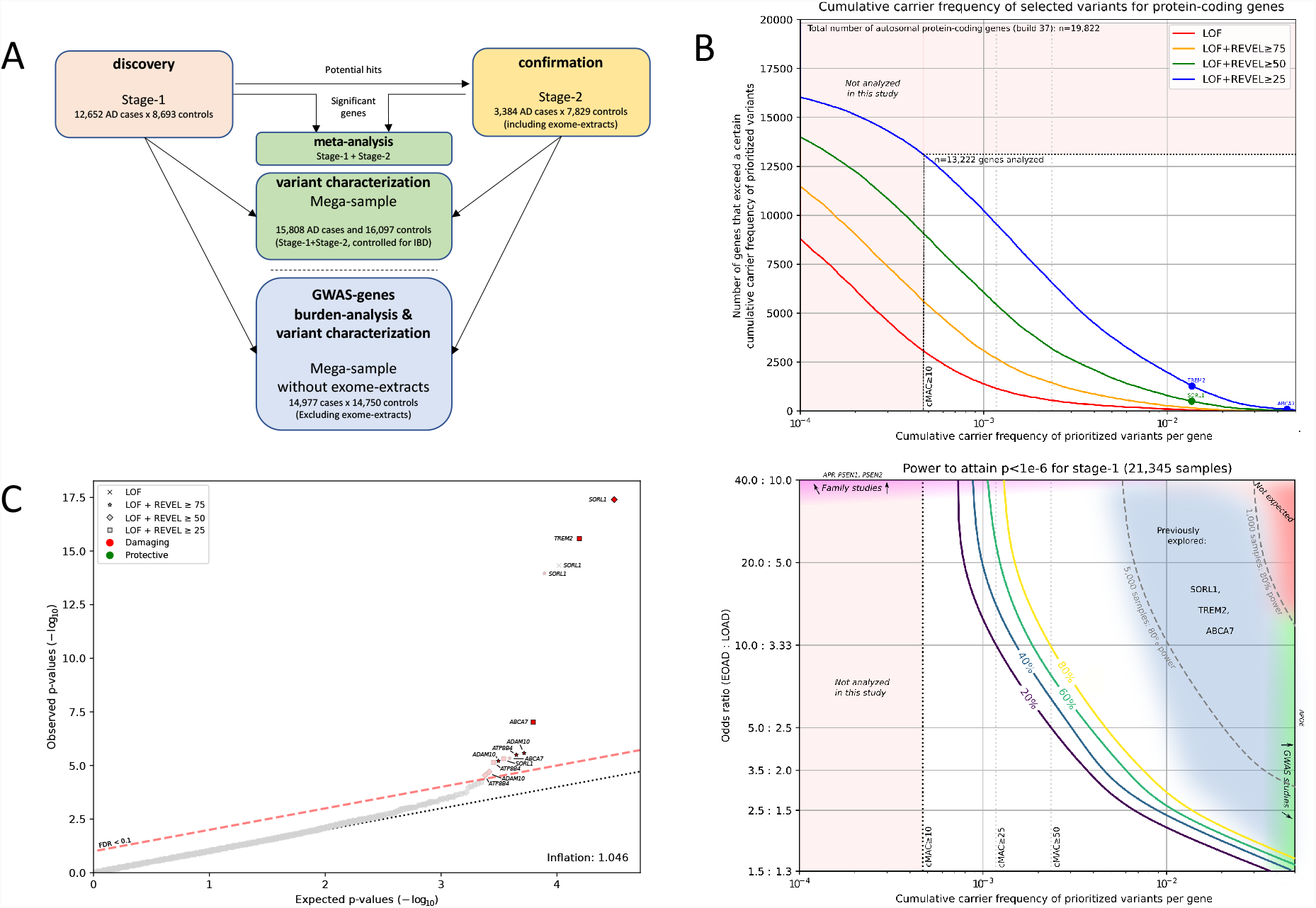
Study set-up and power. **A:** Schematic of study set-up; The AD-association of genes identified in Stage-1 was confirmed in Stage-2, and combined significance was determined in a meta-analysis. To accurately estimate variant effect-sizes for variant-categories/age at onset bins, even those with very low variant counts for which the normal-distribution approximation fails, we analyzed variant characteristics in a mega-sample instead of through meta-analysis. This mega-sample was also used for the GWAS gene burden analysis (without exome-extracts). **B**. Top: the number of genes (Y-axis) that have at least a certain cumulative carrier frequency of prioritized variants (X-axis). Variants were prioritized according to different deleteriousness thresholds. White box: only genes with a cMAC≥10 (cumulative minor allele count of at least 10 alleles across the sample) in the Stage-1 sample of 12,652 cases and 8,693 controls were considered to have a high enough carrier frequency allowing burden analysis. The previously identified *SORL1, TREM2*, and *ABCA7* genes are indicated, revealing that carriers of damaging variants in these genes are relatively common, which has aided their identification in the past. **B**. Bottom: Power analysis for Stage-1, to attain a p<1e^-6^, on the same scale as the top figure B. For comparison purposes, we also plot 80% power thresholds for 5,000 and 1,000 samples (subsampled from Stage-1). Approximate regions are indicated for variants identified with GWAS (red) or family studies (purple), as well as the region in which variant-burdens in *SORL1, TREM2* and *ABCA7* were identified by previous sequencing studies^4-8^ (dark-blue). Common variants with very high effectsizes (red) are not expected to exist. Genes with cMAC<10 were not analyzed (light-red). Power calculations show that, by aggregating more cases and controls, one might be able to identify burdens of rare variants with either (i) a large effect but with an extremely low frequency of carriers or (ii) with a modest average effect but a higher number of carriers. **C**. P-value Q-Q plot of Stage-1 discovery-analysis. Gene-names were indicated in grey when the deleteriousness threshold was not the most significant burden test in that gene.

After sample QC, we first compared gene-based rare-variant burdens between 12,652 AD cases and 8,693 controls (Stage-1 analysis, **Table 1A**). We detected 7,543,193 variants after sample- and variant-QC (**Table 1B**) and annotated LOF variants with LOFTEE and missense variants with the REVEL score, and selected variants with MAF<1%. We defined four deleteriousness thresholds by incrementally including variants with lower levels of predicted deleteriousness: respectively LOF (n=57,543), LOF+REVEL≥75 (n=111,755), LOF+REVEL≥50 (n=211,665), and LOF+REVEL≥25 (n=409,733). Of the 19,822 autosomal protein coding genes, we analyzed the 13,222 genes that had a cumulative minor allele count (cMAC) ≥10 for the lowest deleterious threshold LOF+REVEL≥25 (**see Methods**); 9,168 genes for the LOF+REVEL≥50 threshold; 5,694 for the LOF+REVEL≥75 threshold and 3,120 genes for the LOF-only threshold (**Figure 1B**). For these different deleteriousness thresholds, this analysis has an estimated power of 41%, 22%, 11% and 4%, respectively to attain a signal with p<1e-6, assuming that the differential variant burden for a gene is associated with an odds ratio of 10.0 in EOAD and 3.33 in LOAD (**Table S4**). Therefore, this analysis only has the power to uncover genes for which the differential gene-burden is associated with a large effect size or large numbers of damaging variant carriers (**Figure 1B**). In total, 31,204 tests were performed across 13,222 genes in Stage-1 (single genes were tested with up to four thresholds). Statistical inflation of test results was negligible (*λ*=1.046, **Figure 1C**). Of all burden tests performed, 13 tests, covering 6 genes indicated a suggestive differential variant burden between AD cases and controls (FDR<0.1): *SORL1, TREM2, ABCA7, ATP8B4, ADAM10*, and *ABCA1* (**Table 2A**).

**Table 1:** Quality Control Steps. **A. Sample QC:** Samples were primarily excluded due to non-European ancestry or close family relations, for details see **Table S3A**. Exome-extract samples (between parenthesis) only contain reads that cover the 10 genes discovered in Stage-1. In Stage-2, samples were removed that were duplicated w.r.t. Stage-1. In the mega-analysis, a merged sample QC removed all family relations to the third degree between Stage-1 and Stage-2 (i.e. the size of the mega-sample does not equal the sum of the Stage-1 and Stage-2 samples). **B. Variant QC:** Stage-1 consists of all variants in the union of the exome capture kits. The targeted Stage-2 and Mega analysis regards only the 10 genes identified in Stage-1. Variant QC from the non-targeted Stage-2 and mega-analysis can be found in **Table S3B**. For each gene, we considered in our variant selection 4 different selection thresholds. MAF: minor allele frequency. See supplement for in-depth QC methods.

| Table 1A: Sample Quality Control |  |  |  |  |
| --- | --- | --- | --- | --- |
| Sample |  | nr. of samples |  |  |
| Samples processed |  | Stage-1 | Stage-2 | Mega |
| Samples included in study |  | 25,982 | 26,379 | 52,361 |
| Samples retained after QC (Table S3A) |  | 21,345 | 11,213 | 31,905 |
| Sample totals (of which exome-extracts of targeted genes) |  |  |  |  |
| EOAD |  | 4,060 | 1,627 (446) | 5,643 (446) |
| LOAD |  | 8,592 | 1,757 (385) | 10,165 (385) |
| Controls |  | 8,693 | 7,829 (1,347) | 16,097 (1,347) |

| Table 1B: Variant Quality Control |  |  |  |  |
| --- | --- | --- | --- | --- |
| Variant QC |  | Targeted genes |  |  |
|  |  | Stage-1 | Stage-2 | Mega |
| Variants called (bi-allelic) |  | 12,938,556 | 7,803 | 14,531 |
| Variants retained after QC (Table S3B-C) |  | 7,543,193 | 5,072 | 8,963 |
| Variant selection |  |  |  |  |
| 1. In protein coding autosomal genes (Gencode V19/V29) |  | 6,883,630 | 5,072 | 8,963 |
| Missense variants | 2. Missense variants | 1,486,559 | 894 | 1,873 |
|  | 3. REVEL > 25 | 540,934 | 591 | 1,263 |
|  | 4. MAF < 1% / dosage > 0.5 | 530,072 | 567 | 1,228 |
|  | 5. Missingness (< 20% + no differential missingness) | 353,913 | 428 | 943 |
| LOF variants | 2. Loss-of-function variants (nonsense, frameshift, splice acceptor/donor) | 144,429 | 106 | 255 |
|  | 3. Loftee HC + VEP high impact | 109,550 | 97 | 236 |
|  | 4. MAF < 1% / dosage > 0.5 | 108,016 | 96 | 234 |
|  | 5. Missingness (< 20% + no differential missingness) | 57,543 | 64 | 168 |
| Variant Categories | REVEL 25-50 | 198,068 | 168 | 360 |
|  | REVEL 75-100 | 99,910 | 140 | 320 |
|  | REVEL 50-75 | 54,212 | 119 | 258 |
|  | LOF | 57,543 | 64 | 168 |
| Variant Thresholds | LOF+REVEL≥25 | 409,733 | 491 | 1,106 |
|  | LOF+REVEL≥50 | 211,665 | 323 | 746 |
|  | LOF+REVEL≥75 | 111,755 | 183 | 426 |
|  | LOF | 57,543 | 64 | 168 |

**Table 2.** Stage-1, Stage-2 Association statistics. **A. Stage-1, Stage-2 and Meta-analysis**. Burden tests that were significant in Stage-1 after multiple testing correction using the Benjamini-Hochberg False Discovery Rate (FDR) (<0.1) over 31,204 tests/variant categories. ^#^In Stage-2, we considered only the direction of the AD-association observed in Stage-1. The meta-analysis indicates the combined significance from Stages 1 and 2 (data was combined using the fixed-effect inverse variance method); multiple testing correction for the meta-analysis was performed across all 31,204 tests using the Holm-Bonferroni correction (<0.05). Bold black text: significant p-values; **B. GWAS-targeted analysis in mega-dataset without exome extracts**. Genes in all loci were prioritized as described in the Online Methods, (**Table S5**). *These genes also included the *SORL1, TREM2, ABCA7, ADAM10*, and *ABCA1* genes, which were also identified in the rare variant burden analysis (shown in A) and therefore not shown here. These genes are listed in the context of this analysis in **Table S6**. For both tables the p values were determined using ordinal logistic regression, and a case/control OR was computed for reference. Grey text: result burden test MAF<0.1% unchanged compared to burden test MAF<1%.

| Table 2A. Burden tests Stage-1, Stage-2 and Meta-analysis. |  |  |  |  |  |  |  |  |  |  |  |  |
| --- | --- | --- | --- | --- | --- | --- | --- | --- | --- | --- | --- | --- |
| gene | Variant damagingness threshold | Stage-1 |  |  |  | Stage-2 |  |  | meta |  |  |  |
|  |  | P value | FDR | #variants / #carriers | case / control | P value<sup>#</sup> | #variants / #carriers | case / control OR (95% CI) | P value | h-bonf | case / control OR (95% CI) | pvalue heterog. |
| SORL1 | LOF+REVEL≥25 | 4.8E-06 | **0.017** | 242 / 917 | 1.3 (1.1-1.5) | **1.3E-06** | 122 / 478 | 1.5 (1.2-1.9) | 1.5E-10 | **4.7E-06** | 1.4 (1.2-1.5) | 1.6E-01 |
|  | LOF+REVEL≥50 | 4.0E-18 | **<<0.0001** | 167 / 290 | 2.6 (2.0-3.2) | **1.4E-09** | 79 / 137 | 2.4 (1.7-3.5) | 8.1E-26 | **2.5E-21** | 2.5 (2.1-3.1) | 9.8E-01 |
|  | LOF+REVEL≥75 | 1.1E-14 | **<<0.0001** | 96 / 164 | 3.3 (2.4-4.6) | **5.2E-10** | 45 / 82 | 3.9 (2.3-6.6) | 1.1E-22 | **3.4E-18** | 3.5 (2.7-4.6) | 4.3E-01 |
|  | LOF | 4.7E-15 | **<<0.0001** | 37 / 48 | 15.6 (3.7-37.3) | **1.6E-06** | 16 / 20 | 16.3 (3.8-35.0) | 3.3E-18 | **1.0E-13** | 16.0 (9.5-27.0) | 9.4E-01 |
| TREM2 | LOF+REVEL≥25 | 2.6E-16 | **<<0.0001** | 17 / 291 | 3.6 (2.9-4.6) | **1.6E-07** | 12 / 155 | 2.4 (1.6-3.4) | 5.2E-22 | **1.6E-17** | 3.2 (2.6-3.9) | 6.5E-01 |
| ABCA7 | LOF+REVEL≥25 | 9.5E-08 | **0.001** | 265 / 959 | 1.4 (1.2-1.6) | **9.8E-08** | 170 / 502 | 1.6 (1.3-2.0) | 4.1E-13 | **1.3E-08** | 1.4 (1.3-1.6) | 6.5E-02 |
|  | LOF+REVEL≥75 | 4.6E-06 | **0.017** | 93 / 297 | 1.6 (1.3-2.1) | **4.8E-04** | 54 / 167 | 1.8 (1.3-2.6) | 7.3E-09 | **2.3E-04** | 1.7 (1.4-2.1) | 9.1E-01 |
| ATP8B4 | LOF+REVEL≥25 | 7.2E-06 | **0.02** | 72 / 575 | 1.5 (1.3-1.8) | **3.3E-03** | 40 / 286 | 1.4 (1.0-1.8) | 9.6E-09 | **3.0E-04** | 1.5 (1.3-1.7) | 9.7E-01 |
|  | LOF+REVEL≥50 | 2.8E-05 | **0.068** | 61 / 521 | 1.5 (1.3-1.9) | **1.6E-02** | 34 / 265 | 1.3 (1.0-1.7) | 2.8E-06 | **8.7E-02** | 1.5 (1.3-1.7) | 6.6E-01 |
|  | LOF+REVEL≥75 | 3.2E-06 | **0.014** | 38 / 490 | 1.7 (1.4-2.0) | **2.4E-02** | 22 / 243 | 1.3 (1.0-1.8) | 5.7E-07 | **1.8E-02** | 1.5 (1.3-1.8) | 4.2E-01 |
| ABCA1 | LOF+REVEL≥75 | 6.1E-06 | **0.019** | 93 / 280 | 1.7 (1.3-2.2) | **6.6E-03** | 48 / 159 | 1.6 (1.1-2.3) | 2.6E-07 | **8.0E-03** | 1.7 (1.4-2.1) | 6.3E-01 |
| ADAM10 | LOF+REVEL≥50 | 2.0E-05 | **0.051** | 15 / 17 | 3.2 (1.3-8.1) | **4.0E-02** | 4 / 4 | 8.1 (0.6-42.6) | 2.8E-05 | 8.7E-01 | 3.6 (1.5-8.5) | 5.5E-01 |
|  | LOF+REVEL≥75 | 2.7E-06 | **0.014** | 11 / 12 | 7.5 (1.4-46.8) | 1.5E-01 | 3 / 3 | 5.6 (0.3-41.8) | 4.4E-04 | 1.0E+00 | 7.1 (2.6-19.3) | 1.1E-01 |

| Table 2B. GWAS-targeted analysis on mega-dataset without exome-extracts |  |  |  |  |  |  |  |  |  |  |
| --- | --- | --- | --- | --- | --- | --- | --- | --- | --- | --- |
|  |  | Burden test (variant MAF <1%) |  |  |  |  | Burden test (variant MAF <0.1%) |  |  |  |
| Locus<br>*sentinal GWAS SNP* | gene | Variant<br>damagingness<br>threshold | P value | FDR | #variant /<br>#carriers | case / control<br>OR (95% CI) | pvalue | #variant /<br>#carriers | fraction<br>very rare | case / contro<br>OR (95% CI) |
| *SORL1, TREM2, ABCA7 (see Table 2A and S6) |  |  |  |  |  |  |  |  |  |  |
| SLC24A4/RIN3<br>rs7401792<br>rs12590654 | RIN3 | LOF+REVEL≥25 | 1.6E-05 | 0.0003 | 44 / 622 | 1.4 (1.2-1.6) | 3.4E-02 | 42 / 129 | 21% | 1.4 (1.0-2.1) |
|  |  | LOF+REVEL≥50 | 1.0E-05 | 0.0002 | 23 / 583 | 1.4 (1.2-1.7) | 1.5E-02 | 21 / 89 | 15% | 1.8 (1.2-2.8) |
| *ADAM10, ABCA1 (see Table 2A and S6) |  |  |  |  |  |  |  |  |  |  |
| PTK2B/CLU<br>rs73223431<br>rs11787077 | CLU | LOF+REVEL≥25 | 5.0E-04 | 0.005 | 24 / 26 | 3.6 (1.6-8.3) | 5.0E-04 | 24 / 26 | 100% | 3.6 (1.6-8.3) |
|  |  | LOF+REVEL≥50 | 1.1E-03 | 0.001 | 14 / 15 | 5.4 (1.6-28.6) | 1.1E-03 | 14 / 15 | 100% | 5.3 (1.6-28.6) |
|  |  | LOF+REVEL≥75 | 5.0E-04 | 0.005 | 12 / 12 | 9.9 (1.6-44.0) | 5.0E-04 | 12 / 12 | 100% | 9.8 (1.6-44.0) |
|  |  | LOF | 2.6E-03 | 0.02 | 10 / 10 | 7.3 (1.9-27.2) | 2.6E-03 | 10 / 10 | 100% | 7.3 (1.9-27.2) |
| SPDYE3<br>rs7384878 | ZCWPW1 | LOF+REVEL≥25 | 6.1E-03 | 0.042 | 22 / 77 | 1.8 (1.2-2.9) | 5.0E-03 | 21 / 76 | 99% | 1.8 (1.2-2.9) |
|  |  | LOF+REVEL≥50 | 3.1E-03 | 0.022 | 16 / 70 | 1.9 (1.2-3.1) | 3.1E-03 | 16 / 70 | 100% | 1.9 (1.2-3.1) |
|  |  | LOF+REVEL≥75 | 1.1E-03 | 0.001 | 11 / 15 | 5.0 (1.9-13.5) | 1.1E-03 | 11 / 15 | 100% | 5.0 (1.9-13.5) |
|  |  | LOF | 7.8E-04 | 0.008 | 11 / 15 | 5.0 (1.9-13.5) | 7.8E-04 | 11 / 15 | 100% | 5.0 (1.9-13.5) |
| ACE<br>rs4277405 | ACE | LOF+REVEL≥75 | 9.0E-04 | 0.008 | 38 / 99 | 2.0 (1.3-2.9) | 9.3E-04 | 38 / 99 | 100% | 2.0 (1.3-2.9) |

To confirm these signals, we applied an analysis model consistent with Stage-1 to an independent Stage-2 dataset, which after QC, comprised 3,384 cases and 7,829 controls (**Table 1A**). The effect was tested in the direction observed in Stage-1 (one-sided test). All genes selected in Stage-1 reached p<0.05 (**Table 2A, Stage-2**). Stage-2 effect-sizes of these genes correlated with those observed in Stage-1 (Pearson’s r on log-odds: 0.91). We then meta-analyzed Stage-1 + Stage-2 across the 13 tests using a fixed-effect inverse variance method and corrected for the 31,204 tests performed in Stage-1 (Holm-Bonferroni) (**Table 2A**). This confirmed the AD-association of rare damaging variants in the *SORL1, TREM2, ABCA7, ATP8B4* and *ABCA1* genes. The association signal of the *ADAM10* gene was not exome-wide significant, presumably because prioritized variants in this gene are extremely few and rare, such that the signal can be confirmed only in larger datasets.

Strikingly, most of these genes also map to GWAS loci (*SORL1, TREM2, ABCA7, ABCA1* and *ADAM10)*. This led us to perform a focused analysis on GWAS loci, aiming to identify potential driver genes. To maximize statistical power, we merged the full exomes from the Stage-1 and Stage-2 samples into one mega-sample (**Table 1**). We interrogated genes that were previously prioritized to drive the AD association in the 75 loci identified in the most recent GWAS^3^ (**Table S5, Online Methods**). In 67 genes, we observed sufficient prioritized variants (cMAC ≥10) to test the burden signal in at least one deleteriousness category (a total of 187 tests). In addition to the genes mentioned above, our analysis indicated a suggestive signal of increased AD risk in *RIN3, CLU, ZCWPW1*, and *ACE* (FDR<0.05) (**Table 2B, Table S6**); these signals will have to be confirmed in a larger dataset. Nevertheless, the AD associations in these genes persisted when focusing on the burden of only the very rare variants (MAF<0.1%), suggesting that the rare variant burden is not in linkage with, and thus independent from the GWAS sentinel variant.

Together, the newly associated genes provide additional evidence for a central role for *APP*-processing, lipid metabolism, Aβ-aggregation and neuroinflammatory processes in AD pathophysiology. Like *ABCA7, ATP8B4* is a phospholipid transporter. Rare variants in this gene have been associated with the risk of developing systemic sclerosis, an autoimmune disease^9^. In the brain, *ATP8B4* is predominantly expressed in microglia. Interestingly, GWAS indicated a potential association of *ATP8B4* with AD^3^, mainly through the rare missense variant that was most recurrent in our study (G395S). Of note, the odds ratio point-estimate for *ATP8B4* LOF variants was close to 1, allowing for the possibility that the missense variants that drive the *ATP8B4* association do not depend on a loss-of-function effect. ABCA1 is also a phospholipid transporter; it lipidates APOE^10^ and poor ABCA1-dependent lipidation of APOE-containing lipoprotein particles increases Aβ-deposition and fibrillogenesis^11^. In line with this, the rare N1800H loss-of-function variant in *ABCA1* was previously associated with low plasma levels of ApoE and evidence suggested an association with increased risk of AD and cerebrovascular disease^12^. The α-secretase ADAM10 plays a major role in non-amyloidogenic APP metabolism^13^. Evidence for the AD-association of rare variants in *ADAM10* has remained suggestive until now: two rare missense variants in *ADAM10* were reported before to incompletely segregate with LOAD in a few families^14^ (these variants did not associate with AD in our study, **Table SG6)** and a nonsense variant in the *ADAM10* gene was found to segregate with AD but in a small pedigree^15^. **Error! Bookmark not defined**.*RIN3* has been associated with endosomal dysfunction and APP trafficking/metabolism^16,17^. *CLU* (also known as *APOJ*) has been found to affect Aβ-aggregation and clearance^18^ and *ACE* is suggested to have a role in Aβ-degradation^19^. Thus far, the role of the histone methylation reader *ZCWPW1* remains unclear.

To better comprehend how these genes associate with AD, we analyzed the characteristics of rare damaging variants that contributed to the burden using the mega-sample (**Figure 2, Table 3**) For damaging variants in most genes, we observed increased carrier frequencies in younger cases and larger effect sizes were associated with an earlier age at onset (p=0.0001) (**Table S7**). Yet the variants also contributed to an increased risk of late-onset AD (**Figures 2A-B, Table 3**). The largest effect-sizes were measured for LOF variants in *SORL1, ADAM10, CLU* and *ZCWPW1*, and carriers of such variants had the lowest median age at onset (**Table 3**), implying a key role for these genes in AD etiology. Moderate variant-effect-sizes were observed for LOF variants in *TREM2, ABCA1* and *RIN3*, while the smallest variant-effects were observed in *ABCA7, ATP8B4* and *ACE (***Figure 3, Table 3**).

**Table 3.**
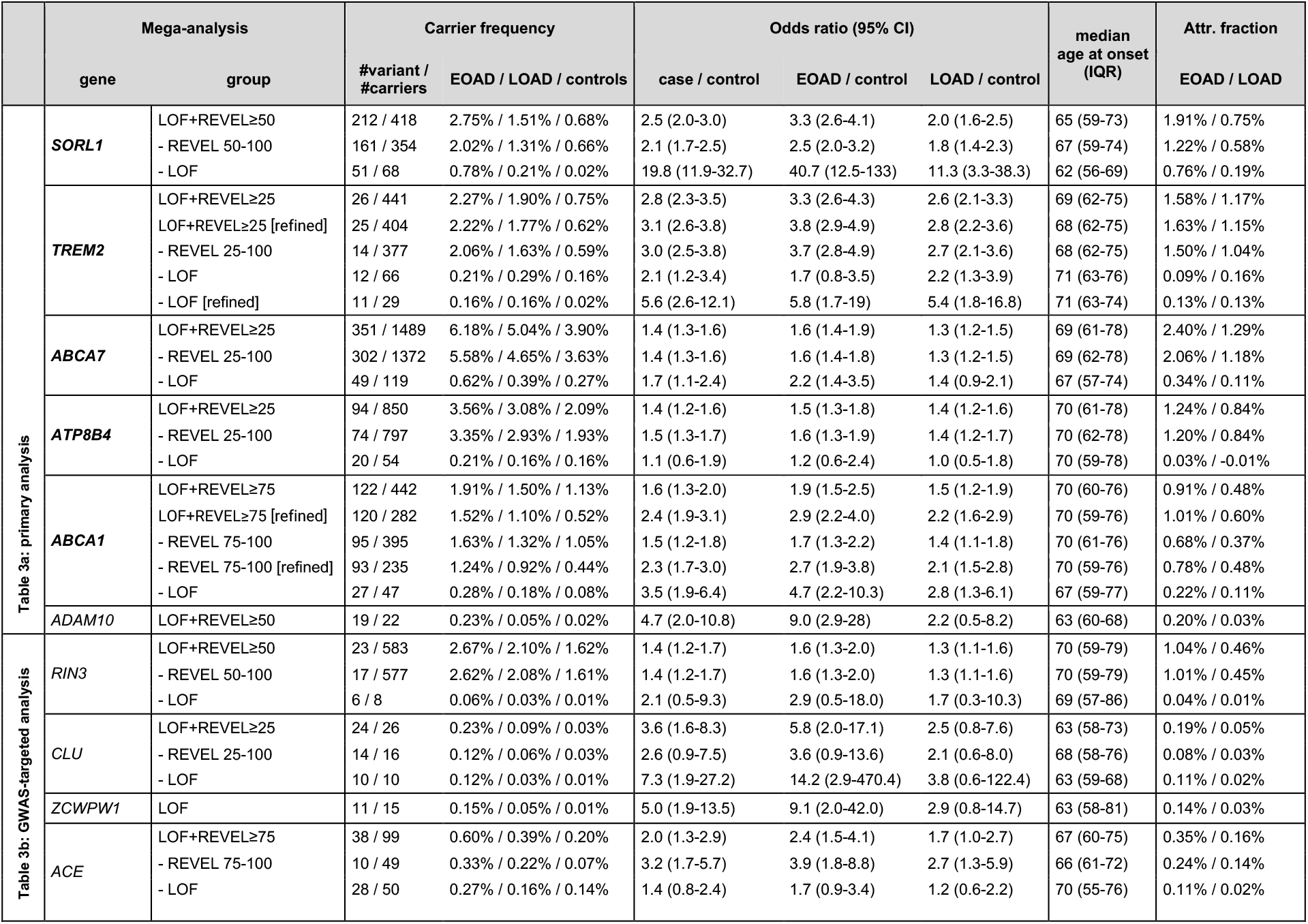
Mega Analysis; effect-sizes and p values. Per gene, the characteristics are shown for the variant deleteriousness threshold with the most evidence for AD association. For genes with sufficient carriers, signals are shown for LOF and missense variants separately. Variants contributing to the burden were validated in a multi-stage analysis (**Table S12**, Online Methods), which resulted in the construction of a *refined* burden for *TREM2* (1 variant removed) and *ABCA1* (2 variants removed). The attributable fraction of a gene is an estimate of the fraction of EOAD and LOAD cases in this sample that have become part of this dataset due to carrying a rare damaging variant in the respective gene (Online methods). Note that several variants were excluded from this analysis (i.e., due to differential missingness) that would otherwise have been included in the burden. See **section 1.12 of the supplement** for a gene-specific discussion of the variants that contribute to the association with AD, and gene-specific **Tables SG1-SG10** for the list of variants considered in the burden-analysis. P values for the mega-analysis are shown in **Table S14**.

**Fig 2:**
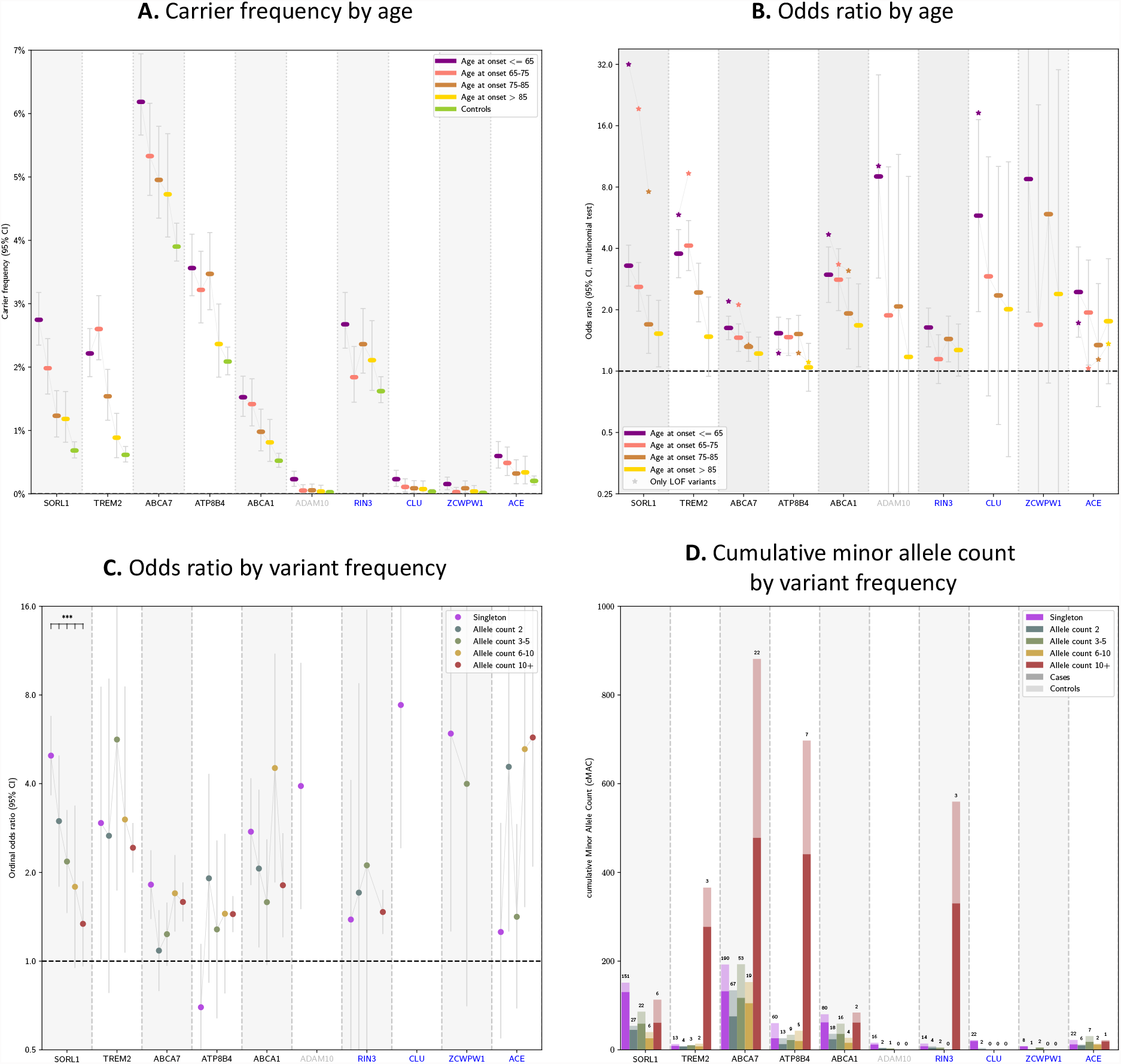
Characterization of gene specific variant features based on the mega-sample. We considered the deleteriousness threshold that provides the most evidence for AD-association (**Table 3**, refined burden). **A. Carrier frequency by age at onset**. A carrier is an individual who carries at least one damaging variant in the considered gene. **B. Odds ratios by age at onset**. The effect-size significantly decreased with age at onset for *SORL1, TREM2, ABCA7, ABCA1, ADAM10* (after multiple testing correction, **Table S7**). **C. Odds ratios by variant frequency**. The rareness of variants in *SORL1* significantly associated with the effect size (**Table S9**). **D. Cumulative minor allele count by variant frequency**: the total number of cases (dark) and controls (light) that carry gene-variants with allele frequencies as observed in the mega-sample. Numbers above the bars indicate the number of contributing variants. Whiskers: 95% CI.

**Fig 3.**
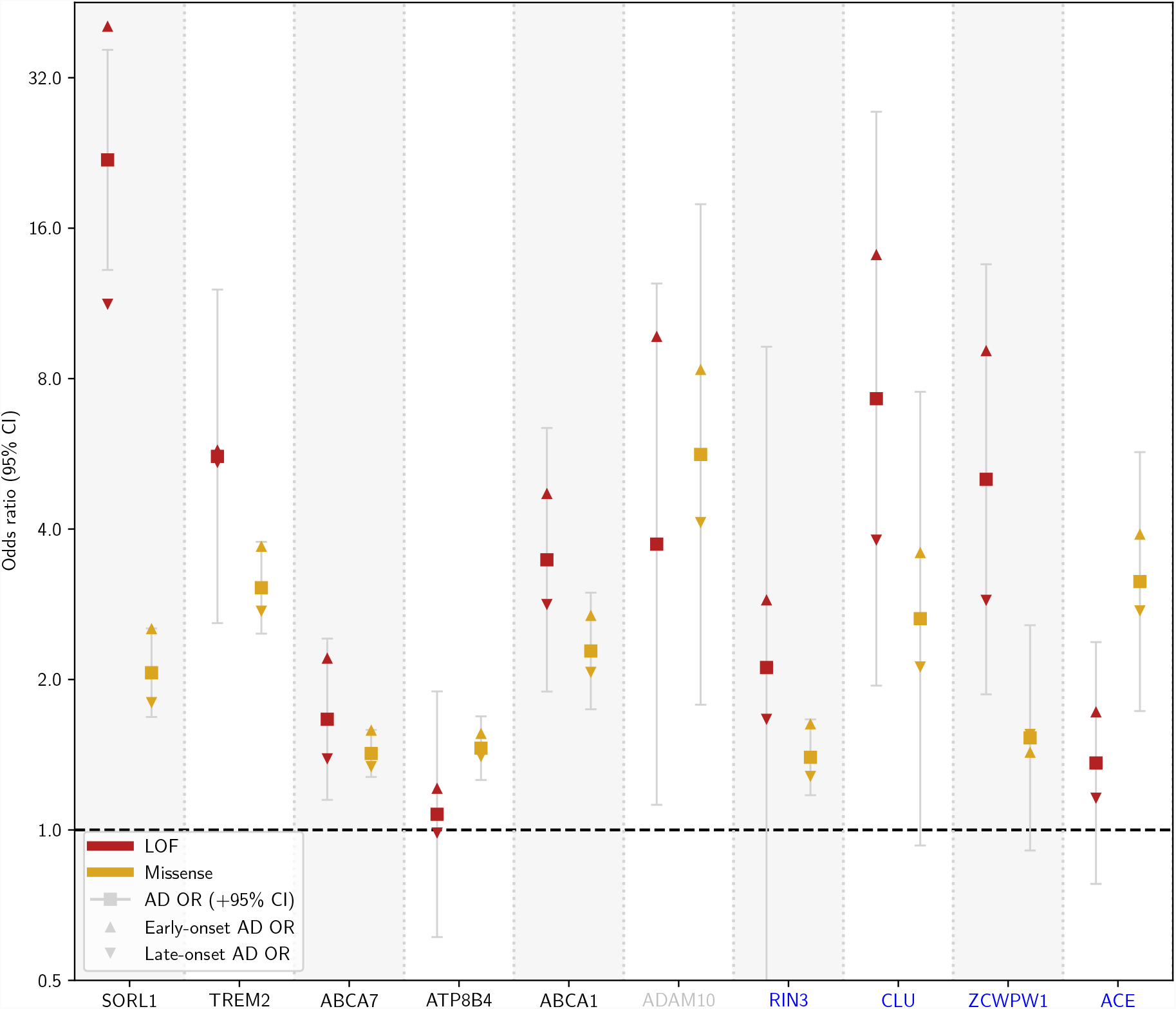
Summary Figure: Odds ratio by age at onset and variant pathogenicity. Odds ratio’s for LOF (red) and missense (yellow) variants. Case/control OR (square, 95% CI), EOAD OR (triangle pointing upwards), LOAD OR (triangle pointing downwards). Missense variants in the considered gene appertained to the variant-deleteriousness threshold that provides the most evidence for its AD-association (**Table 3, refined**). The LOF burden effectsize was significantly larger than the missense burden effect-size in the *SORL1* and we observed similar trends in *ABCA7* and *ABCA1* (**Table S9**, Supplementary Methods). Of note: missense variants in *ZCWPW1* did not contribute to the most significant burden but were shown here for reference purposes (REVEL>25).

Extremely rare variants contributed more to large effect sizes than less rare variants (p=0.03, **Table S8**). Indeed, for *SORL1*, the variants with the lowest variant frequencies had the largest effect-sizes (**Figures 2C, Table S9**), and damaging variants in *ADAM10, CLU* and *ZCWPW1* were all extremely rare (**Figures 2D**). Conversely, we observed that rare but recurrent variants contributed to the AD-association of *TREM2, ABCA7, ATP8B4* and *RIN3* (**Figure 2D**). The effect-sizes of rare, coding variant-burdens were large compared to the effect-sizes of the GWAS sentinel SNPs (**Table S5, S6**). Up to 18% EOAD and 14% LOAD cases carried at least one predicted damaging variant in one of the 10 genes, compared to 9% of the controls (**Table S10**). The fractions of EOAD cases in our sample that could be attributed to a rare variant in a specific gene ranged between 0.1% to 2.4%: (∼2%: *SORL1, TREM2, ABCA7*; ∼1%: *ATP8B4, ABCA1, RIN3*; and <0.5% for the remaining genes), and for LOAD cases this ranged between 0.0% to 1.3% (**Table 3**).

We performed an age-matched sensitivity analysis to investigate possible effects from other age-related conditions, which supported a role in AD for all 10 identified genes (**Figure S2**). Since *APOE* status was used as selection criterion in several contributing datasets, burden tests were not adjusted for *APOE-ε4* dosage; in a separate analysis we observed no interaction-effects between the rare-variant AD-association and APOE-ε4 dosage (**Table S11, Online Methods**). Also, the rare-variant burden-association was not confounded by somatic mutations due to age-related clonal hematopoiesis (**Table S12**).

Together, we report *ATP8B4* and *ABCA1* as novel AD risk factors with exome-wide significance and we report suggestive evidence for the association of rare variants in the *ADAM10* gene with AD risk. Furthermore, we identified *RIN3, CLU, ZCWPW1* and *ACE* as potential drivers in GWAS loci, illustrating how analyses of rare protein-modifying variants can solve this drawback of GWAS studies^20^. Larger datasets will be required to further confirm these signals. Given the association of LOF variants with increased AD-risk, we suggest that the GWAS risk alleles in the respective loci might also be associated with reduced activity of the gene, which will have to be evaluated in further experiments. We observed an increased burden of rare damaging genetic variants in individuals with an earlier age at onset. Nevertheless, damaging variants (including APOE-ε4/ε4) were observed in only 30% of the EOAD cases (**Table S10**), suggesting that additional damaging variants remain to be discovered (**Figure 1B**). Further, the effect of structural variants such as CNVs and repetitive sequences will need to be investigated in future analyses.

The associated genes strengthen our current understanding of AD pathophysiology. When treatment options become available in the future, identification of damaging variants in these genes will be of interest to clinical practice.

## Online methods

In-depth descriptions of all methods are described in the Supplemental Methods.

### Sample processing, genotype calling and quality control (QC)

We collected the exome, WGS or exome-extract sequencing data of a total of 52,361 individuals, brought together by the Alzheimer Disease European Sequencing consortium (ADES), the Alzheimer’s Disease Sequencing Project (ADSP)^21^ and several independent study-cohorts (**Table S1)**. Exome-extract samples only contained the raw reads that cover the 10 genes identified in Stage-1. Across all cohorts, AD cases were defined according to NIAA criteria^22^ for possible or probable AD or according to NINCDS-ADRDA criteria^23^ depending on the date of diagnosis. When possible, supportive evidence for an AD pathophysiological process was sought (including CSF biomarkers) or the diagnosis was confirmed by neuropathological examination (**Table S1**). AD cases were annotated with the age at onset or age at diagnosis (2014 samples), otherwise, samples were classified as late onset AD (366 samples). Controls were not diagnosed with AD. All contributing datasets were sequenced using a paired-end Illumina platform, different exome capture kits were used, and a subset of the sample was sequenced using whole genome sequencing (**Table S2**).

A uniform pipeline was used to process both the Stage-1 and Stage-2 datasets. Raw sequencing data from all studies were processed relative to the GRCh37 reference genome, read alignments of possible chimeric origin were filtered, and a GATK-based pipeline was used to call variants, while correcting for estimated sample contamination percentages. Samples were included in the datasets after they passed a stringent quality control pipeline: samples were removed when they had high missingness, high contamination, a discordant genetic sex annotation, non-European ancestry, high numbers of novel variants (w.r.t. to DBSNP v150), deviating heterozygous/homozygous or transition/transversion ratios. Further, we removed family members up to the 3^rd^ degree, and individuals who carried a pathogenic variant in *PSEN1, PSEN2, APP* or in other genes causative for Mendelian dementia diseases (Stage-1 only), or when there was clinical information suggestive of non-AD dementia. Variants considered in the analysis also passed a stringent quality control pipeline: multi-allelic variants were split into bi-allelic variants, variants that were in complete linkage and near each other were merged. Further, we removed variants that had indications of an oxo-G artifact, were located in Short Tandem Repeat (STR) and/or Low Copy Repeat (LCR) regions, had a discordant balance between reads covering the reference and alternate allele, had a low depth for alternate alleles, deviated significantly from Hardy-Weinberg equilibrium, were considered false positive based on GATK VQSR, or were estimated to have a batch effect. Variants with >20% genotype missingness (read depth <6) and differential missingness between the EOAD, LOAD and control groups were removed. To account for uncertainties resulting from variable read coverage between samples, we analyzed variants according to genotype posterior likelihoods, i.e., the likelihood for being homozygous for the reference allele, heterozygous or homozygous for the alternate allele. To account for genotype uncertainty, the burden test was performed multiple times with independently sampled genotypes and the average p-value across these tests is reported.

### Variant prioritization and thresholds

We selected variants in autosomal protein-coding genes that were part of the Ensembl basic set of protein coding transcripts (Gencode v19/v29^24^, see Supplement) and that were annotated by the Variant Effect Predictor (VEP) (version 94.542)^25^. Only protein coding missense and loss-of-function (LOF) variants were considered (LOF: nonsense, splice acceptor/donor or frameshifts). Missense and LOF variants were required to have respectively a ‘moderate’ and ‘high’ VEP impact classification. Then, missense variants were prioritized using REVEL (Rare Exome Variant Ensemble Learner^26^, annotation obtained from DBNSFP4.1a ^27^ and LOF variants were prioritized using LOFTEE (version 1.0.2)^28^. For the analysis we considered only missense variants with a REVEL score ≥ 25 (score range 0 – 100) and LOF variants annotated ‘high-confidence’ by LOFTEE. Variants were required to have at least one carrier (i.e. at least one one sample with a posterior dosage of >0.5), and a minor allele frequency (MAF) of <1%, both in the considered dataset and the gnomAD v2.1 populations (non-neuro set).

### Gene burden testing

The burden analysis was based on four deleteriousness thresholds by incrementally including variants from categories with lower levels of predicted variant deleteriousness: respectively LOF, LOF + REVEL≥75, LOF + REVEL≥50, and LOF + REVEL≥25. This allowed us to identify the variant-threshold providing maximum evidence for a differential burden-signal. To infer any dependable signal for a specific deleterious threshold, a minimum of 10 damaging alleles appertaining to this deleteriousness threshold was required: i.e., a cumulative minor allele count (cMAC) ≥10. Multiple testing correction was performed across all performed tests (up to 4 per gene). Burden testing was implemented using ordinal logistic regression. This enabled the burden testing to particularly weight EOAD cases, as previous findings indicated that high-impact variants are enriched in early onset (EOAD) cases relative to late onset (LOAD) cases^29^. This implies that the burden of high-impact deleterious genetic variants is ordered according to: burden_EOAD_ > burden_LOAD_ > burden_control_. Ordinal logistic regression enabled optimal identification of such signals, while also allowing the detection of EOAD-specific burdens (burden_EOAD_ > burden_LOAD_ ∼ burden_control_) and regular case-control signals (burden_EOAD_ ∼ burden_LOAD_ > burden_control_). For protective burden signals, the order of the signals is reversed, such that burden_EOAD_ < burden_LOAD_ < burden_control_. We considered an additive model, while correcting for 6 population covariates, estimated after removal of population outliers. P-values were estimated using a likelihoodratio test. Genes were selected for confirmation in Stage-2 if the False Discovery Rate for AD association was <0.1 in Stage-1 (Benjamini-Hochberg procedure^30^). For the GWAS targeted analysis, a more stringent threshold was used (FDR<0.05) due to the absence of a separate confirmation stage. For the meta-analysis, genes were considered significantly associated with AD when the corrected P was <0.05 after family-wise correction using the Holm-Bonferroni procedure^31^. Effect-sizes (odds ratios, ORs) of the ordinal logistic regression can be interpreted as weighted averages of the OR of being an AD case versus control, and the OR of being an early-onset AD case or not. To aid interpretation, we additionally estimated ‘standard’ case/control ORs across all samples, per age category (EOAD versus controls and LOAD versus controls), and for age-at-onset categories ≤65 (EOAD), (65-70], (70-80] and >80 using multinomial logistic regression, while correcting for 6 PCA covariates.

### GWAS driver gene identification

For the 75 loci identified in the most recent GWAS^3^, genes were selected for burden testing based on earlier published gene prioritizations. First, gene prioritizations were obtained from Schwarzentruber et al.^32^ for 33 known loci. For 28 remaining loci, we obtained the Tier 1 prioritization from Bellenguez et al. ^3^, and for the loci without prioritization candidates (14 loci), we selected the nearest gene. In total, 81 protein-coding genes were selected (**Table S5**), of which 67 genes had sufficient damaging allele carriers to be tested for at least one variant selection threshold. Gene burden testing was performed as described above, and multiple testing correction to identify potential driver genes was performed using the Benjamini-Hochberg procedure, with a cutoff of 5%.

### Validation of variant selection

We validated the REVEL variant impact prediction for missense and the LOFTEE impact prediction for LOF variants, for all variants with a MAF < 1%, for which there were at least 15 damaging allele carriers. For protein-modifying variants that were not in the most significant burden selection of a gene due to a low predicted impact, we investigated whether they, nevertheless, showed a significant AD-association (based on a case/control analysis using logistic regression). Vice versa, for variants that were in the burden selection, we investigated whether their effect-size was significantly reduced or oppositely directed from other missense or LOF variants in the burden selection (Fisher exact test). Individual variant-effects were analyzed in the Stage-1 dataset, followed by a confirmation analysis in the Stage-2 dataset. Multiple testing correction was performed per gene, with FDR<0.1 used as threshold for Stage-1 and Holm-Bonferroni (P<0.05) for Stage-2.

### Descriptive measures

A variant-carrier was defined as an individual for whom the summed dosage of *all* the variants in the considered variant category is ≥0.5 (Supplementary methods). Carrier frequencies (CFs) were determined as #carriers / #total samples. Attributable fraction for cases in an age group was estimated as the probability of a case with an age-at-onset in age window *i* being exposed to a specific gene gene burden (*CF*_*case,gene,i*_), multiplied by an estimate of the attributable fraction among the exposed for these cases: 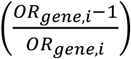 with the odds ratio being an approximation of the relative risk)^33,34^. For large effect-sizes, this estimate approaches the difference in carrier frequency between cases and controls: (*CF*_*case,gene,i*_)−(*CF*_*control,gene*_).

### Sensitivity analyses

We determined if observed effects could be explained by age differences between cases and controls. We constructed an age-matched sample, dividing samples in strata based on age/age-at-onset, with each stratum covering 2.5 years. Case/control ratios in all strata were kept between 0.1 and 10 by down sampling respectively controls or cases. Subsequently, samples were weighted using the propensity weighting within strata method proposed by Posner and Ash^35^. Finally, a case-control logistic regression was performed both on the unweighted and weighted case-control labels, and estimated odds ratios and confidence intervals were compared (**Figure S2**). Also, we determined if somatic mutations due to age-related clonal hematopoiesis could have confounded the results. We calculated for all heterozygous calls in the burden selection the balance between reference and alternate reads, and compared these to reference values (**Table S12**). While APOE was not included as a confounder, we performed a separate APOE interaction analysis (**Table S11**), through a likelihood ratio test between a model: label∼gene_burden_score+apoe_e4_dosage and an interaction model: label∼gene_burden_score+apoe_e4_dosage+apoe_e4_dosage * gene_burden_score. This test was performed on a reduced dataset, from which datasets in which APOE status was used as selection criterion were removed.

### Power analysis

Power calculations were performed for ordinal logistic regression and Firth logistic regression (casecontrol and EOAD vs. rest), **Figure 1B** and **Table S4**. Given odds ratios for EOAD and LOAD cases, and the cumulative minor allele count per gene, we sampled the number of alleles in EOAD cases, LOAD cases and controls according to a multinomial distribution. We randomized these allele-carriers across the dataset, and performed the burden test (as described above). Power for genes with cMAC <10 was set to 0, as these genes were not analyzed.

## Supporting information

Holstege Supplement

## Data Availability

Genetic variants of identified genes, and summary statistics of the rare variant burden in the Stage-1 AD cases/controls comparison will be made available upon journal publication.

## Data availability Statement

The genetic variants analyzed in this study will be included in the Supplementary data files (Tables SG1-SG10) upon the formal publication of this manuscript. Summary statistics of the Stage-1 analysis will be made available through [**holstegelab.eu/data**]. The full datasets generated during and/or analyzed during the current study are not publicly available due to privacy restrictions applicable to genetic data from human subjects.

## Code availability statement

All software and algorithms used in the analysis are described in the Supplement attached to this Letter. Self-contained code can be found at **holstegelab.eu/tools**.

