## Supplementary material for "Exome sequencing identifies rare damaging variants in *ATP8B4* and *ABCA1* as novel risk factors for Alzheimer’s Disease": Holstege Supplement

*Authors contributed equally to this work

[#] To whom correspondence should be addressed

1. Genomics of Neurodegenerative Diseases and Aging, Department of Human Genetics, Amsterdam University Medical Center (location VUmc), Neuroscience Campus Amsterdam, Amsterdam, The Netherlands
2. Alzheimer Center Amsterdam, Department of Neurology, Amsterdam Neuroscience, Vrije Universiteit Amsterdam, Amsterdam UMC, Amsterdam, The Netherlands
3. Delft Bioinformatics Lab, Delft University of Technology, Delft, The Netherlands
4. Normandie Univ, UNIROUEN, Inserm U1245 and CHU Rouen, Department of Genetics and CNRMAJ, F-76000 Rouen, France
5. Univ. Lille, Inserm, CHU Lille, Institut Pasteur de Lille, U1167-RID-AGE facteurs de risque et déterminants moléculaires des maladies liés au vieillissement, Lille, France
6. MRC Centre for Neuropsychiatric Genetics and Genomics, , Division of Psychological Medicine and Clinical Neuroscience, School of Medicine, Cardiff University, Cardiff, UK
7. Department of Neurology, Erasmus Medical Centre, Rotterdam, The Netherlands
8. Department of Internal Medicine, Erasmus Medical Centre, Rotterdam, The Netherlands
9. Department of Epidemiology, Erasmus Medical Centre, Rotterdam, The Netherlands
10. LACDR, Leiden, The Netherlands
11. Nuffield Department of Population Health Oxford University
12. MRC Prion Unit at UCL, UCL Institute of Prion Diseases, London, UK
13. Department of Neurology, II B Sant Pau, Hospital de la Santa Creu i Sant Pau, Universitat Autònoma de Barcelona, Barcelona, Spain
14. CIBERNED, Network Center for Biomedical Research in Neurodegenerative Diseases, National Institute of Health Carlos III, Madrid, Spain
15. Division of Neurogenetics and Molecular Psychiatry, Department of Psychiatry and Psychotherapy, Faculty of Medicine and University Hospital Cologne, University of Cologne, Cologne Germany
16. ADNI consortium; see supplemental authors
17. The John P. Hussman Institute for Human Genomics, University of Miami, Miami, Florida, USA
18. Univ Montpellier, Inserm, INM (Institute for Neurosciences of Montpellier), Montpellier, France
19. Cardiovascular Health Research Unit, Department of Medicine, University of Washington, Seattle, WA, USA.
20. Université Paris-Saclay, CEA, Centre National de Recherche en Génomique Humaine Evry, France
21. Experimental Neuro-psychobiology Laboratory, Department of Clinical and Behavioral Neurology, IRCCS Santa Lucia Foundation, Rome, Italy
22. Department of Neurodegenerative Science, Van Andel Institute, Grand Rapids, MI USA
23. Division of Psychiatry and Behavioral Medicine, Michigan State University College of Human Medicine, Grand Rapids, MI, USA.
24. HudsonAlpha Institute for Biotechnology, Huntsville, AL, USA
25. Department of Neuroscience, Catholic University of Sacred Heart, Fondazione Policlinico Universitario A. Gemelli IRCCS, Rome, Italy
26. University Bordeaux, Inserm, Bordeaux Population Health Research Center, France
27. Department of Neurology, Bordeaux University Hospital, Bordeaux, France
28. UKDRI@ Cardiff, School of Medicine, Cardiff University, Cardiff, UK
29. Department of Biostatistics, Boston University School of Public Health, Boston, MA, USA.
30. Framingham Heart Study, Framingham, MA, USA.
31. Department of Neurology, Boston University School of Medicine, Boston, MA, USA.
32. Department of Epidemiology, Boston University, Boston, MA, USA
33. Department of Medicine (Biomedical Genetics), Boston University, Boston, MA, USA
34. Neurogenomics and Informatics Center, Washington University School of Medicine, St Louis, MO USA
35. Psychiatry Department, Washington University School of Medicine, St Louis, MO
36. Hope Center for Neurological Disorders, Washington University School of Medicine, St Louis, MO
37. Dementia Research Centre, UCL Queen Square Institute of Neurology, London, UK
38. Fondazione IRCCS Ca' Granda, Ospedale Policlinico, Milan, Italy
39. University of Milan, Milan, Italy
40. Univ Brest, Inserm, EFS, CHU Brest, UMR 1078, GGB, F-29200, Brest, France
41. Genome Diagnostics, Department of Human Genetics, VU University, AmsterdamUMC (location VUmc), Van Der Boechorststraat 7, 1081 BT Amsterdam
42. Department of Neurology and Neurological Sciences, Stanford University, Stanford, CA, USA.
43. Department of Epidemiology and Biostatistics, Case Western Reserve University, Cleveland, Ohio, USA
44. Clinical and Experimental Science, Faculty of Medicine, University of Southampton, Southampton, UK.
45. Department of Complex Trait Genetics, Center for Neurogenomics and Cognitive Research, Amsterdam Neuroscience, Vrije University, Amsterdam, The Netherlands.
46. McGill University and Genome Quebec Innovation Centre, Montreal, QC, Canada
47. Department of Human Genetics, Amsterdam UMC, University of Amsterdam, Amsterdam Reproduction and Development Research Institute Amsterdam
48. Dr. John T. Macdonald Foundation Department of Human Genetics, University of Miami, Miami, Florida, USA
49. Institute of Neurology, Catholic University of the Sacred Heart , Rome, Itlay
50. Taub Institute on Alzheimer's Disease and the Aging Brain, Department of Neurology, Columbia University, New York, New York, USA
51. Gertrude H. Sergievsky Center, Columbia University, New York, New York,
52. Institute of Gerontology and Geriatrics, Department of Medicine and Surgery, University of Perugia, Perugia, Italy
53. Human Genetics, School of Life Sciences, University of Nottingham, UK
54. Department of Neuroscience, Psychology, Drug Research and Child Health University of Florence, Florence Italy
55. IRCCS Fondazione Don Carlo Gnocchi, Florence, Italy
56. Penn Neurodegeneration Genomics Center, Department of Biostatistics, Epidemiology, and Informatics; University of Pennsylvania Perelman School of Medicine, Philadelphia, Pennsylvania,
57. Penn Neurodegeneration Genomics Center, Department of Pathology and Laboratory Medicine, University of Pennsylvania Perelman School of Medicine, Philadelphia, Pennsylvania,

**58.** Genomic And Molecular Epidemiology (GAME) Lab, School of Biosciences and Veterinary Medicine, University of Camerino (UNICAM) Camerino, 62032, Italy
**59.** Univ. Lille, Inserm, CHU Lille, UMR1172, Resources and Research Memory Center (MRRC) of Distalz, Licend, Lille France
**60.** Fundació Docència i Recerca MútuaTerrassa and Movement Disorders Unit, Department of Neurology, University Hospital MútuaTerrassa, Barcelona, Spain
**61.** Memory Disorders Unit, Department of Neurology, Hospital Universitari Mutua de Terrassa, Terrassa, Barcelona, Spain.
**62.** Université de Nantes, CHU Nantes, CNRS, INSERM, l'institut du thorax, Nantes, France
**63.** Institute of Social Medicine, Occupational Health and Public Health, University of Leipzig, Leipzig, Germany.
**64.** Neurology Service, Marqués de Valdecilla University Hospital (University of Cantabria and IDIVAL), Santander, Spain.
**65.** Laboratory for Advanced Hematological Diagnostics, Department of Hematology and Stem Cell Transplant, Lecce, Italy
**66.** Department of Psychiatry and Glenn Biggs Institute for Alzheimer's and Neurodegenerative Diseases, San Antonio, TX, USA
**67.** Laboratory of Neuropsychiatry, Department of Clinical and Behavioral Neurology, IRCCS Santa Lucia Foundation, Rome, Italy
**68.** Department of Neurodegenerative Diseases and Geriatric Psychiatry, University Hospital Bonn, Medical Faculty, Bonn, Germany.
**69.** German Center for Neurodegenerative Diseases (DZNE, Bonn), Bonn, Germany.
**70.** Normandie Univ, UNIROUEN, Inserm U1245 and CHU Rouen, Department of Neurology and CNRMAJ, F-76000 Rouen, France
**71.** Reta Lila Weston Research Laboratories, Department of Molecular Neuroscience, UCL Institute of Neurology, London, UK.
**72.** Memory and Aging Center, Department of Neurology, University of California, San Francisco, CA USA
**73.** Cluster of Excellence Cellular Stress Responses in Aging-Associated Diseases (CECAD), University of Cologne, Cologne, Germany

### INDEX

1 SUPPLEMENTAL METHODS 9

**1.1 Sample descriptions 9**

1.1.1 ADES-FR 9
1.1.2 AgeCoDe-UKBonn 11
1.1.3 Barcelona- SPIN 12
1.1.4 AC-EMC 13
1.1.5 ERF 13
1.1.6 Rotterdam Study 14
1.1.7 ADC-Amsterdam 14
1.1.8 Netherlands Brain Bank 15
1.1.9 UMC-Amsterdam 15
1.1.10 100-plus Study 15
1.1.11 EMIF-AD 90-plus Study 16
1.1.12 CBC: Control Brain Consortium 16
1.1.13 PERADES 16
1.1.14 StEP-AD 17
1.1.15 Knight-ADRC 18
1.1.16 UCSF/NYGC/UAB 18
1.1.17 UCL-DRC EOAD 19
1.1.18 ADSP 19

**1.2 Sequence read alignment and variant calling 21**

1.2.1 Exome sequence read processing 21
1.2.2 Processing of WGS reads and exome extracts 22
1.2.3 Chimeric read declipping 23

**1.3 Sample QC 25**

1.3.1 Missingness 25
1.3.2 Contamination 25
1.3.3 Sex-check 25
1.3.4 Population outliers 26
1.3.5 Excess novel SNPs 26
1.3.6 Excess novel indels 26
1.3.7 Heterozygous/homozygous (Het/Hom) and transition/transversion ratios (Ts/Tv) 27

1.3.8 Identity By Descent (IBD) analysis 27
1.3.9 Bad PCR plates 28
1.3.10 Removal of dementia-related (likely) pathogenic variant-carriers 28
1.3.11 AD label 28
1.3.12 Handling of exome-extract samples 28
1.3.13 Merging of Stage-1 and -2 sample QC 29

**1.4 Variant QC 29**
1.4.1 Multi allelic variants 29
1.4.2 Variant merging 29
1.4.3 Oxo-G 30
1.4.4 Short Tandem Repeat (STR) and Low Copy Repeats (LCR) regions 30
1.4.5 Allele Balance 30
1.4.6 Depth Fraction 30
1.4.7 Hardy Weinberg 30
1.4.8 VQSR 31
1.4.9 Pre-variant QC versus final variant QC 31
1.4.10 Variant Batch Detection 31

**1.5 Genotype posterior probabilities 31**
1.5.1 Genotype likelihoods 32
1.5.2 Posterior probability 32
1.5.3 Multi-allelic variants 33
1.5.4 Posterior sample QC-measures 33

**1.6 Oxo-G variant call filtering 34**
1.6.1 Oxo-G statistics 35
1.6.2 Full error model 36
1.6.3 Contrasting error model 36
1.6.4 Genotype likelihood calculation 36
1.6.5 Genotype and variant filtering 37

**1.7 PCA covariates 38**

**1.8 Variant batch detection and correction 38**
1.8.1 Examples of batch effects 39
1.8.2 Algorithm overview 39
1.8.3 Technical covariates 41

1.8.4 Non-technical covariates 42
1.8.5 Forward-backward covariate search 43
1.8.6 Prioritizing non-technical covariates 43
1.8.7 Diploid logistic regression model 44
1.8.8 Tree search for complex haploblock-markers 45
1.8.9 Detection of missing-not-at-random genotypes 46
1.8.10 Two-phase approach 46
1.8.11 Variant batch correction 47
1.8.12 Variant filtering 47

**1.9 Variant selection and annotation 47**
1.9.1 Protein coding transcripts. 48
1.9.2 Variant type. 48
1.9.3 Variant prioritization. 48
1.9.4 Variant frequency. 48
1.9.5 Variant missingness. 48
1.9.6 Variant categorization. 49

**1.10 Analyses and statistical tests 49**
1.10.1 Gene burden test 49
1.10.2 Variant impact thresholds 50
1.10.3 Odds ratios 51
1.10.4 Testing for an association between effect size and variant rareness 52
1.10.5 Sensitivity analysis 52
1.10.6 Variant-specific analysis 52

**1.11 Sanger Validation of identified variants 53**

**1.12 Detailed gene discussion 54**
*1.12.1 SORL1* 54
*1.12.2 TREM2* 55
*1.12.3 ABCA7* 56
*1.12.4 ATP8B4* 57
*1.12.5 ABCA1* 57
*1.12.6 ADAM10* 58
1.12.7 RIN3 58
*1.12.8 CLU* 59
*1.12.9 ZWCWP1* 59

*1.12.10 ACE* 60

**2 SUPPLEMENTAL FIGURES AND TABLES 61**

**2.1 Figures 61**

2.1.1 Figure S1: Age, gender, APOE genotype distribution 61
2.1.2 Figure S2: Sensitivity Analysis: AD vs Age association 62
2.1.3 Figure S3: Read length per study 63
2.1.4 Figure S4: Genotype Quality 64
2.1.5 Figure S5: Genetic sex 65
2.1.6 Figure S6: PCA: Sample population compared to 1,000G population samples 66
2.1.7 Figure S7: first two population PCA components per study 67
2.1.8 Figure S8: Third and fourth population PCA components per study 68
2.1.9 Figure S9: Number of novel SNPs (union of capture kits) 69
2.1.10 Figure S10: Number of novel indels (union of capture kits) 70
2.1.11 Figure S11: Number of novel SNPs (intersection of capture kits) 71
2.1.12 Figure S12: Number of novel indels (intersection of capture kits) 72
2.1.13 Figure S13: Ts/Tv ratio known variants (intersection capture kits) 73
2.1.14 Figure S14: Ts/Tv ratio novel variants (intersection of capture kits) 74
2.1.15 Figure S15: Het/Hom ratio known variants (intersection capture kits) 75
2.1.16 Figure S16: First two PCA components per study, after sample QC. 76
2.1.17 Figure S17: Third and fourth PCA components per study, after sample QC. 77
2.1.18 Figure S18: Fifth and sixth PCA components per study, after sample QC. 78
2.1.19 Figure S19: Inflation in Stage-2 79
2.1.20 Figure S20: Inflation in the mega-analysis dataset 80
2.1.21 Figure S21: Frequency in controls by age last seen 81
2.1.22 Figure S22: Age-at-onset by variant category 82
2.1.23 Figure S23: Attributable fraction per gene and age-at-onset category 83

**2.2 Supplemental Tables 84**

2.2.1 Table S1: Contributing Studies 85
2.2.2 Table S2: Capture Kits 86
2.2.3 Table S3 Sample QC and Variant QC 87
2.2.4 Table S4: Power in stage-1 90
2.2.5 Table S5: List of genes and tests performed for the targeted GWAS analysis 92
2.2.6 Table S6: Burden testing of prioritized genes in GWAS loci. 99
2.2.7 Table S7 Age burden trends in cases and controls separately 100
2.2.8 Table S8 Contribution of extremely rare variants to the burden test 101

2.2.9 Table S9 Variant features 102
2.2.10 Table S10 Carriers of multiple variants in identified genes 103
2.2.11 Table S11: Testing for interaction with APOE - E4 genotype 104
2.2.12 Table S12 Somatic Mutation Check 105
2.2.13 Table S13: Validation of variant selection 106
2.2.14 Table S14 107
2.2.15 Tables SG1-10 107

**3 ACKNOWLEDGMENTS 108**

**3.1 Study participants and personnel involved in sample collection 108**

**3.2 SURF supercomputer facility 108**

**3.3 Study Cohorts 108**
3.3.1 ADES-FR 108
3.3.2 AgeCoDe-UKBonn 109
3.3.3 Barcelona- SPIN 109
3.3.4 AC-EMC 109
3.3.5 ERF 109
3.3.6 Rotterdam Study 109
3.3.7 ADC-Amsterdam 110
3.3.8 100-plus Study 111
3.3.9 EMIF-90+ 111
3.3.10 CBC: Control Brain Consortium 111
3.3.11 PERADES 111
3.3.12 StEP-AD cohort 113
3.3.13 Knight-ADRC 113
3.3.14 UCSF/NYGC/UAB 113
3.3.15 UCL-DRC EOAD 113
3.3.16 ADSP 113

**4 SUPPLEMENTAL AUTHORS 118**
4.1.1 PERADES Cohort: 118
4.1.2 StEP AD investigators 119
4.1.3 Knight ADRC investigators 119
4.1.4 ADNI database 119

**5 REFERENCES 119**

### 1 Supplemental methods

#### 1.1 Sample descriptions

We analyzed a total sample of 52,361 individuals sequenced with Illumina technology. Of these, 24,510 individuals (18,403 after QC) were collected as part of the Alzheimer Disease European Sequencing consortium (ADES), comprising 15 studies from Germany, France, The Netherlands, Spain, Italy, and the United Kingdom. All studies were approved by the ethics committees of respective institutes, and all participants provided informed consent for study participation. These samples were combined with 27,851 samples from the USA (14,155 after QC), the majority of which were from the Alzheimer's Disease Sequencing Project (ADSP), which were described previously[1] (**Table S1**).

Across all studies, AD cases were defined according to NIAA criteria[2] for possible or probable AD or according to NINCDS-ADRDA criteria[3] depending on the date of diagnosis. When possible, supportive evidence for an AD pathophysiological process was sought (including CSF biomarkers) or the diagnosis was confirmed by neuropathological examination (**Table S1**). Cases were annotated with the age at onset or age at diagnosis (2014 samples), otherwise, samples were classified as late onset AD (366 samples). Controls were not diagnosed with AD. All contributing datasets were sequenced using a paired-end Illumina platform, but different exome capture kits were used, and a subset of the sample was sequenced using whole genome sequencing (**Figure S1, Table S1**).

##### 1.1.1 ADES-FR

The ADES-FR project combines WES and WGS data from AD cases and controls from France[4]. Part of the patients are from the CNRMAJ-Rouen center (n=921) and patient ascertainment is described in detail in Nicolas et al.[5] including an update of the inclusions by the French National network CNR-MAJ (national reference center for young Alzheimer patients). Briefly, unrelated cases with early-onset AD (age at onset ≤65 years) from

France were recruited among patients who fulfilled the NIAA criteria[2]. The clinical examination included personal medical and family history assessment, neurologic examination, neuropsychological assessment, and neuroimaging. In addition, cerebrospinal fluid (CSF) biomarkers indicative of AD were available for 67% of the cases. Cases with CSF biomarkers not consistent with AD diagnostics were excluded. A positive family history (i.e., at least a secondary case among first- or second-degree relatives, whatever the age of onset) was present in 45% of cases. Patients were either screened by Sanger sequencing and QMPSF for pathogenic variants in *APP*, *PSEN1* or *PSEN2* prior to WES or by the interpretation of WES data or both. Carriers of pathogenic variants were not included for WES or were secondarily excluded following WES analysis so that none of the CNRMAJ-Rouen patients included in this work prior to shared analyses is a carrier of a pathogenic variant in *APP*, *PSEN1*, *PSEN2* as well as in a list of Mendelian dementia causative genes[6]. In addition, some controls were recruited directly from the CNRMAJ (n=30). Another large part of the samples was from the European Alzheimer's Disease Initiative (EADI) dataset[7]. This study combined clinical prevalent and incident cases of AD (n=1,121) (i) from Lille cross-sectional studies and (ii) from the Three-City (3C) study, a population-based, prospective study with 12-years of follow-up[8]. Diagnoses were established according to the DSM-III-R and NINCDS-ADRDA criteria[3]. Controls were selected among the 3C individuals not diagnosed with dementia after a 12-year follow-up (n=670). In addition, other controls were obtained from the FREX consortium[9]. These controls (n=576) were specifically designed from 6 French cities with the aim of studying and establishing the French population genetic structure of rare variants. Overall, the ADES-FR samples includes 2,042 AD cases (1,088 EOAD and 954 LOAD) and 1,276 controls. All patients and controls provided informed written consent for genetic analyses in a clinical and/or in a research setting, according to each study. In addition, the ethics committee of the Rouen University Hospital approved the use of retrospective data in the context of the ADES-FR project and with other ADES European and American partners (CERNI notifications 2017-015 and 2019-055).

For Stage-2, entire exomes of 529 independent and unrelated AD patients, including 384 patients from the ECASCAD study were included. All had CSF biomarkers consistent with AD (except two patients who had neuropathological confirmation), and 90% of them were

EOAD cases, the remaining 10% cases had an age of onset between 65 and 75 years. As controls, we extracted BAM files of the 11 genes selected in Stage-1 among the genome sequencing data from the FranceGenRef study. Individuals included in this study were selected based on the places of birth of their grandparents within France and at a maximum distance of 30 kilometers. A total of 862 individuals (274 females and 588 males) were sampled from three different studies: 50 individuals (25 females and 25 males) were blood donors sampled in the Finistère district, 354 individuals (177 females and 177 males) were blood donors from the PREGO biobank with ancestries in the other districts of Brittany (Côtes d'Armor, Ile-et-Vilaine, Morbihan) and in the 5 districts of the Pays-de-la-Loire region (Loire-Atlantique, Maine-et-Loire, Mayenne, Sarthe, Vendée), 458 individuals (72 females and 386 males) were volunteers from the GAZEL cohort (www.gazel.inserm.fr/en) who were selected among the volunteers who gave a blood sample and who answered a questionnaire on their parents and grandparents' places of birth. All individuals signed informed consent for genetic studies at the time they were enrolled and had their blood collected.

##### 1.1.2 AgeCoDe-UKBonn

The AgeCoDe-UKBonn sample was derived from the following two sources, the German study on Aging, Cognition, and Dementia in primary care patients (AgeCoDe, n=294) and the interdisciplinary Memory Clinic at the University Hospital of Bonn (UKBonn, n=100).
—**The German study on Aging, Cognition, and Dementia**: The AgeCoDe study is a multicenter prospective general practice-based cohort study since 2001, including community dwelling elderly aged 75 years or older that were recruited at six study sites (Bonn, Düsseldorf, Hamburg, Leipzig, Mannheim, and Munich). The AgeCoDe study was approved by the local ethics committees of the Universities of Bonn, Hamburg, Düsseldorf, Heidelberg/Mannheim, Leipzig, and Munich. Before participation written informed consents were collected from all subjects. The AgeCoDe study aims to identify risk factors and predictors of cognitive decline and dementia[10,11]. Participants were recruited from general practitioner (GP) registries. Inclusion criteria were an age of 75 and older, absence of dementia, one or more visits to the GP in the past year, no hearing or vision impairments

and German as a native language. Exclusion criteria were only home-based GP consultations, severe illness with a fatal outcome within 3 months and a language barrier. The baseline assessment including 3,327 subjects was completed between 2002 and 2003. After the baseline assessment 70 subjects were excluded due to presence of dementia after standard assessment and 40 subjects were excluded with an age below 75 years. Participants were interviewed for follow up every 18 months. All assessments are performed at the participant's home by a trained study psychologist or physician. At all visits, assessment includes the Structured Interview for Diagnosis of Dementia of Alzheimer type, Multi-infarct Dementia, and Dementia of other etiology according to DSM-IV and ICD-10 (SIDAM)[12]. The SIDAM comprises: (1) a 55-item neuropsychological test battery, including all 30 items of the MMSE and assessment of several cognitive domains (orientation, verbal and visual memory, intellectual abilities, verbal abilities/ calculation, visual–spatial constructional abilities, aphasia/ apraxia); (2) a 14-item scale for the assessment of the activities of daily living (SIDAM-ADL-Scale); and (3) the Hachinski Rosen-Scale. Dementia was diagnosed according to DSM-IV criteria. AgeCoDe provided DNA from 294 persons who progressed to late onset AD dementia at any follow up.

—**UKBonn**: The interdisciplinary Memory Clinic of the Department of Psychiatry and Department of Neurology at the University Hospital in Bonn provided early-onset AD patients (n=100). Diagnoses were assigned according the NINCDS/ADRDA criteria[3] and on the basis of clinical history, physical examination, neuropsychological testing (using the CERAD neuropsychological battery, including the MMSE), laboratory assessments, and brain imaging.

##### 1.1.3 Barcelona- SPIN

Neuropathological samples were obtained from the Neurological Tissue Bank of the Biobanc-HospitalClinic-IDIBAPS, and disease evaluation was performed according to international consensus criteria. Clinical samples were recruited from the multimodal Sant Pau Initiative on Neurodegeneration (SPIN) cohort (https://santpaumemoryunit.com/our-research/spin-cohort/)[13], and were evaluated at the Memory Unit at Hospital de Sant Pau (Barcelona). The repository includes clinical data of more than 6,000 participants, >2900 plasma samples, genetic material (DNA and RNA) of >3,200 and >400 subjects, respectively, and >2,000 CSF samples. All controls had normal cognitive scores in the formal neuropsychological evaluation and normal core CSF AD biomarkers, based on previously published cut-offs[14]. AD patients fulfilled clinical criteria of “probable AD dementia with evidence of the AD pathophysiological process”[3] and therefore had abnormal core AD biomarkers (low Aβ1–42 and high t-Tau or p-Tau) in the CSF. The original protocol and the subsequent amendments were approved by our local Ethics Committee at the Sant Pau Research Institute as well as the Committee of the Neurological Tissue Bank. The SPIN cohort is based on blinded enrollment and only clinically relevant biomarker results are disclosed.

##### 1.1.4 AC-EMC

The Alzheimer Center Erasmus MC cohort (AC-EMC) includes patient referred to the Department of Neurology of the Erasmus Medical Center (Rotterdam, the Netherlands). DNA samples from 125 patients with probable AD were included in the current study. The average age at onset was 60 years (range 41-77). A large fraction of the patients had a positive family history, defined as at least one first degree relative with dementia. All patients underwent clinical examination, neuropsychological assessment, neuroimaging, and if indicated, a lumbar puncture. The diagnosis was established according to the National Institute of Neurological and Communicative Disorders and Stroke-Alzheimer’s Disease and Related Disorders Association (NINCDS-ADRDA) criteria for AD[3].The study was approved by the Medical Ethical Committee of the Erasmus Medical Center, and written informed consent was obtained from all participants or their legal representatives.

##### 1.1.5 ERF

The Erasmus Rucphen Family (ERF) Study is a family-based cohort study that is embedded in the Genetic Research in Isolated Populations (GRIP) program in the South West of the Netherlands. The aim of this program was to identify genetic risk factors in the development of complex disorders. For the ERF study, 22 families that had at least five children baptized in the community church between 1850-1900 were identified with the help of genealogical records. All living descendants of these couples and their spouses were invited to take part in the study. Data collection started in June 2002 and was finished in February 2005.

##### 1.1.6 Rotterdam Study

The Rotterdam Study[15] is an ongoing prospective population-based cohort study, focused on chronic disabling conditions of the elderly[16] of which a random subset was exome sequenced. Participants were screened for dementia at baseline and at follow-up examinations using the Mini-Mental State Examination (MMSE) and the Geriatric Mental Schedule (GMS) organic level[17]. Screen-positives (MMSE <26 or GMS organic level >0) underwent extensive examination[18]. Finally, individuals were diagnosed in accordance with standard criteria for dementia (Diagnostic and Statistical Manual of Mental Disorders, Third Edition, Revised (DSM-III-R)) and Alzheimer's disease, NINCDS-ADRDA[3]. Follow-up for incident dementia was complete until January 1st, 2014. The Rotterdam Study has been approved by the Medical Ethics Committee of the Erasmus MC and by the Ministry of Health, Welfare and Sport of the Netherlands, implementing the Wet Bevolkingsonderzoek: ERGO (Population Studies Act: Rotterdam Study). All participants provided written informed consent to participate in the study and to obtain information from their treating physicians.

##### 1.1.7 ADC-Amsterdam

The ADC-Amsterdam cohort includes patients who visit the memory clinic of the Alzheimer Center at the Amsterdam University Medical Center, The Netherlands, and was described previously[19]. DNA samples from 854 patients with probable and possible AD were included in the current study. Additionally, 353 individuals diagnosed with psychiatric and subjective cognitive complaints were included as controls. Individuals in this cohort were extensively characterized to reduce the chance of misdiagnosis. Patients underwent an extensive standardized dementia assessment, including medical history, informant-based history, a physical examination, routine blood and CSF laboratory tests, neuropsychological testing, electroencephalogram (EEG) and MRI of the brain. The diagnosis of probable AD was based on the clinical criteria formulated by the National Institute of Neurological and Communicative Disorders and Stroke—Alzheimer's Disease and Related Disorders

Association (NINCDS-ADRDA) and based on National Institute of Aging–Alzheimer association (NIA-AA)[2]. Clinical diagnosis is made in consensus-based, multidisciplinary meetings. All patients gave informed consent for biobanking and for the use of their clinical data for research purposes. Selection for whole exome sequencing was based on an early age-of-onset (age at diagnosis <70 years) and available CSF biomarkers.

##### 1.1.8 Netherlands Brain Bank

From the Netherlands Brain Bank, we selected brain tissues donated by patients diagnosed with Alzheimer Disease. DNA was isolated and used for WES sequencing.

##### 1.1.9 UMC-Amsterdam

This cohort consists of WES data that were generated as part of a diagnostic work-up. All samples are from healthy adults for whom WES analysis was performed to aid the analysis of a patient, in most cases these were healthy parents of an affected child for whom trio-WES analysis was performed. These parents either have no pathogenic variant, or are carrier of one recessive pathogenic variant that does not affect health.

##### 1.1.10 100-plus Study

The 100-plus Study, is a prospective cohort study of cognitively healthy centenarians that associated with the Alzheimer Center at the Amsterdam University Medical Center. Detailed participant recruitment and procedures were described previously[20]. Trained researchers visited the centenarians at their home residence annually, where they were subjected to questionnaires regarding demographics, lifestyle, medical history, physical well-being and objective measurements of cognitive and physical functions. Cognitive function is tested by an extensive neuropsychological testing battery. For the current study, DNA samples 375 centenarians were included who completed at least one neuropsychological test at baseline, and exome sequencing from 349 centenarians passed QC (removal was mostly due to kinship). Centenarians who scored >22 on the MMSE were regarded as controls, while centenarians who scored ≤22 were regarded as cases[21]. The Medical Ethics Committee of the Amsterdam UMC approved this study and informed consent was obtained from all participants. The study has been conducted in accordance with the declaration of Helsinki.

##### 1.1.11 EMIF-AD 90-plus Study

The EMIF-AD 90+ study[22] is a cohort-study of the oldest-old (90+), situated at the Amsterdam UMC and the University of Manchester. The study contributed n=72 controls. Controls were tested to have a Mini-Mental State Examination (MMSE) >=26 and a global Clinical Dementia Rating (CDR) score of 0 at baseline.

##### 1.1.12 CBC: Control Brain Consortium

The Control Brain Consortium was previously described[23]. It consists of whole-exome sequencing in 478 samples derived from several brain banks in the United Kingdom and the United States of America. Samples were included when subjects were, at death, over 60 years of age, had no signs of neurological disease and were subjected to a neuropathological examination, which revealed no evidence of neurodegeneration. The data was made publicly available at www.alzforum.org/exomes/hex.

##### 1.1.13 PERADES

The PERADES sample (Defining Genetic, Polygenic and Environmental Risk for Alzheimer's Disease) comprises individuals with Alzheimer's disease (AD) and healthy controls recruited across UK, Italy and Spain. The majority of the individuals are from the UK (n=4095 with samples recruited in Cardiff: n=2405), while the rest (n=841) were recruited in Spain and Italy. More specifically the recruitment centres were: MRC Centre for Neuropsychiatric Genetics and Genomics, Cardiff University, Cardiff, UK; Institute of Psychiatry, London, UK; University of Cambridge, Cambridge, UK; University of Southampton, Southampton, UK; University of Nottingham, Nottingham, UK; Catholic University of Rome, Rome, Italy; Santa Lucia Foundation, Rome, Italy; Instituto di Neurologia Policlinico Universitario, Rome, Italy; University of Milan, Milan, Italy; Laboratory of Gene Therapy, San Giovanni Rotondo, Italy; University of Perugia, Perugia, Italy; University of Cantabria and IDIVAL, Santander, Spain and the Regional Neurogenetic Centre (CRN), ASP Catanzaro, Lamezia Terme, Italy. The collection of the samples within the MRC Centre for Neuropsychiatric Genetics and Genomics, Cardiff University was through national recruitment through multiple channels, including specialist NHS services and clinics, research registers and Join Dementia Research (JDR) platform. The participants were assessed at home or in research clinics along with an informant, usually a spouse, family member or close friend, who provided information about and on behalf of the individual with dementia. Established measures were used to ascertain the disease severity: Bristol activities of daily living (BADL), Clinical Dementia Rating scale (CDR), Neuropsychiatric Inventory (NPI) and Global Deterioration Scale (GlDS). Individuals with dementia completed the Addenbrooke's Cognitive Examination (ACE-r), Geriatric Depression Scale (GeDS) and National Adult Reading Test (NART) too. Control participants were recruited from GP surgeries and by means of self-referral (including existing studies and Joint Dementia Research platform). For all other recruitment, all AD cases met criteria for either probable (NINCDS-ADRDA, DSM-IV) or definite (CERAD) AD. All elderly controls were screened for dementia using the Mini Mental State Examination (MMSE) or ADAS-cog, were determined to be free from dementia at neuropathological examination or had a Braak score of 2.5 or lower. Control samples were chosen to match case samples for age, gender, ethnicity and country of origin. Informed consent was obtained for all study participants, and the relevant independent ethical committees approved study protocols. The whole exome sequencing (WES) was performed in-house at the MRC Centre for Neuropsychiatric Genetics and Genomics, Cardiff University. With the Nextera technology (Nextera Rapid Capture Exome v1.2), DNA was simultaneously fragmented and tagged with sequencing adapters in a single step. The enriched libraries were sequenced using the Illumina HiSeq 4000 (Illumina, USA) as paired-end 75 base reads according to manufacturer's protocols.

##### 1.1.14 StEP-AD

The overall goals of the Stanford Extreme Phenotypes in AD (StEP AD) project are to identify and characterize novel genetic variants that promote resilience to AD pathology in the presence of the APOE4 allele or that drive pathogenesis in the absence of the APOE4 allele. Genomes are collected from several sources, some intramural and some extramural. Invariably, the cognitive assessment protocols for these different sources vary somewhat but all include APOE genotyping, extensive neuropsychological testing, collection of one or more AD biomarkers, and consensus adjudication.

Genomes were sequenced for subjects in the following three categories: (1) Protected APOE4 carriers that have the APOE3/4 genotype, are at least 80 years old, and have normal cognition. If additional follow-up is expected we will accept subjects as young as 77; (2) Super-protected APOE4 carriers that have the APOE4/4 genotype, are at least 70 years old, and have normal cognition (if additional follow-up is expected subjects as young as 67 will be accepted); (3) APOE4-negative, early-onset cases that have the APOE2/2, 2/3, or 3/3 genotype and are diagnosed with probable AD before age 65. Most are also negative for known PSEN1, PSEN2 or APP mutations.

##### 1.1.15 Knight-ADRC

The samples Samples from the Charles F. and Joanne Knight Alzheimer’s Disease Research Center (Knight ADRC) were recruited at Washington University School of Medicine (WUSM) in Saint Louis, MO (USA). (REF). All the cases received a diagnosis of dementia of the Alzheimer's type (DAT), using criteria equivalent to the National Institute of Neurological and Communication Disorders and Stroke-Alzheimer's Disease and Related Disorders Association for probable AD[3,24]. Cognitively normal participants received the same assessment as the cases, and were deemed nondemented. Prior written consent, participants are genotyped for APOE4 allele and screened for known mutation in APP, PSEN1, PSEN2, MAPT, GRN, or C9orf72 by the Clinical and Genetics Core of the Knight ADRC. The approval number for the Knight ADRC Genetics Core family studies is 201104178.

##### 1.1.16 UCSF/NYGC/UAB

Studies in the UCSF/NYGC/UAB dataset were described previously[25]. Cases were selected from the University of California, San Francisco (UCSF) Memory and Aging Center with an intentional selection of early-onset cases to maximize the likelihood of identifying genetic contributors, along with healthy older adult controls (a total of 664 cases and 102 controls). All UCSF cases and controls were clinically assessed during an in-person visit to the UCSF Memory and Aging Center (MAC) that included a neurological exam, cognitive assessment, and medical history. Each participant's study partner (i.e., spouse or close friend) was also interviewed regarding functional abilities. A multidisciplinary team composed of a neurologist, neuropsychologist, and nurse then established clinical diagnoses for cases according to consensus criteria. This cohort was intentionally depleted of cases with known Mendelian variants associated with neurodegenerative diseases. A small number of samples (19 cases and 21 controls) were obtained from the University of Alabama at Birmingham (UAB) from an expert clinician who employed the same diagnostic procedures.

##### 1.1.17 UCL-DRC EOAD

University College London Dementia Research Centre (UCL-DRC) early-onset Alzheimer's disease cohort included patients seen at the Cognitive Disorders Clinics at The National Hospital for Neurology and Neurosurgery (Queen Square), or affiliated hospitals. Individuals were assessed clinically and diagnosed as having probable Alzheimer's disease based on contemporary clinical criteria in use at the time, including imaging and neuropsychological testing where appropriate. All individuals consented for genetic testing and had causative mutations for Alzheimer's disease (*PSEN1, PSEN2, APP*) and prion disease (*PRNP*) excluded prior to entry into this study.

##### 1.1.18 ADSP

ADSP Discovery phase (used in Stage-1): Cases and controls were selected from over 30,000 non-Hispanic Caucasian subjects from multiple cohorts described in detail elsewhere[26]. All controls were greater than 60 years and were cognitively normal based on direct assessment. All cases met NINCDS-ADRDA criteria for possible, probably, or definite Alzheimer's disease. All cases had a documented age-at-onset, and for those with pathologically conformed AD, an age-at death. APOE genotypes were available for all. Cases were selected to have a minimal AD risk based on sex, age and APOE genotype. Controls were selected as those with the least probability of converting to AD by age 85. Controls were older (86.1 years, SD = 5.2) than cases (76.0 years, SD = 9.2). The selection criteria and the rationale for study design are described elsewhere[27]. Eventually, 5,096 cases and 4,965 controls were selected for exome sequencing by this protocol, as well as 682 additional cases from multiplex families with a strong AD family history.

ADSP Discovery extension and Augmentation phase (used in Stage-2): Under funding provided by NHGRI, an additional 3,000 subjects were whole genome sequenced. This included 1,466 cases and 1,534 controls. Of these 1,000 each of Non-Hispanic White (NHW), Caribbean Hispanic (CH), and African American (AA) descent were sequenced. Of these a total of 739 autopsy samples were sequenced [568 cases (500 NHW cases and 68 AA cases) and 171 controls (164 NHW and 7 AA)]. The Case-Control and Enriched Case Study spans 24 cohorts provided by the Alzheimer's Disease Genetics Consortium (ADGC) and the Cohorts for Heart and Aging Research in Genomic Epidemiology (CHARGE) Consortium.
The Augmentation Phase encompasses sequencing done under private and NIH funding by investigators who are not members of the ADSP. The investigators for these studies have agreed to share their GWAS, WGS and WES data with the ADSP. Private funding has been provided by industry and anonymous donors. Under the NIA AD Genetics Sharing Policy and the NIAGADS Data Distribution Agreement, individual NIA funded investigators studying the genetics and the genomics of AD provide their data to NIAGADS.

Alzheimer's Disease Neuroimaging Initiative (ADNI) (used in Stage-2): A public-private partnership, the purpose of ADNI is to develop a multisite, longitudinal, prospective, naturalistic study of normal cognitive aging, mild cognitive impairment (MCI), and early Alzheimer's disease as a public domain research resource to facilitate the scientific evaluation of neuroimaging and other biomarkers for the onset and progression of MCI and Alzheimer's disease. In 2017, ADNI geneticists began collaborations with the ADSP. Whole genome sequence data on 809 ADNI subjects (cases, mild cognitive impairment, and controls) have been harmonized using the ADSP pipeline. Data used in the preparation of this article were obtained from the Alzheimer's Disease
Neuroimaging Initiative (ADNI) database (adni.loni.usc.edu). The ADNI was launched in 2003 as a public-private partnership, led by Principal Investigator Michael W. Weiner, MD. The primary goal of ADNI has been to test whether serial magnetic resonance imaging

(MRI), positron emission tomography (PET), other biological markers, and clinical and neuropsychological assessment can be combined to measure the progression of mild cognitive impairment (MCI) and early Alzheimer's disease (AD). For up-to-date information, see www.adni-info.org.

#### 1.2 Sequence read alignment and variant calling

We included raw sequencing data with three different types in our sample:
—**Exome sequences (ES):** reads cover the exonic regions of the genome according to a predefined 'capture kit'. Regions covered by capture kits differ according to kit-versions, or supplier.
—**Whole Genome Sequencing (WGS);** reads cover the whole genome.
**—Exome extracts:** reads that cover the target genes + 1000bp padding,

##### 1.2.1 Exome sequence read processing

Raw sequencing data from all studies were processed on a single site (Cartesius Supercomputer provided by SURF, in the Netherlands), and processed with a uniform pipeline. Reads were extracted from FastQ, BAM, CRAM or SRA files. For each lane/read group separately, paired reads were converted to SAM format using FastQToSam or picard RevertSam (Picard Tools version 2.10.5[28]), processed with Picard MarkIlluminaAdapters and subsequently transformed to interleaved fastq format with Picard SamToFastq (while setting marked adapter regions to base quality 2). Next, reads were aligned to the human reference genome (build 37 including unlocalized contigs and the Epstein-Barr virus sequence.) using the BWA MEM algorithm (BWA version 0.7.15-r1140)[29]. Alignments were processed with Samblaster (version 0.1.24) to add mate tags[30]. Read group alignments were then merged and duplicate reads were marked using Picard MarkDuplicates.

We found that the presence of novel Indels and novel SNPs in certain samples correlated with the presence of larger amounts of soft-clipped reads, indicative of the presence of chimeric DNA fragments. Each sample for which the percentage of soft-clipped base alignments exceeded 0.5% was therefore processed with a custom tool (see section 1.2.3) which identified and removed parts of reads that were likely of chimeric origin. This tool was executed after the Picard MarkDuplicates step. Then, reads were sorted to chromosome order by samtools sort (version 1.8)[31].

We estimated contamination percentages using VerifyBamID2[32], retrieved 4 September 2018), while correcting for the 2 PCA components (default), based on common SNPs (allele frequency ≥0.01) present in the 1000-genomes dataset (phase3, version 5b){The 1000 Genomes Project Consortium, 2015 #2305}. Base quality scores were recalibrated using GATK BQSR (version 3.8-1)[33] on the sample capture kit region + 100bp padding. Known indels were obtained from the Mills and 1000G gold standard indels in the GATK resource kit[33]. Known SNPs were obtained from dbSNP (version 150) and gnomAD (version 2.0.2)[34]. Subsequently, variants were called on the sample capture kit region + 100bp padding using HaplotypeCaller[33], while using the '-contamination' correction option, with the estimated contamination percentages. Ploidy was set to 1 for chromosome Y, and 2 for the other chromosomes, minPruning was set to 2, and the new quality model (--newqual) was used. Results were exported as gVCF format. Finally, gVCFs were combined per study in batches with a maximum size of 500 samples using GATK CombineGVCFs. Then, variants were called using GATK GenotypeGVCF[33], using the new quality model and setting max-alternate-alleles to 20. Variants were then annotated with GATK variant score recalibrator (VQSR) using allele specific annotations, while for all other options the best practices were followed.

##### 1.2.2 Processing of WGS reads and exome extracts

WGS samples were aligned according to the same pipeline and variants were called for the genomic region covered by the union of the exome capture kits. Then, genotypes were called using GATK GenotypeGVCF based on both exome and WGS gVCFs. Exome-extracts were also processed with the same pipeline. Resulting gVCFs were combined for the targeted regions with exome and WGS gVCFs (using GATK GenotypeGVCF). VQSR annotations were found to be less reliable if trained on a dataset that covered only the target genes, due to the relatively low number of variants. Therefore, VQSR variant annotations obtained on non-extracted samples (covering all genes and many variants) were transferred with priority to the dataset that contained also the extracted samples (covering only the target genes). Existing VQSR annotations were kept for variants unique to the exome-extract dataset.

##### 1.2.3 Chimeric read declipping

Chimeric fragments consist of multiple genomic sequences, joined together into one sequence. Sequencing of such fragments can result in reads that do not entirely align to the genome, and/or align at multiple locations. This results in so-called ‘soft-clipped’ alignments, where parts of the read sequence are not aligned. These soft-clipped regions cause issues for the variant caller, as it uses not just the aligned part of the reads, but also the unaligned soft-clipped regions during local reassembly and variant calling. The reason for this is that these clipped sequences can be an indication of an insertion variant. In case these clipped regions are caused by chimera’s, this is however not a correct strategy, and can cause false variant calls. To prevent their effect on variant calling, we i) estimate the extent of the chimera problem by quantifying the number of soft-clipped alignments, and ii) remove these soft-clipped sections for affected samples if they are (likely) caused by chimeras. To do this, the soft-clipped sections are turned into hard-clipped alignments, in which the underlying sequence is removed (the read is shortened), such that the variant caller cannot revive the clipped sequence during variant calling. In the following description, we assume paired end sequencing (in which both ends of the fragment are sequenced, resulting in two reads). We remove the following soft-clipped sequences:

i) One well-known type of artificial chimera occurs when the sequenced fragment is shorter than the read length. Fragments have adapters at the end, used as starting point for sequencing. In these cases, the 3’ end of read 1 will cover the adapter of read 2, and vice versa. Due to this, read 1 and 2 will have overlapping alignments with possibly soft-clipped 3’ends. Such read pairs can be detected based on their overlapping alignments. To remove the adapter sequence, we align the known adapter sequence to determine the clipping point, and hard-clip the identified sequences from there.

ii) A genomic chimera can have a join-point at different sites in the sequence fragment.
—If the chimeric join point occurs between read 1 and 2, or close to the end of read 1 or 2, then read 1 and 2 will (usually) be aligned at a distance from each other. If this distance

is >100kb, or one of the reads is unmapped, we remove the soft-clipped regions at the 3’end of both reads.

—If there are multiple, mostly non-overlapping, alignments for a read at different genomic locations, it is usually an indication that the chimeric join point occurs somewhere in the middle of that read. The overlapping parts of these alignments are pruned (in all alignments for that read). Then, soft-clipped sequences in the alignments that face each other are hard-clipped.

In the above situation, it frequently occurs also that the fragment is short. The chimeric join point might then be present in both reads. If both reads have multiple alignments, we handle each read as described above.

—If the fragment is short, but not very short, read 1 might have multiple alignments, while read 2 has a soft-clipped 3’end (or vice versa). For example, for genomic region A and B, a chimeric fragment might read AABBB. Read 1 (AABB) might then have multiple alignments, one for the AA and one for the BB section. Read 2 (BBBA) however might have only an alignment for B, but not for A, as the sequence from A is too short to obtain an accurate alignment. The chimeric sequence A in read 2 will therefore be soft-clipped. We detect these situations based on overlapping alignments for fragment B, and hard-clip the soft-clipped 3’end of read 2.

—If the chimeric sequence consists of a very short piece at the 5’ end of either read 1 or 2, this part might not be aligned as it is too short. It is in these situations unclear if the sequence has a chimeric origin, as such unaligned pieces can also be caused by indels. We find that in samples affected by chimeras, it is beneficial to remove these soft-clipped 5’ends. While this reduces the coverage of indels, in most cases many fragments still cover the complete indel. Also, differences in coverage between samples occurs commonly in exomes, where the covered regions are highly variable between capture kits, and handling this is part of the downstream pipeline (see posterior probabilities).

—After alignment pruning and removal of the soft-clipped regions caused by chimera’s, we unalign the alignments that are <= 1bp in length, we transform supplementary alignments to primary alignments if the primary alignment is unaligned, drop unaligned supplementary alignments, update alignment tags, and validate the read records and cigar strings.

#### 1.3 Sample QC

Results of the sample QC steps are shown in **Table S3a**.

Before sample QC, we performed a pre-variant QC step, to remove bad quality variants (see Variant QC steps for details) that might impact sample quality statistics. In addition, we required that variants cover at least 25% of the samples with at least read depth 6. Next, sample QC was performed.

##### 1.3.1 Missingness

We removed samples that had a GQ<20 for 40% of the variants in its own exome kit, or a depth < 6 for 35% of the variants in its own exome kit. Additionally, we removed samples for which chromosomes were missing (GQ < 20 for 99% of the variants on a chromosome in the samples exome kit).

##### 1.3.2 Contamination

Samples with a contamination percentage > 7.5% were removed.

##### 1.3.3 Sex-check

We performed a sex-check, by comparing annotated sex with genetic sex (**Figure S5**). Genetic sex was determined based on the coverage of the sex chromosomes. Coverage was determined using off-target reads. Only coverage in regions outside capture kits (+500 bp padding), outside peaks in coverage called with MACS (version 1.4)[35] and outside segmental duplications (Segmental Dups track downloaded from UCSC which includes the PAR regions[36]). Coverage was determined in 20kb windows, and normalized for GC content using linear regression. Regions of 20kb with more than 100 N bases were discarded. X and Y chromosome coverage was normalized by dividing by the autosome coverage. Thresholds were set empirically, based on the distribution of male and female samples (see supplemental **Figure S5**).

##### 1.3.4 Population outliers

Next, we performed a PCA analysis to identify population outliers. Variants that were in the intersection region of all capture kits, and had a minor allele frequency ≥0.005 and a depth ≥6 for 90% of the samples, were used for this purpose. Variants were pruned with bcftools +prune tool (version 1.8)[31] with max LD set to 0.2 in 500kb windows. Only variants that were also in the 1000 genomes dataset (phase 3, v5b) were kept. PCA was performed on dosages (based on genotype calls for 1000G, and based on genotype probabilities for the study samples). Variant dosages were first normalized, as described[37], based on statistics obtained on the 1000G samples. Then, PCA was performed on the 1000G samples, and all ADES samples were mapped to this PCA space (**Figure S6**). Finally, we removed outliers for each of the first 4 PCA components (**Figure S7, Figure S8**), where outliers were defined as samples that fell outside the range *median(pca_component) ± 8 * mad(pca_component)*, where *mad* is the median absolute deviation and the *pca_component* vector only contains the ADES samples.

For the Stage-2 dataset, which contains a large fraction of non-European individuals, an outlier approach was not sufficient. Therefore, a k-nearest neighbor classifier (SKLearn v0.20.3, k=10) was trained on the first 10 PCA components, using the 1000G samples, to predict their ancestry (distinguishing Africans, Europeans, Admixed Americans, East Asicans, and South Asians). This predictor was applied to all samples in the Stage-2 data, and samples predicted to be non-European were removed. Subsequently, we continued with the outlier approach already described.

##### 1.3.5 Excess novel SNPs

##### 1.3.6 Excess novel indels

We calculated and compared the number of novel SNPs and the number of novel indels per study, both in the union of the capture kits (**Figure S9 and Figure S10)** and the intersection of the capture kits (**Figure S11, Figure S12)**. Novel variants were defined as variants that were not present in DBSNP v150. These statistics were calculated based on posterior dosages (described below). Thresholds were set at the *median value + 6 * mad* for novel SNPs and *+12*mad* for novel Indels.

##### 1.3.7 Heterozygous/homozygous (Het/Hom) and transition/transversion ratios (Ts/Tv)

Furthermore, we performed a per-sample QC on the following statistics (calculated on the intersection of the capture kits): Ts/Tv ratio of known SNPs (**Figure S13**), and Ts/Tv ratio of novel SNPs (**Figure S14),** Het/Hom rate of known SNPs (**Figure S15**). Known SNPs are those that are present in dbSNP v150, while other SNPs are considered novel. The acceptable range for Het/Hom was set to ±6 * mad. For Ts/tv measures, only a lower limit of -6 * mad was used.

##### 1.3.8 Identity By Descent (IBD) analysis

We performed an IBD analysis on the remaining samples using Seekin[38]. We kept variants with a minor allele frequency ≥0.005, and for which at least 90% of the samples had depth >=6. Variants were pruned with bcftools +prune tool (version 1.8), with max LD set to 0.2 in 500kb windows. Only variants that were also in the 1000G dataset were kept. We performed a PCA as described before. Using Seekin (version 1.0), we corrected for these PCA components using the options 'modelAF' and 'getAF', using 4 PCA components. Next, kinship was determined using all variants with the heterogeneous estimator of Seekin[38]. Duplicate samples with inconsistent annotation were removed (inconsistent status, *APOE* genotype, or gender, or more than 2 years difference in age at onset for cases). Otherwise, we kept the sample with the most complete annotation: we preferred samples with age (at onset), and *APOE* genotype over samples without. Also, we preferred whole genome sequenced samples over exomes, and samples with lower missingness over samples with higher missingness. For related samples up to 3$^{rd}$ degree (marked by the threshold of >9.375% shared identity by descent, which is the middle value between the expected value for 3$^{rd}$-degree (12.5%) and 4$^{th}$-degree (6.25%)), we preferred (in order) cases over controls, samples with more clinical data (age (at onset), apoe status), WGS samples, and samples with higher coverage.

Additionally, Stage-1 and 2 samples were processed together, to detect samples that were duplicated between Stage-1 and 2. These samples were removed from Stage-2.

##### 1.3.9 Bad PCR plates

We removed all samples on 3 PCR plates that were enriched with gender mismatches.

##### 1.3.10 Removal of dementia-related (likely) pathogenic variant-carriers

Next, on the Stage-1 set, we performed a manual curation of causative variants in a short list of Mendelian dementia genes. We extracted rare variants in the following two gene lists and interpreted them following the American College of Medical Genetics and Genomics and the Association for medical Pathology[39], (i) autosomal dominant AD genes: *APP*, *PSEN1*, *PSEN2* (autosomal dominant AD), *GRN*, *MAPT*, *FUS*, *TARDBP*, *VCP*, (fronto-temporal lobar degeneration spectrum), *NOTCH3* (CADASIL), *PRNP* (Prion diseases); (ii) autosomal recessive genes: *NPC1*, *NPC2* (Niemann-Pick type C disease), *TYROBP*, *TREM2* (*homozygous LOF: Nasu-Hakola disease, 1 carrier)*). Carriers of variants that reached enough evidence to be rated at least as likely pathogenic (class 4) were excluded from the analysis, whatever their disease status. Of note, for autosomal recessive genes, heterozygous carriers were not excluded, only carriers of bi-allelic pathogenic variants were excluded.

##### 1.3.11 AD label

We excluded samples for which clinical information was indicative of non-AD dementia (e.g. vascular dementia). In addition, part of the case-control samples included minimal neuropathological information. Among them, we further excluded samples with discordant Braak stages, i.e. cases with stage <2 (n=265) and controls with stage >4 (n=43).

##### 1.3.12 Handling of exome-extract samples

Part of the Stage-2 dataset consists of exome-extracts, which only cover the targeted genes with 1000bp padding. For these samples, we relied on the study QC. Separate checks were performed for missingness (no outliers), contamination (1 outlier), and population (no outliers).

###### 1.3.13 Merging of Stage-1 and -2 sample QC

For the mega-analysis sample, Stage-1 and 2 QC were merged, while adding a separate IBD step to additionally remove <= 3rd degree family relations as described above that remained between samples in Stage-1 and -2.

#### 1.4 Variant QC

Throughout an extensive QC, we attempted to find root causes for the presence of false variants. We identified two significant issues that were not handled by the default variant calling pipeline: false positive variants due to (soft-clipped) chimeric alignments and oxygenation of G bases.

After removal of samples excluded by the sample QC, variant statistics were recalculated. Then, we performed variant QC as described in **Table S3b (non-exome-extract samples)** and **Table S3c (all samples, only targeted regions)**.

##### 1.4.1 Multi allelic variants

First, multi-allelic variants were split into bi-allelic variants, and indels were normalized, using the bcftools norm tool. The tool was modified to also split the phased PGT fields, such that downstream variant merging was possible. Additionally, the splitting of the genotype likelihoods and read counts was modified (PL and AD fields), which is detailed in the next section. We removed bi-allelic variants that had as alternate allele '*' (which reflects overlap with a deletion variant), as well as multi-allelic variants for which the reference allele was lower in frequency than the frequency for at least two alternate alleles.

##### 1.4.2 Variant merging

Variants that were in close vicinity, in cis and always occurred together, were merged into single events, to account for for example nearby frameshifts that cancel each other out. Only indels with ≤10bp distance and SNPs with ≤2bp distance were considered for merging. We used the read-phasing output of GATK (PID/PGT fields) to determine which variants occurred in-phase.

##### 1.4.3 Oxo-G

In some samples novel variants were enriched for G>T and C>A variants, caused by the oxygenation of G bases during sample processing[40]. Using a custom tool (see below), that uses per-sample statistics from Picard CollectSequencingArtifactMetrics, we identified and filtered variants and variant calls that could be attributed to this issue. We removed variants with an average OXO sensitivity > 1.5, or a remaining total dosage after OXO correction ≤0.1.

##### 1.4.4 Short Tandem Repeat (STR) and Low Copy Repeats (LCR) regions

STR and LCR regions were obtained respectively from the simple tandem repeats track by TRF from UCSC, and the LCRs as identified by the mdust program[41]. Variants in these regions were excluded.

##### 1.4.5 Allele Balance

The balance between reference and alternate reads (allele balance) was determined both for heterozygous and homozygous calls. Allele balance was calculated based on posterior genotype probabilities (see below). Variants that had an average allele balance < 0.25 or > 0.75 for heterozygous calls, or < 0.9 for homozygous calls were removed.

##### 1.4.6 Depth Fraction

The relative depth of heterozygous calls to other calls was determine, based on posterior genotype probabilities (see below). Variants for which the heterozygous depth was < 20% of the depth of other calls were removed.

##### 1.4.7 Hardy Weinberg

Hardy-Weinberg scores (all samples and control samples: hw_all and hw_control) were calculated based on posterior genotype probabilities (see below). We removed variants for which the p-value for control samples was $< 5 * 10^{-8}$.

##### 1.4.8 VQSR

Variants that were tagged by the variant quality score recalibration method from GATK were removed. For Stage-1, for SNPs we removed variants from the VQSR > 99.5% sensitivity tranche, while for indels we removed variants from the VQSR > 99.0% sensitivity tranche. For the Stage-2 and mega datasets, these sensitivity thresholds were too low, possibly due to higher quality input and/or more included samples. This resulted in a larger fraction of removed variants, with higher ts/tv values than obtained in Stage-1. We therefore conservatively set the threshold to 99.8% for SNPs and 99.5% for indels, to attain similar removal rates of variants for Stage-1 (2.1%) and 2 (1.8%), and the mega analysis (2.0%) (**Table S3b,c**).

##### 1.4.9 Pre-variant QC versus final variant QC

For the pre-variant QC, which is performed prior to performing the sample QC, we performed all the above steps. Additionally, we removed variants with a missingness rate > 25%. Genotype calls which had a depth < 6 were considered missing. For the final variant QC, the missingness step was not performed, as it is included as part of the variant selection. Compared to the pre-variant QC, the final variant QC had variant batch detection as an additional step.

##### 1.4.10 Variant Batch Detection

Finally, a custom tool was developed to remove variants that still presented batch effects that were not explainable by population structure or phenotype effects (see below). On variants identified to have a batch effect, we performed variant batch correction, by setting batches that caused problems for a certain variant to missing. Afterwards, variants that still had a Variant Batch Detector (VBD) score > 25, or a VBD score > 15 and MAF < 0.005, were removed from the analysis.

#### 1.5 Genotype posterior probabilities

Due to the use of different capture kits and whole genome sequencing (WGS) data, the analysed dataset has highly variable coverage patterns across the samples. Many variants have as a consequence less than 100% coverage across the samples. In burden testing, a missingness percentage of up to 20% is allowed. This requires an accurate handling of missing genotype calls in variants that contribute to the burden score. In cases of low and absent read coverage, direct calling of the genotype is not possible. Therefore instead, a probabilistic approach is used, in which each genotype is assigned a certain probability.

##### 1.5.1 Genotype likelihoods

The GATK variant caller outputs the likelihood of each sample genotype in the PL field of the VCF. These likelihoods are based on the available sequencing reads for a sample. In case of missing data, each genotype is considered equally likely (i.e. p=⅓ in case of diploid chromosomes for ref/ref, ref/alt and alt/alt genotypes). These likelihoods cannot be used directly in a burden analysis, as by assuming equal likelihoods for each genotype the allele frequency of samples with missing coverage would effectively by 50%, and likely substantially differ from that of samples with coverage.

##### 1.5.2 Posterior probability

This is solved by the use of posterior probabilities. Here the allele frequency in the study sample is used as a prior in assigning genotype probabilities. Using Bayes theorem, posterior genotype probabilities take the following form (assuming a diploid setting): $P(g) = \frac{L(g)\psi(g)}{\sum_i^G L(i)*\psi(i)}$, where *P(g)* is the posterior probability for genotype *g*, with *g* encoded as 0,1 or 2 for respectively the reference, heterozygous and homozygous alternate genotype. *L(g)* is the genotype likelihood as given by the variant caller. The genotype frequency $\psi(g) = \frac{2}{(2-g)!g!}\omega^g(1-\omega)^{2-g}$ is derived from the alternate allele frequency $\omega$, assuming Hardy-Weinberg equilibrium. Notably, the allele frequency $\omega$ needs to be derived from the study sample, such that $\omega$ matches the allele frequency in samples with coverage, thereby preventing biases. A difficulty is that accurate estimation of this allele frequency requires posterior genotype probabilities. Here we follow the approach previously described by Li et al[42] using an EM-algorithm in which iteratively posterior probabilities and the allele frequency are estimated, until convergence (maximum difference in allele frequency between iterations is $1e^{-7}$) is reached. Finally, posterior dosages in the diploid case were calculated as $d = P(1) + 2\,P(2)$.

##### 1.5.3 Multi-allelic variants

As described in the previous section, variants with multiple alleles are split into bi-allelic variants prior to analysis. For this, the bcftools norm tool is used. However, splitting of the genotype likelihood was adapted from the default approach in bcftools. The standard REF/ALT interpretation of the resulting biallelic likelihoods was considered problematic for the analysis, as often the alleles would be neither REF nor ALT. Genotype probabilities would then not sum to 1. We adapted therefore to a NON_ALT/ALT interpretation of bi-allelic variants. Specifically, this meant that genotype likelihoods were converted to probabilities, and then summed to obtain the NON_ALT/NON_ALT, NON_ALT/ALT and ALT/ALT genotype probabilities (separately for each ALT in the multi-allelic variant to create multiple bi-allelic variants).

Notably, in the absence of coverage, the variant caller considers each multi-allelic genotype equally likely. In this situation, the NON_ALT/NON_ALT genotype becomes the most likely genotype, as it sums more genotypes. As this causes biases, we correct for this, using an additional correction factor equal to *1 / (#summed multi-allelic genotypes)* for each bi-allelic genotype. Next to the genotype likelihood, the read count field (AD field) was also modified to follow the above described NON_ALT/ALT interpretation. To that end, read counts that contributed to the NON_ALT/NON_ALT and NON_ALT/ALT genotypes were summed during variant splitting.

##### 1.5.4 Posterior sample QC-measures

Standard sample QC measures, when calculated on variant calls, are affected by samples with low or missing coverage. To prevent that, these measures were instead based on genotype posterior probabilities:

***Nr. Of indels/SNPs:*** Determined by summing (across all samples) posterior dosages.

***Ts/Tv ratio:*** Determined by summing posterior dosages of transition variants and dividing them by the summed posterior dosages of transversion variants

***Het/Hom ratio:*** Determined by summing (across all samples) the posterior genotype probability of the heterozygous genotype, and dividing it by the summed posterior genotype probability of the homozygous genotype.

Posterior variant QC-measures

***Heterozygous allele balance:*** Defined as $\frac{\sum_i^N P_i(1) r_{ref}}{\sum_i^N P_i(1)(r_{ref}+r_{alt})}$, where *P*$_i$*(1)* is the posterior genotype probability for the heterozygous genotype for sample *I*, *N* is the number of samples, and $r_{ref}$ and $r_{alt}$ are the number of reads carrying the reference or alternate genotype.

***Homozygous allele balance:*** Defined as $\frac{\sum_i^N P_i(2) r_{alt}}{\sum_i^N P_i(2)(r_{ref}+r_{alt})}$, where *P*$_i$*(2)* is the posterior genotype probability of the homozygous genotype for sample *i*.

***Heterozygous depth ratio:*** Defined as $\frac{\frac{\sum_i^N P_i(1)(r_{ref}+r_{alt})}{\sum_i^N P_i(1)}}{(r_{ref}+r_{alt})/N}$.

***Hardy-Weinberg equilibrium:*** Posterior genotype probabilities assume Hardy-Weinberg equilibrium (HWE), thereby biasing variants with high rates of missingness towards HWE. Hardy-Weinberg equilibrium is therefore tested on non-probabilistic genotype calls, after filtering out samples with a read coverage < 6.

#### 1.6 Oxo-G variant call filtering

During sample preparation, oxidation of G-nucleotides can lead to the generation of 8-oxoguanine lesions in DNA. These lesions lead to false positive G-T variants, and, dependent on the protocol step in which the oxidation occurs, also false positive C-A variants[40]. While this is primarily an issue for somatic variant calling, it also impacts germline rare-variant calls, in particular in exomes where coverage is variable. In modern protocols, these effects have mostly been mitigated, however, in older samples these false positive mutations can be a significant source of errors. Next to oxoG errors, similar problems are known to occur in DNA obtained from formalin-fixed samples. In these samples, deamination can occur, converting cytosine to uracil (C>U), thereby creating false positive C->T (and G->A) mutations. While the approach below handles these types of errors as well, this problem was not encountered in a significant manner in the dataset.

A modern variant caller such as GATK determines nucleotide-specific base error rates based on a comparison of the sequenced reads to the genome (in the case of GATK through base quality score recalibration (BQSR)). In GATK, this error rate is modelled on the observed nucleotide in the read (e.g. in case of a G->T mutation a T for reads aligned to the positive strand and an A for reads aligned on the negative strand). Although G-oxidation will lead to a somewhat higher base error rates in T and A nucleotides, the variant caller does not recognize that these errors occur mainly when the genomic reference contains respectively a G (or C in case of C->A mutations). This leads to underestimated error rates and, in the end, false positive variant calls. Briefly, our approach to detect and filter these oxo-G affected variant calls is therefore based on comparing i) the dosage as determined when considering a error model that does not consider oxoG errors ii) the dosage as determined with a model that does consider (sample-specific) oxoG errors. The ratio of these two dosages is considered a 'sensitivity' score, which is used to filter genotype calls and/or variants. Dosages are computed using a genotype likelihood calculation detailed below, and are 'posterior dosages' (see previous section): continuous numbers between 0 and 2, which take into account the confidence in the genotypes and the frequency of the variant in the study sample. In the variant QC pipeline, genotype calls with a sensitivity > 1.5 are set to missing, after which variant QC statistics are recalculated. Variants are flagged for exclusion if they have an average sensitivity > 1.5 or a summed dosage with the oxo-G error model < 0.1. The average sensitivity of a variant is here defined as the ratio of the summed normal dosages and the summed oxo-G-corrected dosages. In more detail, the method consists of the following steps:

##### 1.6.1 Oxo-G statistics

To determine the parameters for the base error model, we estimate for each sample the rate at which oxidation and other base errors occur, dependent also on different sequence contexts (neighboring bases affect the G-oxidation rates). These per-sample statistics are collected using Picard CollectSequencingArtifactMetrics. Next to base errors, we also obtain summary error metrics per sample, based on measures available as part of the CollectSequencingArtifactMetrics. These consider two forms of the oxoG errors: pre-adapter (in this case G->T errors occur in forward reads, and C->A errors in backward reads) and bait-bias (in this case G->T errors occur in the exome template strand (often the positive strand), while C-A errors occur in the reverse strand).

##### 1.6.2 Full error model

The error model describes mutation-specific error rates (in contrast to the usual read-nucleotide specific error rates). It takes into account sequence context (a single nucleotide before and after the variant). Strand-specific and forward/backward read specific error rates are averaged: although this information would be useful, it is not available per sample in the variant file (VCF), and a direct link between the original reads in the bam file and the read count in the VCF file is not straightforward to make due to the reassembly step performed by the variant caller.

##### 1.6.3 Contrasting error model

A contrasting error model is created which exclusively models non-oxoG related errors. To this end, we select samples that are not affected by oxoG-related issues, based on the previously described summary metrics. As these summary metrics are sequence-context specific, we obtain a worst-case summary metric per sample, by taking the highest error value across all sequence contexts per sample. Samples with an error rate > 0.0001 for either pre-adapter or bait-bias errors are excluded. Using the remaining samples, regression models are trained which predicts (sequence context-specific) G->T and C->A mutation rates. These regression models are used to fill in G->T and C->A mutation rates for the samples that were excluded due to oxoG effects. Features for these regression models are the (sequence-context-specific) mutation rates for all mutations except G->T and C->A. To handle the extensive collinearity in these features, we reduce the feature space to 10 dimensions by using PCA, and make use of ridge regression.

##### 1.6.4 Genotype likelihood calculation

For each sample, genotype likelihoods are calculated both using the contrasting and full error model. Read counts ($r_{ref}$ and $r_{alt}$ for respectively reads carrying the reference and the alternate allele) are obtained from the VCF file. Based on the error model, sequence context, and reference and alternate allele, ref->alt ($e_{ra}$) and alt->ref ($e_{ar}$) error rates are obtained. For a sample *s* (identifier omitted for brevity), and assuming a diploid setting, the likelihood of each genotype is calculated then as:

ref/ref: $(1 - e_{ra})^{r_{ref}} e_{ra}{}^{r_{alt}}$

ref/alt: $\left(\frac{(1-e_{ra})+e_{ar}}{2}\right)^{r_{ref}} \left(\frac{(1-e_{ar})+e_{ra}}{2}\right)^{r_{alt}}$

alt/alt: $(1 - e_{ar})^{r_{alt}} e_{ar}{}^{r_{ref}}$

Likelihoods are normalized to sum to 1, and then converted to posterior probabilities ($p_{ref/ref}$,$p_{ref/alt}$ and $p_{alt/alt}$) as outlined in the previous section. The dosage per sample is then calculated as $d_s = p_{ref/alt,s} + 2p_{alt/alt,s}$ (where *s* refers to a specific sample) while sensitivity per sample is determined as: $s_s = d_{contrasting,s}/d_{full,s}$. Here, *full* and *contrasting* refer to the used error model to calculate the dosage. In practical use, we found that estimated oxoG-related errors are underestimated. This can be attributed to two factors: i) information loss as no information on read strand, and presence of mutations on forward and backward reads could be used. This could have diluted the estimated oxoG related-errors by a factor 2, ii) a selection bias, as false positive variants caused by this issue are likely sites that present more extreme oxoG-related errors, either by chance or due to (possibly unmodelled) sequence characteristics. To alleviate this issue, an error multiplication factor *f* was introduced, such that errors considered in the full model are rescaled according to $f(e_{full} + e_{contrasting}) + e_{contrasting}$. In practice, using $f = 5$ led to an adequate filtering of oxoG related variants.

##### 1.6.5 Genotype and variant filtering

Next to a genotype sensitivity measure, we also calculate a variant sensitivity measure: $s_{variant} = \frac{\sum_{samples} d_{contrasing,sample}}{\sum_{samples} d_{full,sample}}$. Variants were exluded from the analysis if $s_{variant} > 1.5$, or if $\sum_{samples} d_{full,sample} < 0.1$. For variants with $s_{variant} > 1.1$ we performed genotype filtering, setting to missing all genotypes where the genotype sensitivity $s_s > 1.5$. Afterwards, variant QC measures (missingness, Hardy-Weinberg, allele balance, etc) are recalculated.

#### 1.7 PCA covariates

For both variant QC and burden testing, PCA population covariates were estimated. These were calculated after sample QC, using an approach described previously[32]. Variants that were in the intersection region of all capture kits, and had a minor allele frequency ≥0.005 and a depth ≥6 for 90% of the samples, were used for this purpose. Variants were then pruned with bcftools +prune tool (version 1.8)[31] with max LD set to 0.2 in 500kb windows. PCA was performed on dosages (based on genotype probabilities). Variant dosages were first normalized, as described[37], after which PCA was performed.

To estimate PCA covariates for the Stage-2 exome-extract samples in concordance with the covariates estimated for the non-extract samples, we selected the variants from their target areas that also occurred in the set of variants used for determining the PCA in the non-extract samples. Then, using the non-extract samples, a linear regression model was learned to map these common variants to PCA covariate values, for each covariate separately. Spearman rank correlations between predictions and actual values were significant up till PCA component 7. Correlations were 0.7, 0.47, 0.35, 0.23, 0.49, 0.12 and 0.06 for the first 7 PCA components. PCA distributions for extract samples looked similar to those obtained for non-extract samples (**Figure S16-18**).

#### 1.8 Variant batch detection and correction

For genetic studies, statistical power is a primary concern. This necessitates large-scale collaborations between sites, as well as the collection of samples that have been sequenced across a large time period. In such settings, it is often impossible to control which capture kits are used, if exome or WGS sequencing is performed, and many other relevant sequencing parameters such as read or fragment lengths. In the ADES consortium, this has resulted in the use of 18 different (versions of) capture kits, the use of both exome and WGS sequencing, read lengths that vary from 50 to 150 bp (**Figure S3**), and many other differences. Moreover, the different contributing studies also have very different case/control balances, ranging from exclusively cases to almost exclusively controls. When performing variant association, this presents a problem, as this step is highly sensitive to batch effects. Even after sample and variant QC, we found that certain variants still present batch effects that lead to spurious associations.

##### 1.8.1 Examples of batch effects

It is not always immediately clear what the cause of such remaining batch effects is. Some examples which were encountered:
—Certain capture kit methods use restriction enzymes to cut sequence fragments before sequencing. We observe that mutations in these restriction sites can at some loci lead to an artificial loss of heterozygosity in the sequencing reads, resulting in a lower-than-expected allele frequency. Additionally, it is not possible to filter out PCR duplicates for these kits, leading to possible false positive mutations.
—For capture kits that fragment DNA at relatively 'fixed' positions in the genome we also observe an increase in batch effects. Explanations for this might include position-related biases in reads or mutations that affect the read coverage of one haplotype. This is observed for capture kits that use restriction enzymes for fragmentation, but to a lesser extent also for those that use transposases, which can have tagmentation biases[43]. Finally, such batch effects are also present in probe-based kits for variants that in terms of read length are distant from a capture probe.
—Increased batch effects are also observed in WGS samples when compared to exome samples. A possible explanation might be that WGS samples have sequence reads originating from the whole genome, in contrast to exome capture kits. In some cases, this could result in sequences being misaligned at certain locations that are not present when using (certain) exome capture kits.
While not every batch effect can be easily be predicted based on causal mechanisms, the presence of many different batches in the dataset still enables the detection of the variants with problematic batch effects.

##### 1.8.2 Algorithm overview

To this end, we developed a method to detect variants that are affected by such batch effects. The main challenge is to distinguish between non-technical effects that present as batch effects (such as a variant that is enriched in a certain country, and/or only in AD cases) and real batch effects that are caused by technical issues. This is solved by using a two-step approach. In the first step, the algorithm attempts to explain the presence of a variant in specific carriers only through population structure, presence of haploblocks, and/or phenotype effects. Secondly, it is determined if the explanation for the presence of a variant in specific carriers significantly improves if also technical covariates (membership of study batches, various sequencing parameters, etc.) are allowed. Variants for which this is the case are considered to be affected by technical issues, and are either corrected (detailed below) or not considered in the analysis. Below, we first detail the covariates that are used, the algorithm that is used to select the covariates, the regression model, how the presence of not-at-random missing genotypes (i.e. missingness depends on having a specific genotype) is detected, and finally how the algorithm is used in practice.

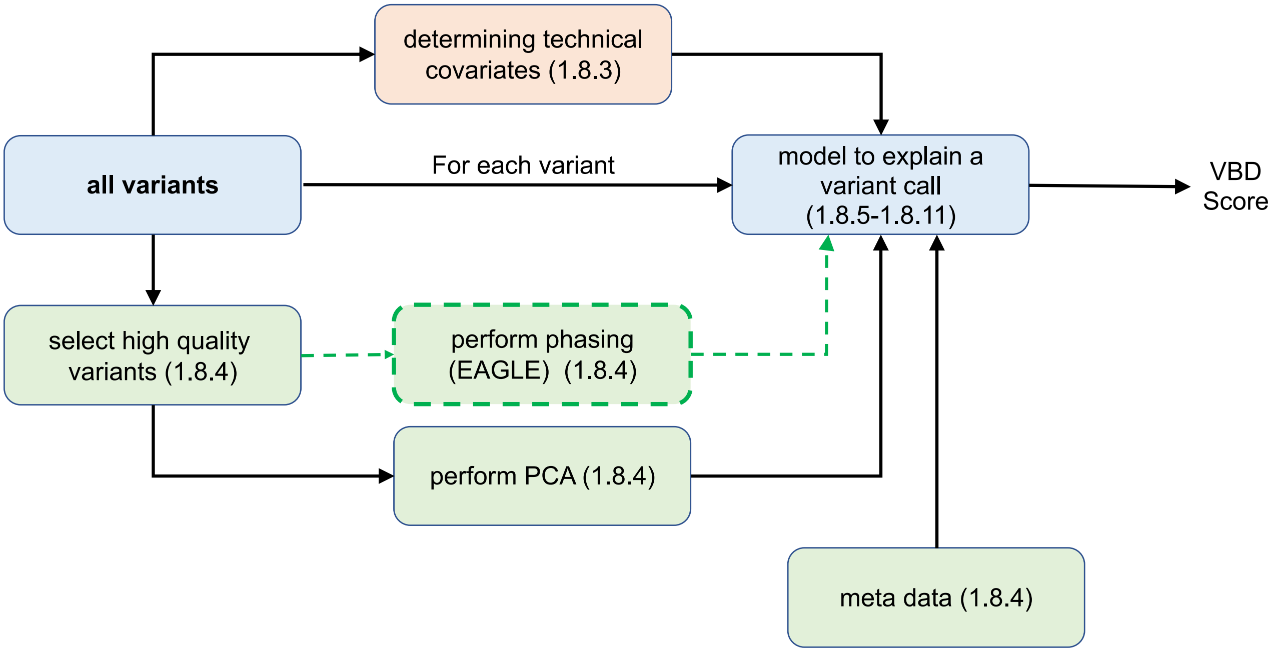

**Overview of batch effect detection strategy.** Each step of the batch effect detection is represented, and further explained below in the sections indicated in the boxes

##### 1.8.3 Technical covariates

Statistics were generated with SAMtools[31], Picard[28], verifybamid2[32], and custom scripts. Covariates (which are vectors that contain for each sample a value) were defined for the following properties:

- **Batch, study, capture kit:** Covariates describing (for each sample) membership (no: 0, yes: 1) for each batch, study or (version of a) capture kit.
- **Read length, insert size:** Covariates describing read length and average fragment insert size. In addition, covariates were added describing the distance to the nearest capture probe (which differs across the samples due to the use of different kits), both in absolute terms, as well as relative to fragment size or read length (**Figure S3**). For WGS samples, 0 was used as the distance.
- **Contamination:** Contamination percentage as determined by Verifybamid2 (see sample QC)
- **Missingness:** Sample missingness (defined as genotype quality GQ < 40, for variants that are in the intersection of all capture kits, **Figure S4**.
- **Size selection:** The standard deviation of fragment insert-sizes divided by the average of fragment insert sizes. Indicative of the extent of size selection that was performed on the fragments.
- **Read error rate:** Error rate of the reads (mismatches / bases mapped).
- **GC ratio:** Depth of sequences with 35% GC / Depth of sequences with 50% GC
- **Mismapping ratio:** Fraction of fragments for which the two reads map to different chromosomes
- **Duplicate ratio:** Fraction of duplicated reads.
- **Not mapped ratio:** Fraction of reads that are not mapped.
- **Read quality variability:** Standard deviation of average Illumina quality scores across read cycles (a cycle corresponds to a single base position in each read).
- **Fraction of N nucleotides:** Percentage of bases being the N (unknown) nucleotide.
- **Insertion/deletion error fraction:** Nr. of insertions or deletions divided by the nr. of bases mapped.
- **Ts/tv rate, Het/Hom rate, Novel SNPs/Indels rate:** Sample statistics as defined in the sample QC.

— **Gender:** Genetic sex (**Figure S5**).

— **Supplementary reads / fraction of soft-clipped bases:** Fraction of reads with supplementary alignments, and fraction of mapped bases that are soft-clipped.

— **Pre-adapter/Bait oxo-G error pattern:** Phred-scaled error indicating the presence of an oxoG error pattern. 'Pre-adapter' indicates oxoG errors that occurred before adapter ligation, such that read 1 carries G->T mutations and read 2 carries C->A mutations, while 'Bait' indicates an oxoG pattern which is exome bait-specific.

— **Presence of Illumina adapters or poly-A tails:** Fraction of reads with respectively Illumina adapters or poly-A tails.

##### 1.8.4 Non-technical covariates

— **PCA covariates:** The top 10 PCA covariates, calculated after sample QC, using an approach described previously, and detailed above[32].

— **Age:** sample age (controls) or age-at-onset (cases). Missing values are imputed to the mean age.

— **AD status:** case or control status

— **Haploblock markers:** to obtain haploblock markers, we select nearby high-quality variants (passing variant QC, with minor allele frequency > 0.025% and a missingness < 10% (missingness defined as read depth < 6)). These variants were phased using Eagle v2.4[39], with default settings. The resulting haploid genotype calls were used as covariates (algorithm detailed below). The region from which these 'nearby' variants are obtained was by default the 50kb up- and downstream from the variant that was tested for batch effects, with the exception of variants that were within 100bp (as there might be complex false positive events that present as multiple variants close together, which could present a false in-linkage signal). The region can be extended from 50kb up to a maximum of 250kb if there are too few variants (<25), or it can be reduced in size if too many are found (>1000).

— **Complex haploblock markers:** In addition, a search is performed for combination of these nearby variants to better mark the haploblock(s) in which the tested variant occurs (detailed below). Allowed Boolean operations are AND and NOT (e.g., a covariate can be defined which is true if variant 1 AND NOT variant 2 are present in a sample).

##### 1.8.5 Forward-backward covariate search

The above covariates are used in a regression model (detailed below) to explain the tested variant. Covariates are selected using a greedy forward selection/backward elimination approach. First, all covariates are normalized to a range 0-1. A covariate set *E* is defined, which contains covariates that are excluded from the regression, that is, their regression parameter is clamped to 0. Furthermore, a covariate set *I* is defined, which contains covariates that are part of the regression: the parameters of these covariates are optimized using a maximum-likelihood approach. Initially, all covariates are in set *E*, and the regression model is fitted using only an intercept.
For all covariates in set *E*, the maximum likelihood gradient is determined. The covariate with the maximum gradient value is selected, and added to set *I*, after which the regression fit is reoptimized. If the AIC (Akaike Information Criterion[44]) score of the fit is improved, this step is accepted, and a new gradient search is performed to select the next covariate. If the AIC however decreases, the variant is removed from set *I.* The above steps are then repeated for the covariate with the next highest likelihood gradient. The forward search is stopped if none of the top 10 covariates improve the AIC metric. If more than 10 covariates are in set *I*, a backward elimination step is performed, in which each covariate in set *I* is in turn dropped from the regression to determine if this improves the AIC score. This step is subsequently repeated every time when 5 new covariates have been added to set *I*.

##### 1.8.6 Prioritizing non-technical covariates

To prioritize non-technical explanations for the presence of a variant, the above feature search is first performed using only non-technical covariates, until no model improvements can be found. The resulting AIC score is noted as the *non-technical score*. Next, technical covariates are added to the covariate set *E*, and the feature search is continued until no model improvements can be found anymore. The resulting score at that point is noted as the *technical score*. The final variant batch detection score is then calculated as the delta between these two scores, that is: *vbd score = technical score - non-technical score*.

#### 1.8.7 Diploid logistic regression model

For haploid genotypes (chromosome Y), the above algorithm can be performed using a logistic regression model, in which $\gamma_j = lr(\alpha + \beta x_j)$ Here, j is the sample, lr is the logistic function, $\alpha$ is the intercept, $x_j$ is the covariate vector for sample j, and $\beta$ is the vector with covariate regression parameters. Normally, in a standard logistic regression, $\gamma_j \in \{0,1\}$. However, due to low coverage data, $\gamma$ is adapted to represent for each sample the probability of the alternate genotype being present (note: not the posterior probability, but the probability given by the variant caller). Standard implementations of logistic regression usually perform a simplification of the maximum likelihood which assumes dichotomous labels. Therefore, a slightly more generic version of logistic regression was implemented which does not make this assumption. Let $p_j(a,\beta) = lr(a + \beta x_j)$. The log-likelihood then takes the following form: $LL(a,\beta) = \sum_j log\left(\gamma_j p_j(a,\beta) + (1-\gamma_j)\left(1 - p_j(a,\beta)\right)\right) - \lambda \sum_i \beta_i^2$. This function is maximized in terms of $a$ and $\beta$. A small regularization term $\lambda = 0.005$ is added to prevent problems with singularities.

In case of diploid genotypes, this model does not suffice, as each sample can have either a reference, heterozygous or homozygous alternate genotype. The approach is to model this as what can be seen as two coupled logistic regression models. Conceptually, in a simplified sense: $d_j = lr(\alpha + \beta g_{j,1} + \theta x_j) + lr(\alpha + \beta g_{j,2} + \theta x_j)$, where $d_j$is a dosage for sample j, in the range [0,2], Here, gj,i is the matrix containing covariates that represents (complex combinations of) phased variants of sample j for haplotype i, and xj is the vector with covariate values for sample j that are haplotype-independent, with vector $\theta$ containing the associated parameter values. Note that the two models share all parameters, but can differ (for phased variants) in their covariates.

More in detail, this is not modelled through dosages, but through genotype probabilities rj, hj and oj, containing respectively the (non-posterior) genotype probabilities of the reference, heterozygous and homozygous alternate genotypes for sample j.

Let $p_{j,i}(\alpha,\beta,\theta) = lr(\alpha + \beta g_{j,i} + \theta x_j)$, which will be noted more shortly as $p_{j,i}$, then the maximum likelihood formulation takes the following form:

$LL(a, \beta, \theta) = \sum_j log(r_j (1 - p_{j,1})(1 - p_{j,2}) + h_j(p_{j,1}(1 - p_{j,2}) + (1 - p_{j,1}) p_{j,2}) + o_j p_{j,1} p_{j,2}) - \lambda(\sum_k \beta_k^2 + \sum_l \theta_l^2)$

To optimize this likelihood (as well as for the logistic regression model above), gradients were derived, and the optimization was implemented using the SLSQP optimizer available through Scipy[41].

##### 1.8.8 Tree search for complex haploblock-markers

Earlier, a forward selection-backward elimination algorithm was described to optimize the set of covariates. The main reason to use such an algorithm is clarified here. To tag a haploblock uniquely, the status of multiple SNPs is usually required to define an accurate marker (e.g. the marker is true if variant 1 is present, but not variant 2). Such markers are needed to define the haploblock(s) in which a tested variant occurs. Adding all possible combination of nearby variants would computationally be prohibitively expensive. Regular variant imputation algorithms have a similar problem, and solve this by using Hidden Markov Models on top of phased population haplotypes. It is however not immediately apparent how such an approach can be combined with a regular covariate regression framework as described above. Instead, to still enable the multi-variant haploblock markers, the forward-backward search is used to explore a tree of increasingly complex multi-variant haploblock markers.

The algorithm starts as described, with a set *E* of all covariates that are inactive, i.e. not part of the regression, and an empty set *I* which will contain all covariates that become ‘active’, i.e. that are selected to be part of the regression model. Next to the covariates that do not represent a genetic variant, set *E* contains at the start only single-variant haplotype markers and no complex multi-variant haplotype markers. That is, the haplotype marker set $Q \subseteq E$ is equal to *M*, where *M* is the set of single-variant markers that are near the tested variant (see section on ‘non-technical covariates’ for how this set of markers is selected). Once a marker $q \in Q$ is moved to set *I*, we extend set *Q* (and thereby set *E*). For a positive association of *q* with the tested variant, we perform: $Q = Q \cup \{q \wedge m, q \wedge \neg m \vee m \in M\}$, while for a negative association of *q* we perform: $Q = Q \cup \{\neg q \wedge m, \neg q \wedge \neg m \vee m \in M\}$. Upon removal of marker *q* from set *I*, the reverse operation is performed. Note that usually in this case, one of the complex markers directly dependent on $q$ has already been added to set *I.*

##### 1.8.9 Detection of missing-not-at-random genotypes

While missing genotype calls are usually only observed due to lack of read coverage, this is not always the case. In certain situations, missingness was found to correlate with genotype status in certain batches (e.g. non-reference calls were more likely to be missing). This is not detected through the above algorithm, as for a missing genotype call all possible genotypes have the same probability, and therefore the sample has, as designed, no effect on the likelihood of the regression model. To detect these situations, the regression model optimized with the non-technical covariates (first step of algorithm) was used to impute the dosage of all samples. Then, a Fisher exact test was performed for each batch and contributing study, to detect possible allele frequency differences between samples for which the genotype call is missing, and for samples for which the genotype is not missing. More in detail, an imputed posterior dosage is determined using the maximum likelihood fit of the 'non-technical' regression model: $d_j = p_{j,1}(1 - p_{j,2}) + (1 - p_{j,1})\, p_{j,2} + 2p_{j,1}p_{j,2}$. Next, an allele-based Fisher exact test (number of alleles is 2 times number of samples) is performed for each batch and study separately, contrasting samples with a missing genotype call with samples with a non-missing genotype call. P-values < $1e^{-6}$ are considered indicative of a problematic batch effect.

##### 1.8.10 Two-phase approach

In some cases, variants that were used as haploblock markers themselves carried large batch effects. Due to this, nearby variants with a similar batch effect pattern were not detected as having such a batch effect. To prevent this from occurring, a two-phase approach was adopted. In the first phase, VBD was run without any haploblock markers. This meant that the non-technical regression model only used the PCA and phenotype covariates. This results in a conservative scoring, as less of the variant is explained by non-technical covariates. Variants that scored a VBD score > 25 in this phase were excluded as haploblock marker in the second phase. In the second phase, the algorithm was then performed as described above, but without the haploblock markers that were excluded by the first phase.

##### 1.8.11 Variant batch correction

For many variants, problematic technical effects were limited to certain batches. In such cases, exclusion of the whole variant seemed unwarranted. To correct these variants, we performed a batch correction step. Variants with a VBD score > 25, or a VBD score > 15 and a MAF < 0.05%, or a batch with a missing genotype batch p-value < 1e-6 were considered for correction. The correction process was performed iteratively, and continued until the VBD score < 10, and the minimum missing genotype batch p-value > 1e-4, or if the variant could not be corrected further. In each iteration, correction was performed in two steps. First, the correction process walked through the technical covariates in order of their addition to the regression model. If such a technical covariate described a batch, study or capture kit and led to an AIC score jump of at least 5, the genotypes for the variant under consideration were set to missing for all samples of such a batch, study or capture kit. This process was stopped once a covariate was encountered that did not fall under these criteria. Second, the correction process walked through all batches with a missing genotype batch p-value <1e-4, which were set to missing as well. If no batches had a p-value <1e-4, but there were contributing studies with a missing genotype p-value <1e-4, then studies were considered instead. Variant were annotated both with VBD results before and after correction.

##### 1.8.12 Variant filtering

Finally, variants were considered for analysis if after correction they had a VBD score < 25, or a VBD score < 15 if they had a MAF < 0.05%.

#### 1.9 Variant selection and annotation

For the association tests, we performed variant selection (**Table 1b** / **Table S3b,c**).

##### 1.9.1 Protein coding transcripts.

We selected variants in autosomal protein-coding genes that were annotated by VEP (version 94.5[45]) to affect the Ensembl basic set of protein coding transcripts. VEP annotates both with Gencode v19 (build 37 native) and Gencode v29 (liftover from build 38)[46]). Transcripts of both Gencode versions were merged based on their identifier, with preference given to the v29-based annotation. Transcripts that passed our filter (protein coding + basic tag) in v19 but not in v29 were not considered.

##### 1.9.2 Variant type.

We only kept variants that directly affected the protein (missense, stop_gained, splice_acceptor, splice_donor or frameshift annotation). For LOF annotations, we only kept those variants with a 'HIGH' VEP impact classification, while for missense annotations we required a 'MODERATE' VEP impact classification.

##### 1.9.3 Variant prioritization.

We prioritized missense variants using REVEL (Rare Exome Variant Ensemble Learner)[47] (annotation obtained from DBNSFP4.1a[48]) and only kept variants with a score ≥ 25 (score range 0 - 100). LOF variants were prioritized using LOFTEE[34] (version 1.0.2), and only LOF variants that had a LOFTEE 'high-confidence' flag were kept.

##### 1.9.4 Variant frequency.

Of these, we only kept variants that were estimated to have at least one carrier, and had a minor allele frequency (MAF) of <1% in both our dataset and the gnomAD v2.1 non-neuro populations.

##### 1.9.5 Variant missingness.

Finally, we removed (5) variants with >20% genotyping missingness (genotypes with a read depth < 6 are considered missing), or that did not pass a filter for differential missingness between the EOAD, LOAD and control groups (Fisher-Exact test comparing EOAD cases versus controls and LOAD cases versus controls). The threshold was set at p<1e-20. For the mega-analysis, this was found to be too strict, due to the increased number of samples leading to increased significance for smaller differential missingness deviations. Therefore, for the mega-analysis, we set the threshold to p<1e-30.

##### 1.9.6 Variant categorization.

Variants were divided in 4 deleteriousness categories: a LOF category, and 3 missense categories: REVEL ≥ 75, REVEL 50-75 and REVEL 25-50 (**Table 1b**).

#### 1.10 Analyses and statistical tests

##### 1.10.1 Gene burden test

Based on previous findings in SORL1, TREM2 and ABCA7[4] an enrichment can be expected of high impact rare risk variants in early onset cases compared to late onset cases. A regular case/control test (in which only a subset of the cases is EOAD) would be inefficient in picking up such signals. The alternative, performing an additional test that specifically tests for burden in EOAD cases, would however also be inefficient as (1) the additional signal from the LOAD cases would be excluded from the analysis and (2) adding such a test would lead to additional correction for multiple testing. Therefore, we combined both case-control and EOAD tests into one, through the use of ordinal logistic regression, where the genetic risk for AD is considered to increase EOAD > LOAD > control. This test is optimally suited for picking up differential variant loads between the sample categories (EOAD > LOAD > Control), but it can also pick up regular case-control signals for which genetic risk is equally distributed across EOAD and LOAD cases (EOAD ~ LOAD > Control) as well as EOAD-specific signals (EOAD > LOAD ~ Control). The burden test was implemented with the ordinal regression implementation available in the MASS package (version 7.3-51.5) for R (version 3.4.3). Six PCA population covariates (calculated on the samples remaining after sample QC, using an approach described previously[37], and detailed above, were used, **Figure S16-S18**), and p-values were calculated using a likelihood ratio test ('lrtest' function from the lmtest package, version 0.9-35). An additive model was considered, by summing the dosages of the minor alleles of selected variants. To prevent biases due to missing or low coverage, we sampled the dosage of each variant call (i.e. 0,1 or 2) according to the posterior probabilities (see above) of the reference, heterozygous or homozygous genotypes. While this sampling provides the same pointwise estimates as an (averaged) dosage approach, it takes into account the uncertainty of the genotype. Contrary to the dosage approach, it allows for a distinction between a genotype with probabilities 0/1/0 (for respectively a reference, heterozygous and homozygous genotype) and a genotype with probabilities 0.33/0.33/0.33 (note that both genotypes have an averaged dosage of 1).

The burden test was performed multiple times with independently sampled genotypes, to account for genotype uncertainty. P-values and beta values were averaged across these runs, while standard deviations were first converted to variances and then averaged. Repeated runs were performed until either the standard deviation of the mean of log10 transformed p-values became < 0.01, 100 runs were reached, or a mean p-value > 0.01 was obtained with at least 25 runs, or a mean p-value > 0.1 with at least 5 runs.

##### 1.10.2 Variant impact thresholds

We tested the evidence for a differential burden for four sets of variants with incrementing levels of predicted deleteriousness: the LOF+REVEL≥25 threshold includes the variants from all deleteriousness categories, while the LOF+REVEL≥50 threshold and LOF+REVEL≥75 threshold condition on the variants with higher levels of predicted deleteriousness. Finally, the LOF threshold includes only variants that are predicted to lead to a complete loss-of-function. The rationale behind this is that for each gene, by concentrating maximum evidence for a differential burden-signal in one test, we maximize the power to identify a differential burden in this gene. Genes were only tested if the cumulative minor allele count (cMAC) of predicted damaging variants was ≥10. Multiple testing correction was performed across all performed tests (up to 4 per gene) using the False Discovery Rate procedure[49]. Genes were considered for replication if the false discovery rate was ≤20%. In order to confirm the AD-association of the genes identified in Stage-1, we used the Stage-2 dataset: p values were corrected using the Holm-Bonferroni method[50], while accounting for number of tests performed in Stage-2. Finally, for the meta-analysis we corrected p values using the Holm-Bonferroni methods, while accounting for the number of tests performed in Stage-1.

Carrier frequency and cumulative Minor Allele Frequency

A carrier of a set of variants was defined as a sample for which the summed dosage of those variants was ≥0.5. Carrier frequencies (CFs) were determined as *#carriers / #samples*. Confidence intervals for the CFs were assumed to be described through a Beta distribution (where *a=#carriers*, and *b=#samples* - *#carriers*). To accommodate situations for certain age-at-onset bins, in which the number of carriers was (close to) 0, a prior was added to a and b based on the carrier count in samples not included in the age-at-onset bin, scaled such that *a=0.1*. The cumulative Minor Allele Frequency (cMAF) for a set of variants and samples was defined as the sum of the minor allele frequencies (MAFs) of the included variants in those samples. When the summed frequency of these variants is <1%, the cMAF can be considered to have a similar uncertainty distribution as the MAF, which can be described using a Beta distribution, where *a=#cumulative Minor Allele Count (cMAC)* and *b=2 * #samples - cMAC*. Similar as for the CF, a prior was added based on the observed allele counts in non-included samples, scaled such that *a=0.1*.

##### 1.10.3 Odds ratios

Effect sizes (odds ratios, ORs) of the ordinal logistic regression can be interpreted as weighted averages of the OR of being an AD case versus control, and the OR of being an early-onset AD case or not. Next to ordinal ORs, we estimated ‘standard’ ORs. This was done across all samples (case/control), as well as per age category (EOAD versus controls and LOAD versus controls), as well as for smaller age-at-onset categories: ≤65 (EOAD), (65-70], (70-80] and >80. Standard ORs were estimated using multinomial logistic regression, using the R nnet package (version 7.3-12), with correction for 6 PCA covariates. For low cMAC values, logistic regression has difficulties in obtaining accurate odds ratios and confidence intervals, as the normal distribution approximation for the log(OR) parameter starts to break down. For these situations (where cMAC≤10, or <3 for either cases or controls), the OR and its confidence intervals were estimated directly based on the cMAF of cases and controls: *OR = (cMAFcase / cMAFcontrol) / ((1 - cMAFcase) / (1 - cMAFcontrol)*. While the uncertainty of this OR is difficult to evaluate directly, it is governed by the uncertainty in cMAFcase and cMAFcontrol. Confidence intervals were therefore estimated through the earlier described beta distribution approximation for the cMAF, by repeated sampling of possible cMAFcase and cMAFcontrol values.

##### 1.10.4 Testing for an association between effect size and variant rareness

To determine if there was a significant trend in effect sizes between the different variant frequency categories (1, 2, 3-5, 6-10, 10+ damaging alleles), an ordinal logistic regression test was performed with constrained beta's $|b_1| \leq |b_2| \leq |b_{3-5}| \leq |b_{6-10}| \leq |b_{10+}|$, and compared to a H0-model with a single beta (**Figure 2C, Table S9**).
Optimization was performed by first estimating b in an unconstrained model, followed by adding the model constraints. Likelihood-ratios in this setting follow a chi-bar-squared distribution. Significance (FDR < 0.05) was therefore determined through sample label permutation, based on the bootstrapping approach outlined in Garre et al[51]. The number of permutations was limited to 10.000.

##### 1.10.5 Sensitivity analysis

A sensitivity analysis was performed to determine if effects were potentially due to age differences between cases and controls (**Figure S2**). An age-matched sample was constructed by dividing samples in strata based on age/age-at-onset, with each stratum covering 2.5 years. Case/control ratios in all strata were kept between 0.1 and 10 by down-sampling respectively controls or cases. Subsequently, samples were weighted using the propensity weighting within strata method proposed by Posner and Ash[52]. Finally, a case-control logistic regression was performed both on the unweighted and weighted case-control labels, and estimated odds ratios and confidence intervals were compared.

##### 1.10.6 Variant-specific analysis

We performed a variant-specific analysis of the genes considered as significantly or suggestively associated with AD, to detect gene-specific idiosyncrasies not covered by our uniform exome-wide analysis. We checked for outlier variants among those that were included in the burden test, determining which ones had a significantly lower or opposite effect size (fisher exact test) compared to other included variants of the same category (missense or LOF). Furthermore, we determined which rare missense or potential LOF variants did associate with AD (logistic regression test, at least 15 carriers), irrespective of REVEL/LOFTEE. We performed corrections for multiple testing per gene using FDR, reporting only variants with a threshold of FDR < 0.2 (**Table S6**). In Stage-2, we replicated these variants, accepting them as true if they attained a (per-gene) Holm-Bonferoni corrected p-value < 0.05. We calculated burden odds ratios both with and without the 3 confirmed outlier variants (**Table 3**).

#### 1.11 Sanger Validation of identified variants

We performed a validation step using an existing dataset containing Sanger validation calls for variants in the SORL1 gene, the gene in which we detected by far the most variants.

a. In a subset of 1,908 samples (from the ADC and Rotterdam Study datasets), we detected 76 singleton variants, and (irrespective of QC status) we tested them all using Sanger sequencing[53]. For the current work, we reanalyzed this dataset in the context of the current pipeline: of the 76 detected SORL1 variants. N=41 SORL1 variant calls passed QC in our current dataset and these were all confirmed through Sanger sequencing (100% true positive rate). For the remaining 35 SORL1 variants: N=8 variants were not present in the current dataset due to sample exclusion (all flagged due to ≤3rd degree family relations (IBD)). N=15 SORL1 variants were excluded in the case-control analysis, as they were flagged by our variant batch detector, mostly due to differences in missingness between cases and controls. For such variants, individual variant calls are usually still reliable, as batch effects are generally derived from the missing calls, indeed, they were all confirmed through Sanger sequencing. N=14 SORL1 variant calls were flagged/not called by our pipeline, and indeed were not confirmed with Sanger sequencing (100% true negative rate).

b. We also obtained Sanger sequencing results for the Rouen study, where Sanger sequencing is performed as part of standard clinical practice and was also collected for several studies[4,5] some of which are not yet published. A total of 69 variant calls that passed QC were tested through Sanger sequencing: 28 in SORL1, 32 in ABCA7 and 9 in TREM2. All variant calls were confirmed as true positives (100% true negative rate).

#### 1.12 Detailed gene discussion

We investigated specific features of the AD-association for the genes identified with the rare-variant analysis using the mega-sample (including exome-extracts, and using the refined burden categories for *TREM2* and *ABCA1*) (**Table 3).** For each gene, we investigated (i) the variant carrier frequency (**Figure 2A),** (ii) the odds ratios of the AD associations (**Figure 2B,C**), (iii) the age at AD onset (**Figure 2A, B**), (iv) missense and LOF categories (**Figure 3**), and (v) variant population frequency (**Figure 2A,D**).

##### *1.12.1 SORL1*

In the ***SORL1*** gene we identified a total of 567 unique coding missense and predicted LOF variants that passed QC (**Table SG1**). The 212 rare variants appertaining to the LOF+REVEL≥50 threshold, carried by 418 individuals, provided the strongest evidence for the AD association (p = 8.1E-26 (**Table 2**). The burden of such variants is concentrated in the younger AD cases: 2.75% of the EOAD cases and 1.51% of the LOAD cases carries at least one such variant compared to 0.68% of all controls. The association with AD is mainly driven by variants which are individually extremely rare and mostly singletons, (151/212 variants were singletons) (**Figure 2D**). Unique for the *SORL1* gene is the significant correlation between lower variant frequency and higher damagingness (**Figure 2C, and Table S9**). LOF variants associated with a 40.7-fold increased risk of EOAD (95%CI 12.5-133) and 11.3-fold increased risk of LOAD (95%CI 3.3-38.3), missense variants with REVEL≥50 associated with a 2.5-fold (95%CI 2.0-3.2) and 1.8-fold (95%CI 1.4-2.3) increased risk of EOAD and LOAD, respectively. In the variant-based analysis in the Stage-1 dataset, we identified two individual *SORL1* variants that associated with AD. We identified a rare variant V1459I (**Table S13**) which was not included in the Stage-1 burden test because of its low REVEL score of 9, but we observed a suggestive association (OR 2.5, 95% CI: 1.22-5.07, FDR: 0.038). In Stage-2 we did not replicate this signal: OR=0.86 (95% CI 0.37-2.01), such that this variant was *not* included in the burden test of the refined analysis of the mega-dataset. Second, we detected the S2175R missense variant as outlier in the Stage-1 dataset (OR=0.53, 95% CI 0.19-1.47; FDR 0.038): it had a significantly lower OR than other missense variants. The Stage-2 dataset included too few carriers, such that we were not able to replicate this effect. Therefore, this variant was not removed from the analysis.

##### 1.12.2 *TREM2*

We identified a total of 95 unique missense and LOF variants that passed QC (**Table SG-2**). In the burden tests, the LOF+REVEL≥25 threshold provided the strongest evidence for an AD association (p=5.2E-22); After refinement, we identified 25 variants appertaining to the LOF+REVEL≥25 threshold, carried by 404 individuals: 2.22% of the EOAD cases and 1.77% of the LOAD cases carries at least one such variant compared to 0.62% of all controls. TREM2 LOF variants associated with a 5.8-fold (95%CI 1.7-19) increased risk of EOAD and 5.4-fold (95%CI 1.8-16.8) increased risk of LOAD (after removal of the transcript specific LOF variant, see **Table S13**). Missense variants associated with a 3.7-fold (95%CI 2.8-4.9) increased risk of EOAD, and a 2.7-fold (95%CI 2.1-3.6) increased risk of LOAD. Although damaging TREM2 variants that drive the AD association are rare, a major fraction of the association signal was carried by missense variant R47H, and recurring A105V and Q33* variants, and the other part by the rarest variants (13/25 variants were singletons). Note that R62H was not included due to an allele frequency > 1% and a low REVEL score (0.04).

In the variant-based analysis (**Table S6**) we identified a significant association for the D87N variant (OR 2.6, 95%CI, 1.6-4.6, MAF: 0.14%, FDR: 0.01) which was not included in the burden test because its REVEL score was too low (20) (**Table S13).** However, we could not replicate this signal in the Stage-2 analysis, such that the variant was not included in the refined analysis of the mega-sample. For LOF variants, we detected an outlier splice acceptor variant rs538447052 (OR: 1.9, 95%CI: 0.7-5.1, MAF: 0.06%), which only affected the non-canonical ENST00000373122 transcript. This variant had a significantly lower odds ratio (outlier FDR: 0.041) compared to the other LOF variants that affect all transcripts. We were able to replicate this outlier effect in the Stage-2 analysis, such that we removed it in the refined analysis of the mega-sample.

##### 1.12.3 *ABCA7*

For the *ABCA7* gene, we identified 684 unique missense and LOF variants (**Table SG-3**). The 351 variants appertaining to the LOF+REVEL≥25 variant threshold, carried by 1,489 individuals provided the strongest evidence for an AD association (p=4.1E-13) (**Table 2**); 6.2% of the EOAD cases and 5.04% of the LOAD cases carries at least one such variant compared to 3.90% of all controls. The AD-association is driven by variants which are individually extremely rare and mostly singletons (190/351 variants were singletons) (**Figure 2C**), but also by several more common variants (**Figure 2D**). LOF and missense variants in the *ABCA7* gene were respectively associated with a 2.2-fold (95%CI 1.4-3.5) and 1.6-fold (95%CI 1.4-1.8) increased EOAD risk, the risk for LOAD was slightly lower. (**Figure 3**). With a variant specific analysis, we identified 3 missense variants detected as outlier in the burden test (**Table S56**): i) R19W (outlier FDR: 1.1%), with an OR of 1.09 (95% CI: 0.4-3.2). ii) V1599M (outlier FDR: 0.1%), with an OR of 0.84 (95%CI: 0.61-1.15, MAF:0.4%) iii) G1820S (outlier FDR 20%, OR: 0.67 (95%CI 0.28-1.6). In the refinement analysis, these variants were not removed as these outlier effects were not replicated in Stage 2. Of note, our discovery analysis excluded two relatively often occurring LOF variants, flagged in our QC pipeline for differential missingness. However, for these variants, it was possible to reliably calculate a single-variant association (by excluding samples with low depth). The first variant is the recurrent splice region variant c.5570+5G>C which previously showed a splicing defect[54] as it fell out of our variant selection criteria (coding exons and canonical, ±2 bp splice sites). A loss of function effect was demonstrated in vitro for this variant[55]. The second variant is the LOF frameshift variant 708-710:EEQ/X (earlier observed by de Roeck et al[56]). Finally, we did not have the possibility to call an intronic variable number tandem repeat (VNTR) variant which was recently associated with an increased risk of developing AD, suggesting that the level of association of ABCA7 in AD is still likely underestimated in our study[57]. Also, it is important to keep in mind that the real impact of some LOF mutations in ABCA7 may be restricted by a transcript rescue mechanism[56].

##### 1.12.4 ATP8B4

We identified 257 variants in the *ATP8B4* gene (**Table SG-4**), and the 94 variants appertaining to the LOF+REVEL≥25 threshold, carried by 850 individuals, provided the strongest evidence for an AD-association (p=9.6E-09) (**Table 2)**; 3.56% of the EOAD cases and 3.1% of the LOAD cases carries at least one such variant compared to 2.1% of all controls. However, unique for the *ATP8B4* gene, the burden was mainly focused on REVEL 75-100 and REVEL 25-50 variants, while other variant categories, as well as singleton variants, did not significantly associate (**Figure 2C**). Instead, the AD-association was driven mainly by one missense variants: G395S with a variant-OR of 1.6 (95%CI1.35-1.91), MAF 0.91%.

Additionally, with a variant specific analysis (**Table S6**), we identified H987R with OR 3.14 (95%CI 1.55-6.34), and MAF 0.03%, which was not added to the burden due to a low REVEL score (26). However, the variant-association did not replicate in Stage 2 (OR 1.58 95%CI 0.45-5.53), such that the variant was not added to the burden in the Mega analysis. Furthermore, we identified variant P83A as an outlier with OR: 0.81 (95%CI 0.28-2.38) in Stage 1, but due to the low number of carriers, the signal did not replicate in Stage 2 (OR 0.85, 95%CI 0.10-6.98) such that this variant was not removed from the burden analysis. Note that the OR point-estimate for *ATP8B4* missense variants (OR=1.5; 95%CI: 1.3-1.7) is higher than the OR for LOF variants (OR=1.1; 95%CI: 0.6-1.9). A possible explanation could be that the risk-increasing effect of the association-driving missense variants depends on a gain-of-function effect rather than on a loss-of-function effect. However, evidence for this will need to be collected by comparing larger sample sizes and functional experiments.

##### 1.12.5 ABCA1

We identified 429 missense and LOF variants in the *ABCA1* gene that passed QC (**Table SG-5**). In the burden analysis, the LOF+REVEL≥75 threshold provided the strongest evidence for an AD-association (p=2.6E-07, **Table 2)**; After refinement, this appertained to 120 variants, carried by 1.5% of the EOAD cases and 1.1% of the LOAD cases, compared to 0.5% of all controls. The AD-association is mainly driven by variants which are individually extremely rare and mostly singletons (80/120) variants are singletons), but also by more common variants, in particular N1800H (MAF: 0.08%). (**Fig 2C,D**). The burden of damaging *ABCA1* variants (LOF+REVEL≥75 variant threshold) is concentrated in younger AD patients. LOF and missense variants in the *ABCA1* gene were respectively associated with a 4.7-fold (95%CI 2.2-10.3) and 2.7-fold (95%CI 1.9-3.8) increased EOAD risk, and this was lower for LOAD cases (**Table 3**). With our variant specific test, we detected 2 variants as outliers: i) a missense variant P85L which had an OR of 0.92 (95%CI 0.56-1.51) and MAF 0.2% (FDR  1.9%) and the outlier signal replicated in Stage 2 with OR 0.85 (0.46-1.56). We found an additional outlier signal for missense variant D1018G with OR 0.81 (95%CI: 0.29-2.22), and MAF:0.05% with FDR  13%. This signal also replicated in the Stage 2 with an OR 0.42 (95%CI 0.13-1.34). Therefore, we removed these variants from in refined analysis.

##### *1.12.6 ADAM10*

We identified 101 missense and LOF mutations that passed QC (**Table SG-6**). The 19 variants appertaining to the LOF+REVEL≥50 threshold, carried by 22 individuals, provided the strongest evidence for an AD-association (p=2.8E-05, **Table 2)**; 0.23% of the EOAD cases and 0.05% of the LOAD cases carries at least one such variant compared to 0.02% of all controls. With the rare occurrence of such variants (16/19 variants are singletons), it is difficult to detect an exome-wide significant signal, even for variants with the strongest AD-associations in this large sample. We found that LOF+REVEL≥50 variants were suggestively associated with a 9.0-fold (95%CI 2.9-28) increased risk of EOAD. We note that one splice-acceptor LOF variant, carried by a single control in Stage-1, only affects transcripts ENST00000402627 and ENST00000561288. These transcripts, being 71 and 38 amino acids long, miss the majority of the canonical transcript (748 amino acids). This individual was last checked at age 89.

##### 1.12.7 RIN3

For the *RIN3* gene, we identified 278 unique missense and LOF variants (**Table SG-7**). The 23 variants appertaining to the LOF+REVEL≥50 variant threshold, carried by 583 individuals, provided the strongest evidence for an AD association (p=1.6E-05) (**Table 2**); 2.7% of the EOAD cases and 2.1% of the LOAD cases carries at least one such variant compared to 1.6% of all controls. While 14/23 variants were singletons, the majority of the AD-association was driven by 2 more common variants:  Y793H (MAF 0.84%) and W63C (MAF: 0.08%) (**Figure 2C,D**). *RIN3* shows moderate effect sizes: LOF variants in the *RIN3* gene were associated with a 2.9-fold (95%CI 0.6-32.0) and 1.7-fold (95%CI 0.4-18.6) increased risk of EOAD and LOAD, respectively, while missense variants associated with a 1.6-fold (95%CI 1.3-2.0) and 1.3-fold (95%CI 1.1-1.6) respectively (**Figure 3, Table 3**).

##### *1.12.8 CLU*

For the *CLU* gene, we identified 105 unique missense and LOF variants (**Table SG-8**). The 24 variants appertaining to the LOF+REVEL≥25 variant threshold, carried by 26 individuals, provided the strongest evidence for an AD association (p=5.0E-04) (**Table 3**); 0.23% of the EOAD cases and 0.09% of the LOAD cases carries at least one such variant compared to 0.03% of all controls. Most variants were extremely rare, 22/24 were singletons (**Figure 2C,D**). We observed large effect sizes. LOF variants in the *CLU* gene were associated with a 14.2-fold (95%CI 2.9-470.4) and 3.8 (0.6-122.4) increased EOAD and LOAD risk respectively (**Figure 3, Table 3**).

##### *1.12.9 ZWCWP1*

For the *ZWCPW1* gene, we identified 117 unique missense and LOF variants (**Table SG-9**). The 11 variants appertaining to the LOF variant threshold, carried by 15 individuals, provided the strongest evidence for an AD association (p=7.8E-04) (**Table 2**); 0.15% of the EOAD cases and 0.05% of the LOAD cases carries at least one such variant compared to 0.01% of all controls. The AD signal was driven by LOF variants only, each of which was very rare, and 8/11 variants were singletons (**Figure 2C,D**). Effect sizes of ZWCPW1 were also large, with LOF variants being associated with a 9.1-fold (95%CI 3.1-90.1) and 2.9-fold (95% CI: 0.8-27.4) increased risk of EOAD and LOAD, respectively (**Figure 3, Table 3**).

##### *1.12.10 ACE*

For the *ACE* gene, we identified 363 unique missense and LOF variants **Table SG-10**). The 38 variants appertaining to the LOF+REVEL≥75 variant threshold, carried by 99 individuals, provided the strongest evidence for an AD association (p=9.0E-04) (**Table 3**); 0.60% of the EOAD cases and 0.39% of the LOAD cases carries at least one such variant compared to 0.20% of all controls. (**Figure 2D,E**). Effect sizes were moderate. LOF variants associated with 1.7-fold (95% CI: 0.9-3.4) and 1.2-fold (95% CI: 0.6-2.2) increased risk for EOAD and LOAD respectively. Remarkably, missense variants showed a larger association of 3.9 (95% CI: 1.8-8.8) and 2.7 (95%CI: 1.3-5.9) respectively (**Figure 2F, Table 3**).

### 2 Supplemental Figures and Tables

#### 2.1 Figures

##### 2.1.1 Figure S1: Age, gender, APOE genotype distribution

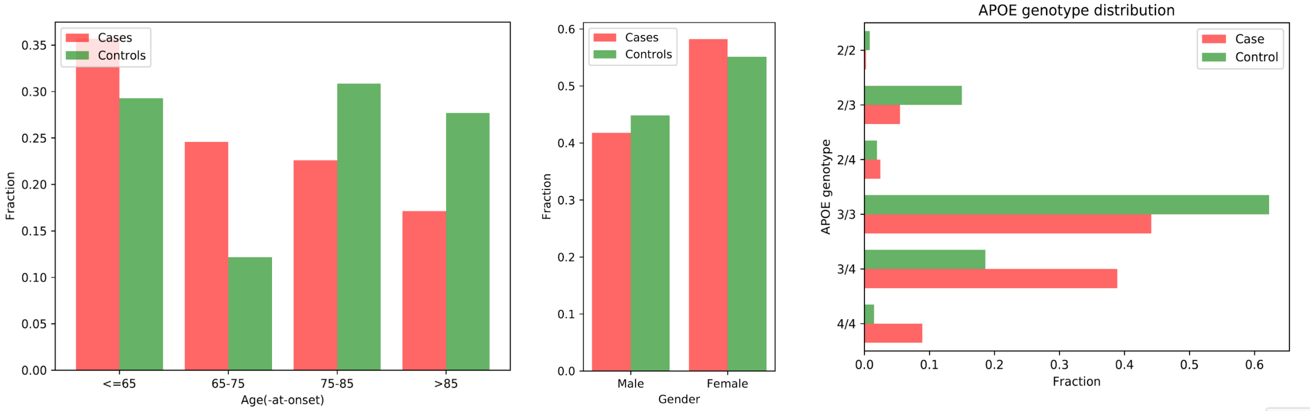

Age, gender and APOE genotype distribution of all samples, stratified by case/control status.

#### 2.1.2 Figure S2: Sensitivity Analysis: AD vs Age association

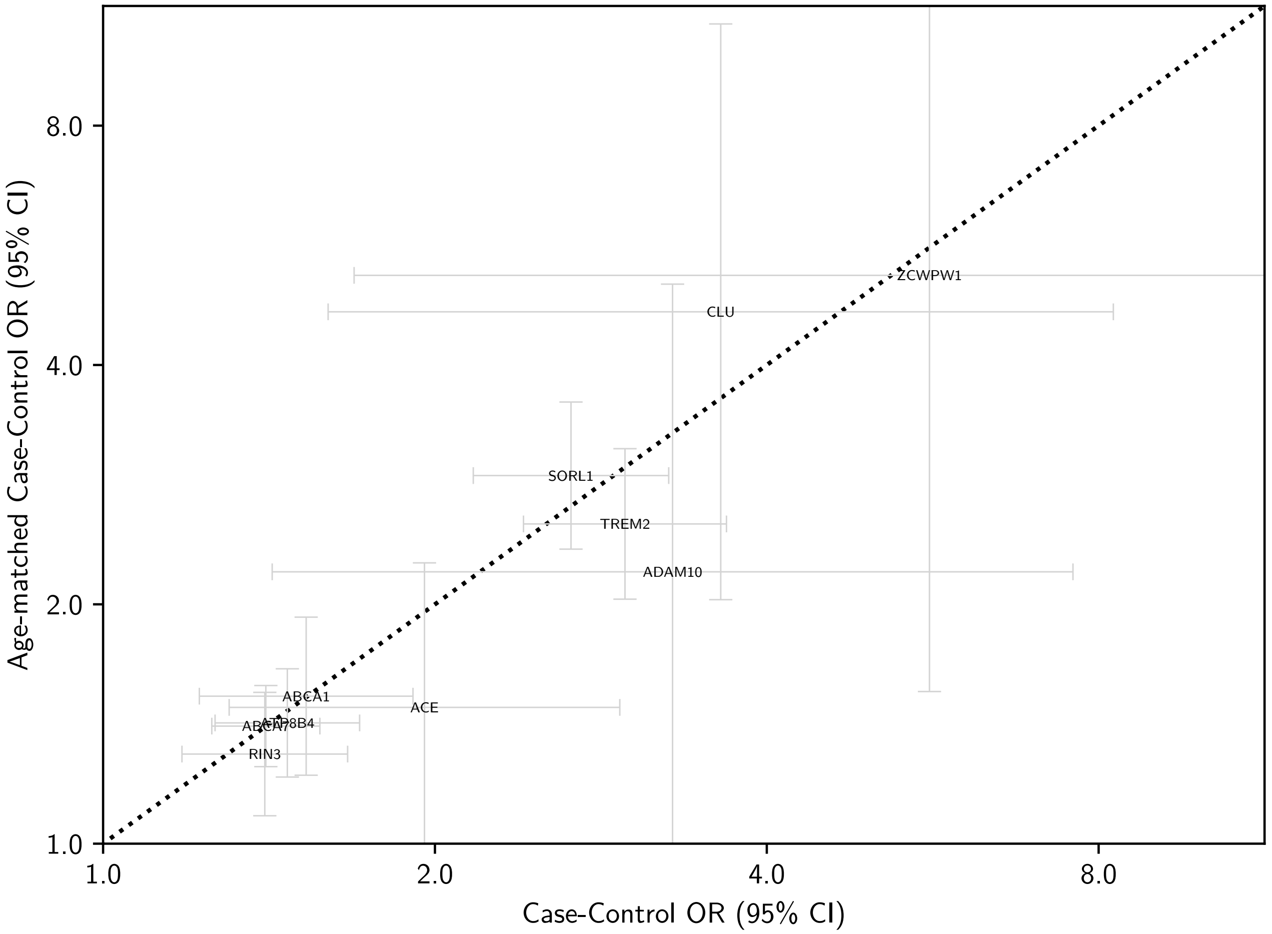

Sensitivity analysis of the gene burden tests (for the most significant deleteriousness thresholds, **Table 2**). Comparison of the case/control odds ratio of an age-matched and a normal analysis. Age-matching was performed as described in the methods. Based on the confidence intervals, we cannot exclude that the signals in *ACE*, *ADAM10* and *ZCWPW1* are affected by other age-related conditions. Note however, that the signals in *ADAM10* and *ZCWPW1* are based on very few variants, such that confidence intervals are expected to be wide.

#### 2.1.3 Figure S3: Read length per study

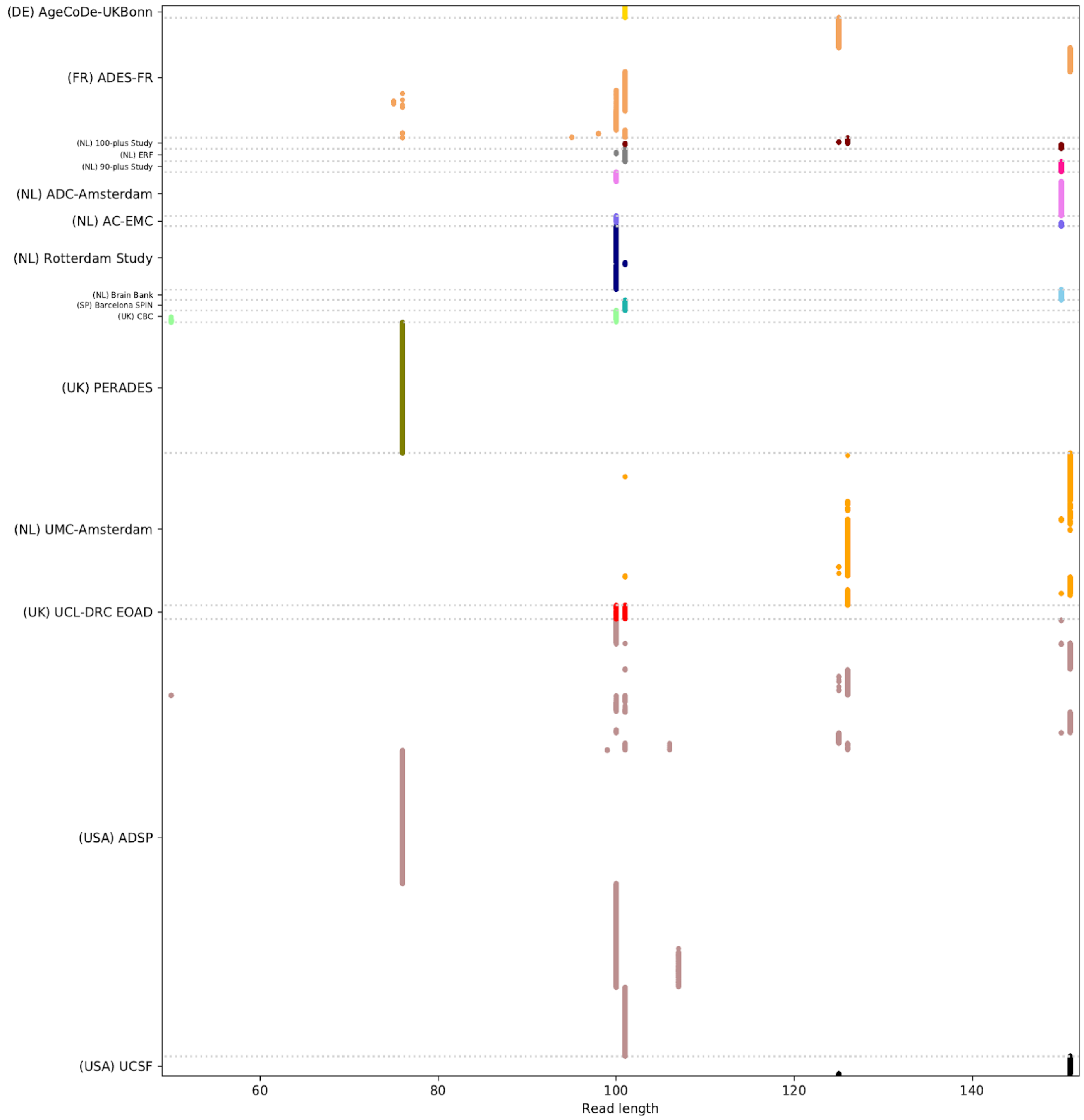

Illumina read length, by study for all samples (dots).

#### 2.1.4 Figure S4: Genotype Quality

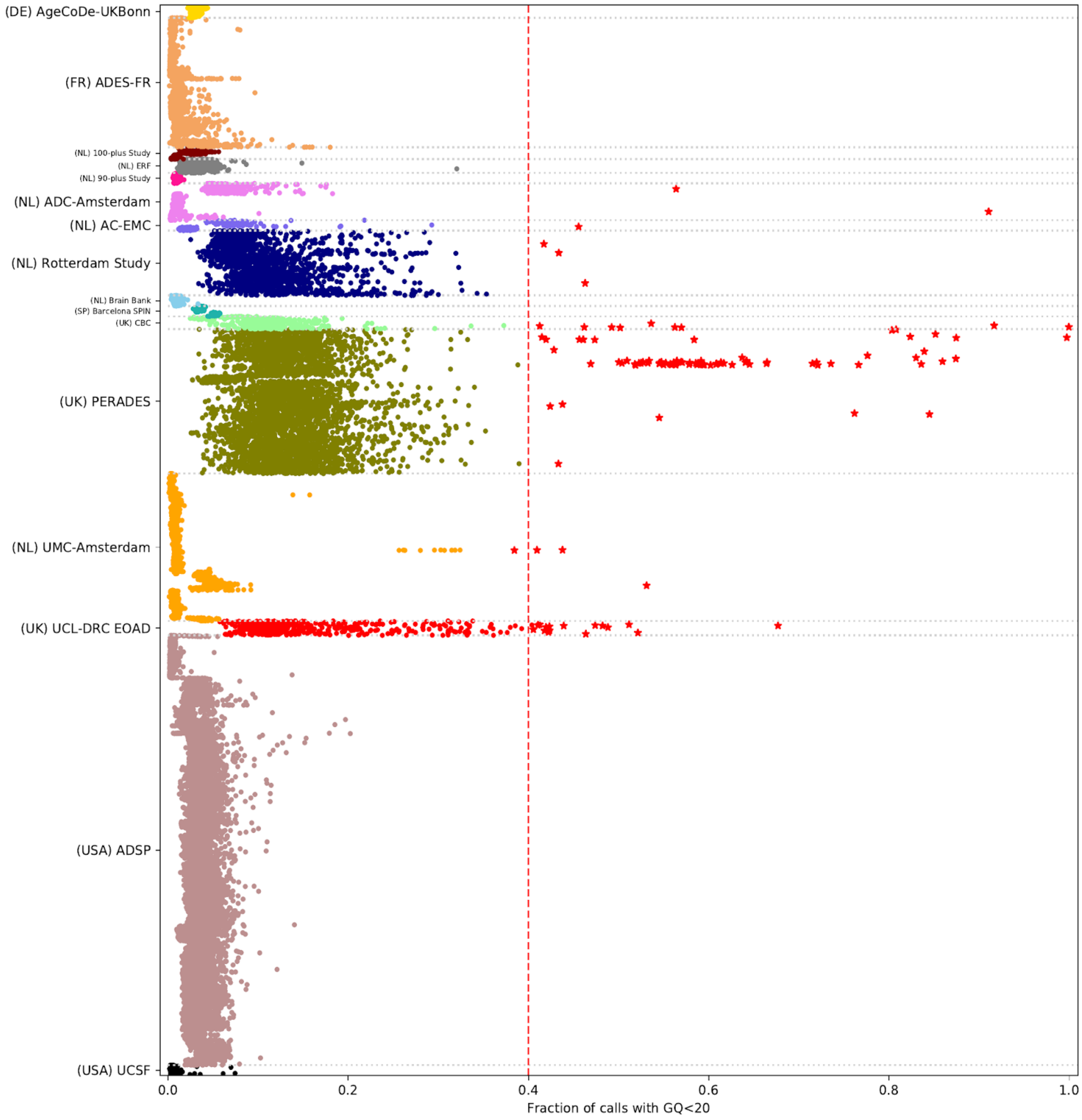

Fraction of genotype calls with a genotype quality < 20. Each sample was evaluated in context of its capture kit. Samples that are considered outliers due to missingness are indicated with a red '*' symbol.

#### 2.1.5 Figure S5: Genetic sex

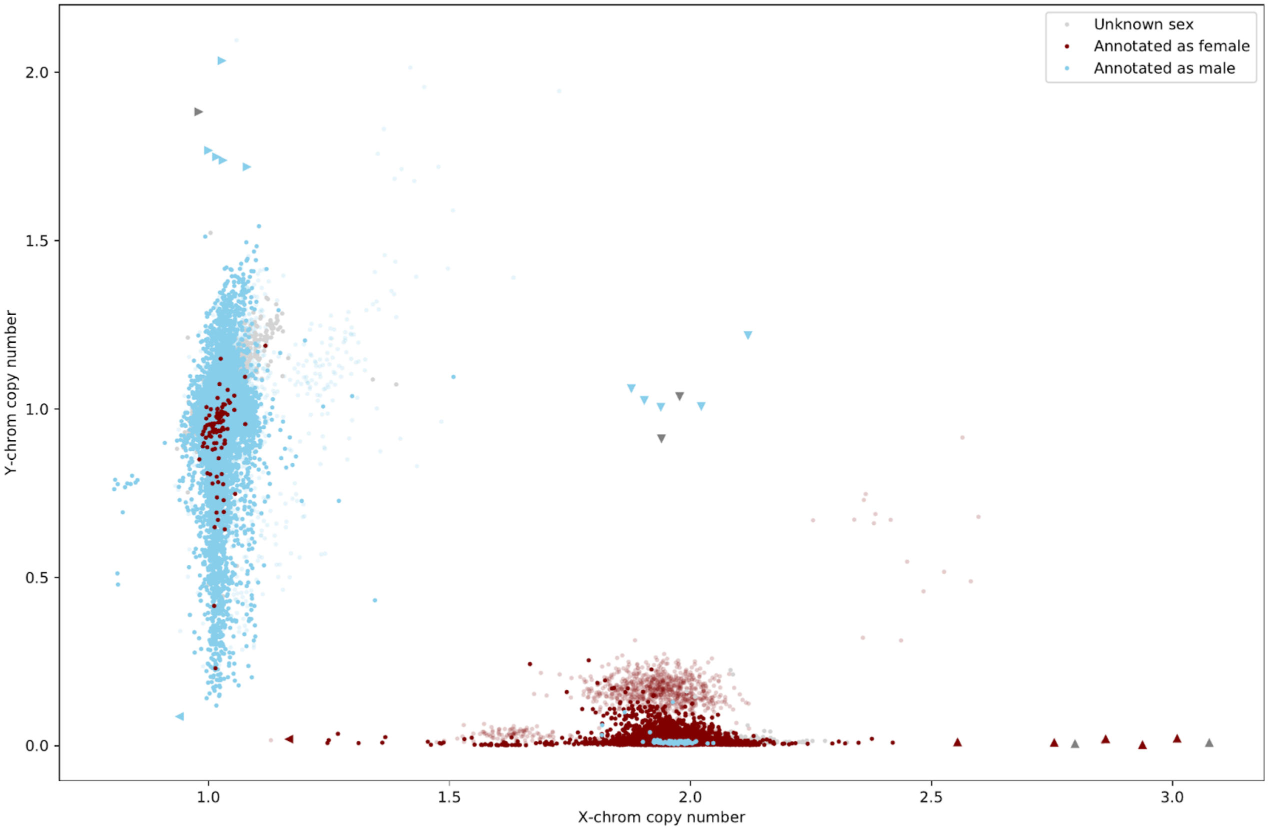

Check of sex chromosome copy number versus clinical sex annotation. Samples that failed the sex check were plotted last to increase their visibility. Samples that were classified as XXY, XXY and XXX are indicated by respectively right, down and upwards pointing triangle symbols. Samples with increased uncertainty due to low coverage were plotted with increased translucency. A number of samples failed this check, and were found to be enriched in 3 sequencing plates. Samples from these plates were excluded from the analysis.

#### 2.1.6 Figure S6: PCA: Sample population compared to 1,000G population samples

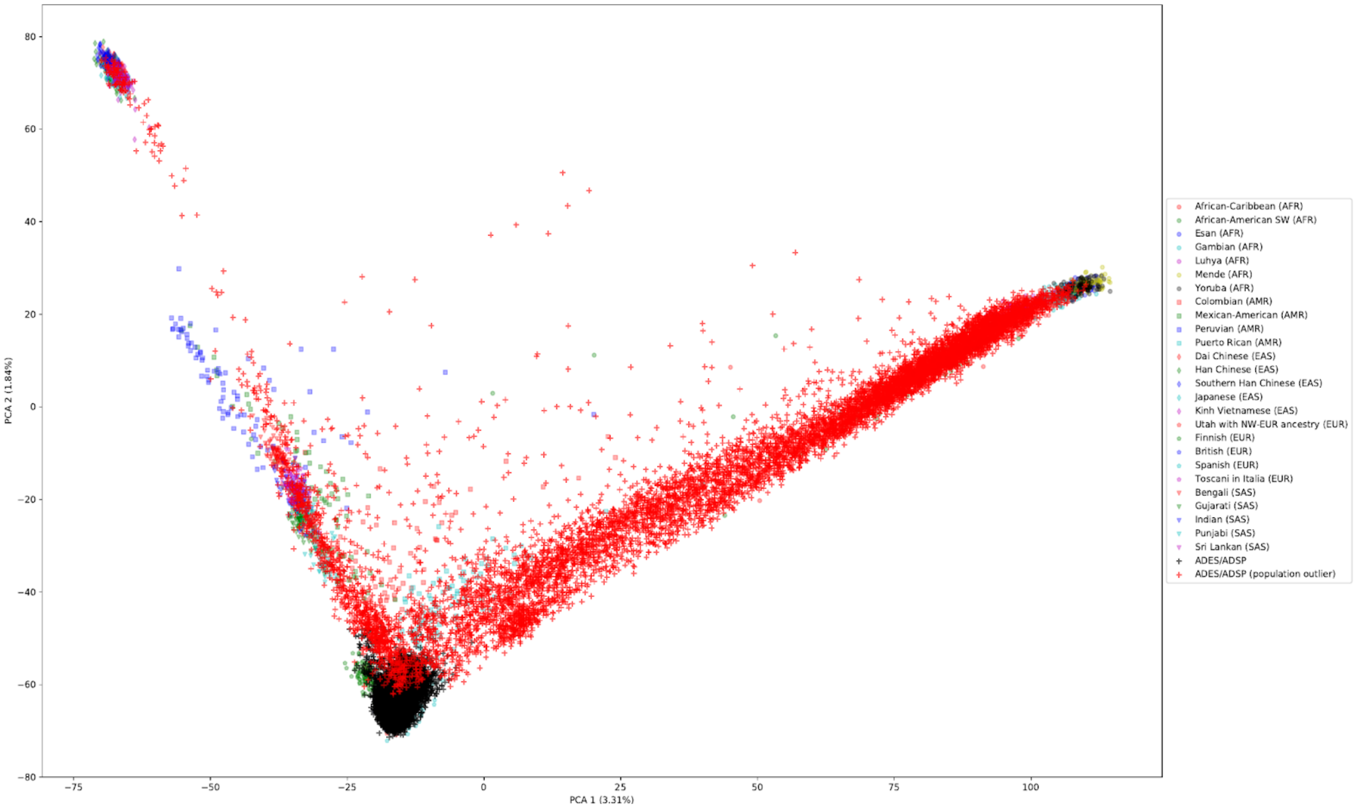

First two PCA components of the study samples, together with 1000 genome samples for reference. Samples in red are considered population outliers. For Stage-1 and Stage-2 studies, except samples with only exome-extracts.

#### 2.1.7 Figure S7: first two population PCA components per study

First two PCA components per study. Samples indicated as a ‘x’ are outliers.

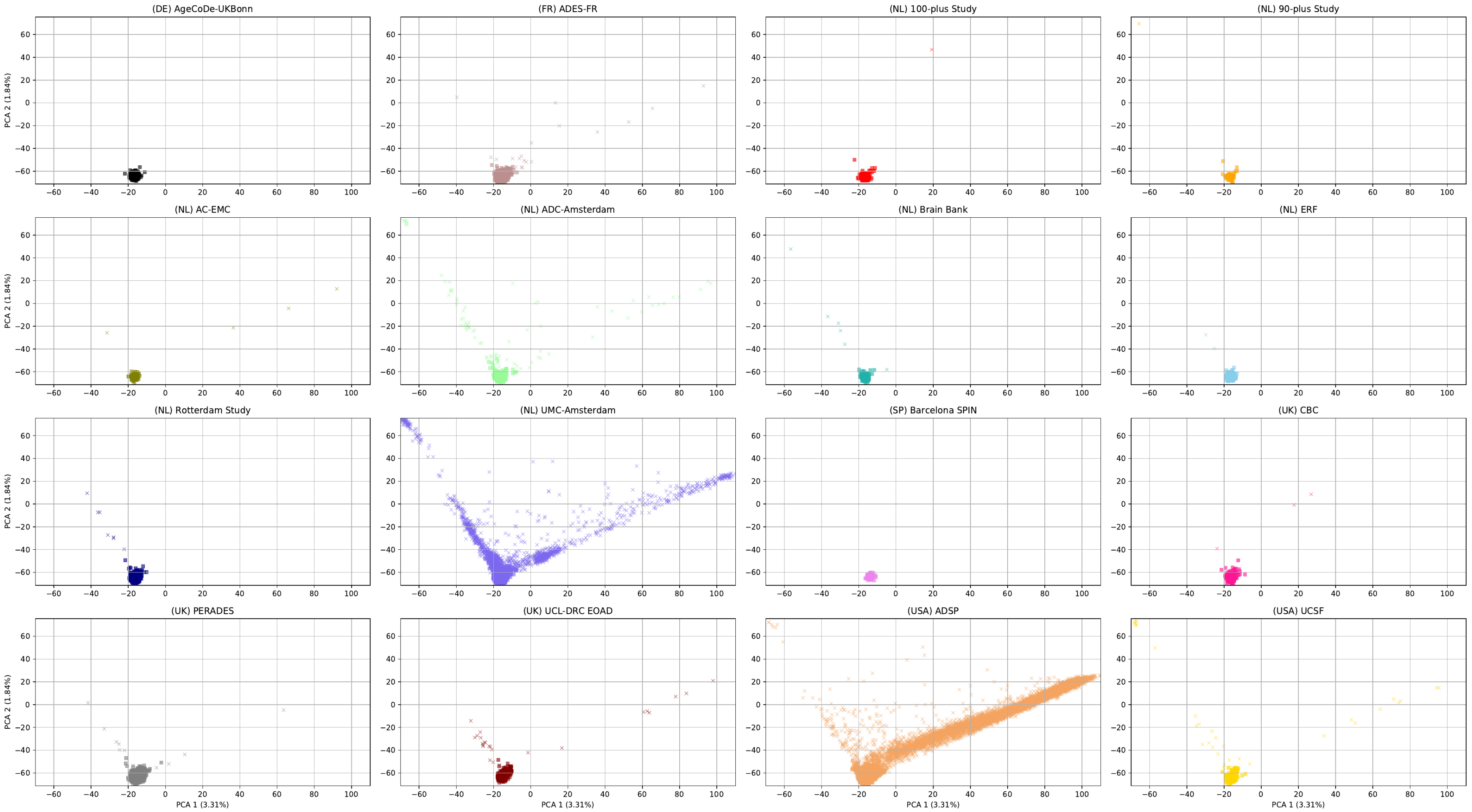

#### 2.1.8 Figure S8: Third and fourth population PCA components per study

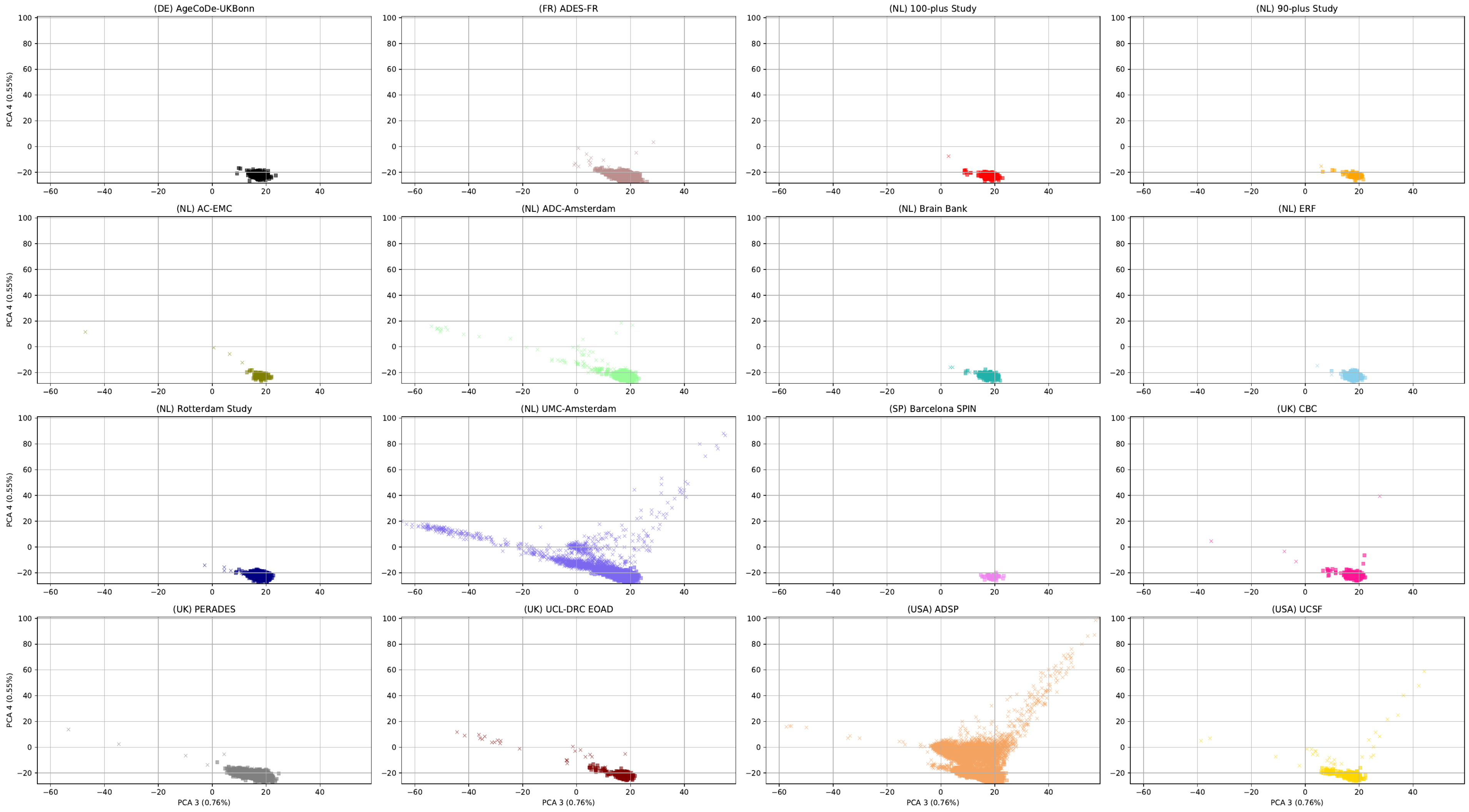

Third and fourth PCA component for each study. Samples indicated as a 'x' are outliers.

#### 2.1.9 Figure S9: Number of novel SNPs (union of capture kits)

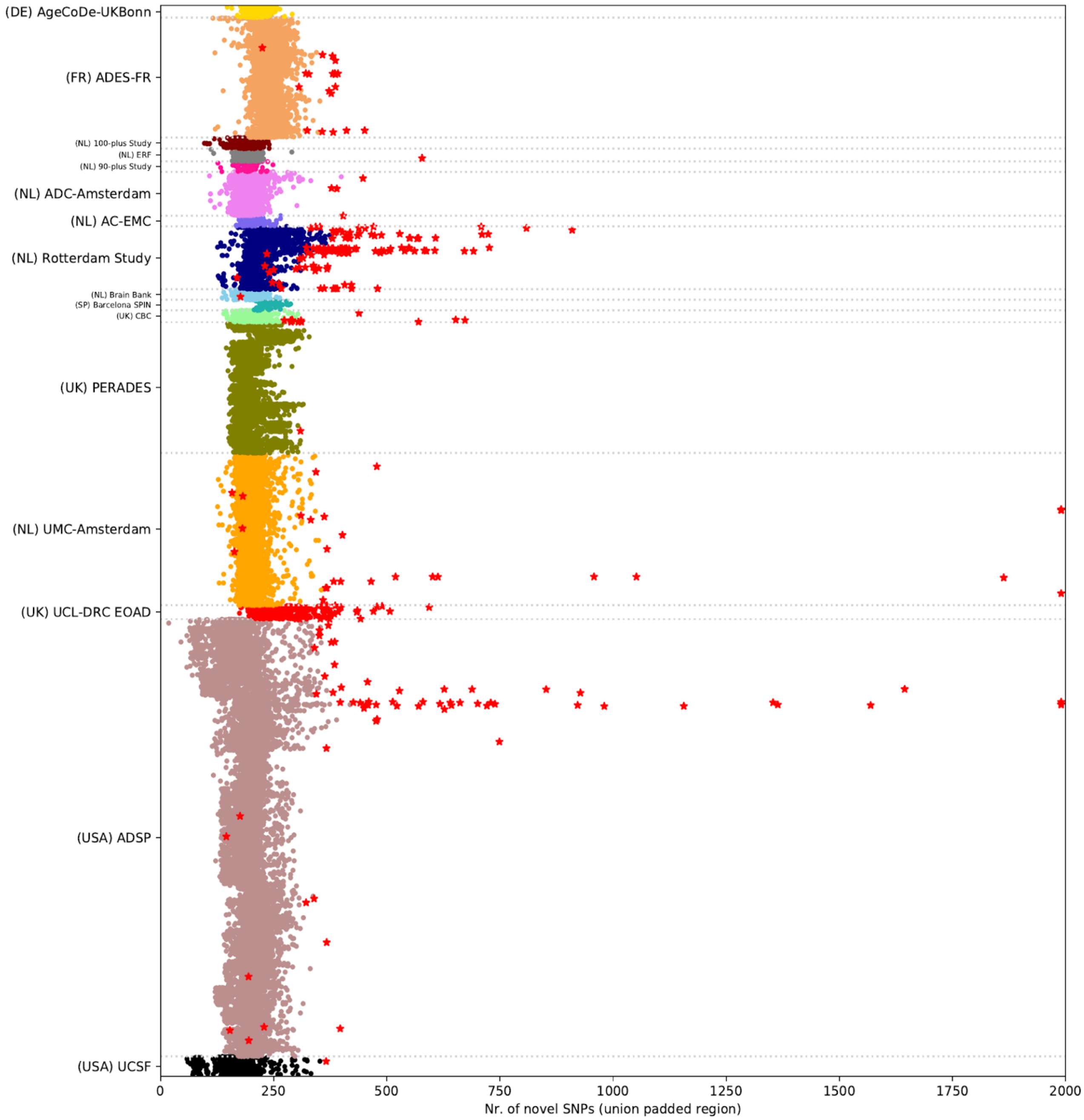

Nr. of novel SNPs per sample, in the region representing the union of all capture kits + 100bp padding. QC outliers are shown as red stars. Variants are classified as novel if they are not present in DBSNP v150. Per geographical region, the comprehensiveness of the annotation of local rare variants in DBSNP might vary.

##### 2.1.10 Figure S10: Number of novel indels (union of capture kits)

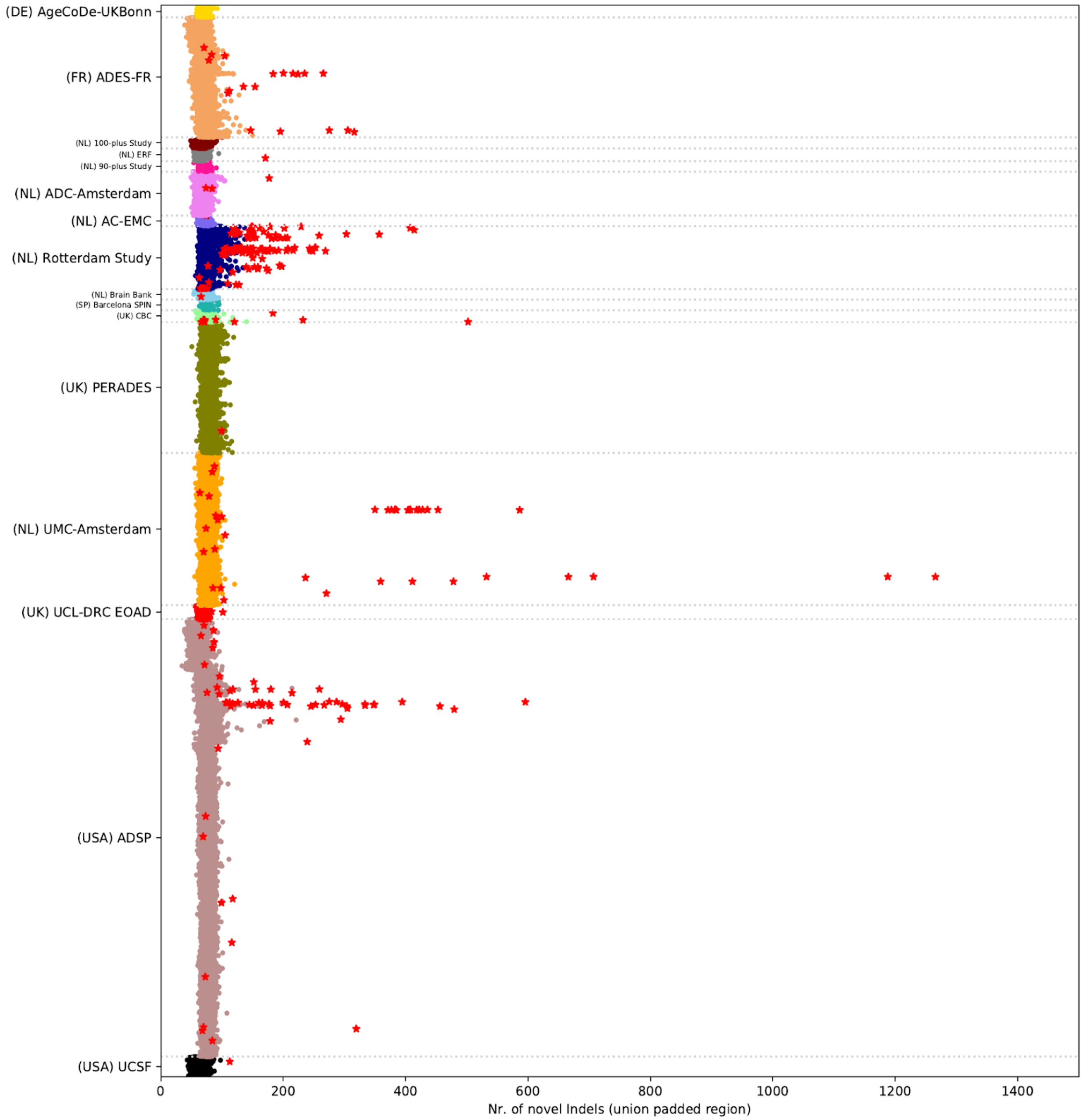

Nr. of novel indels per sample, in the region representing the union of all capture kits + 100bp padding. QC outliers are shown as red stars. Variants are classified as novel if they are not present in DBSNP v150. Per geographical region, the comprehensiveness of the annotation of local rare variants in DBSNP might vary.

#### 2.1.11 Figure S11: Number of novel SNPs (intersection of capture kits)

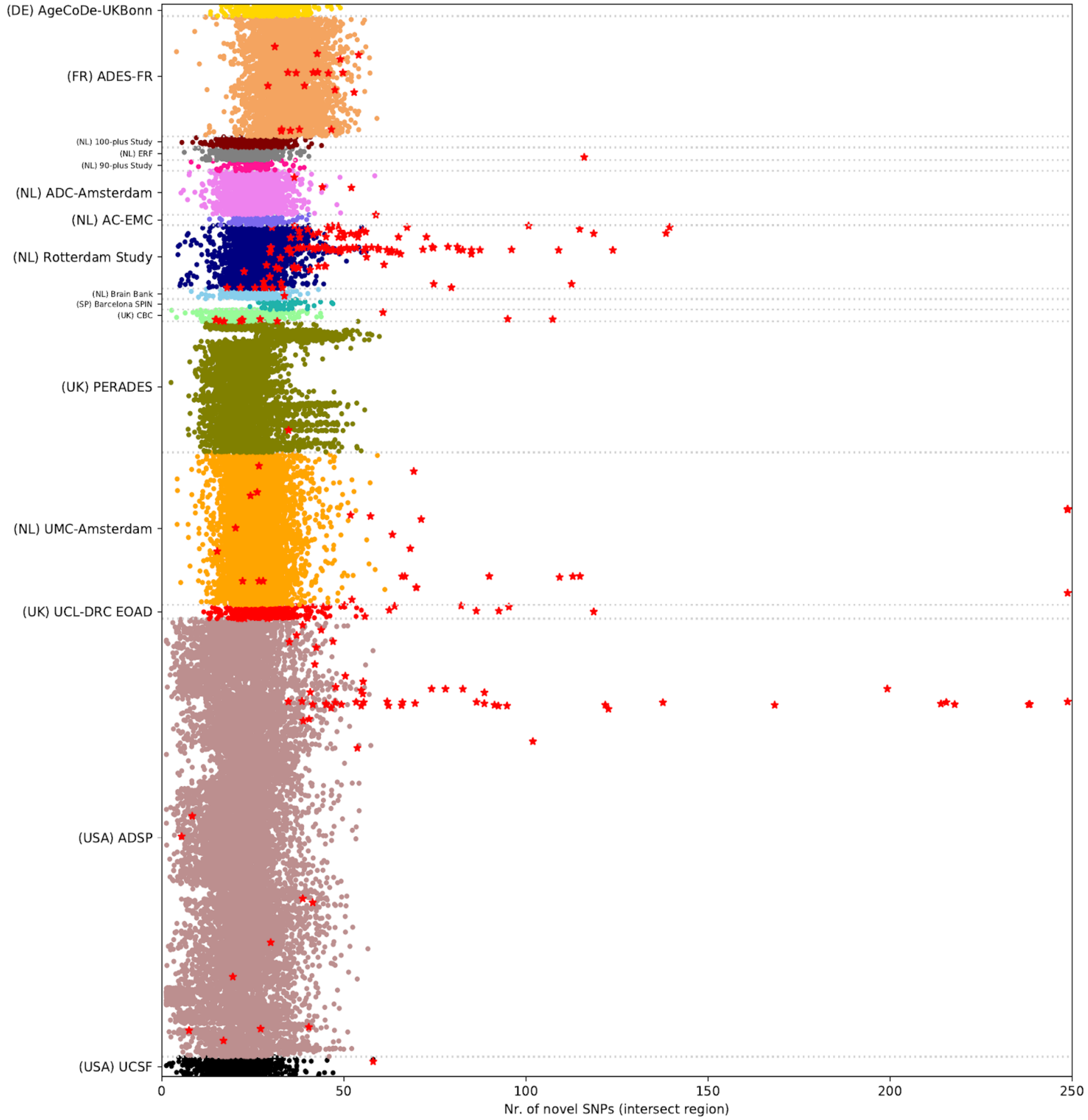

Nr. of novel SNPs per sample, in the intersection of all capture kits. QC outliers are shown as red stars. Variants are classified as novel if they are not present in DBSNP v150. Per geographical region, the comprehensiveness of the annotation of local rare variants in DBSNP might vary.

#### 2.1.12 Figure S12: Number of novel indels (intersection of capture kits)

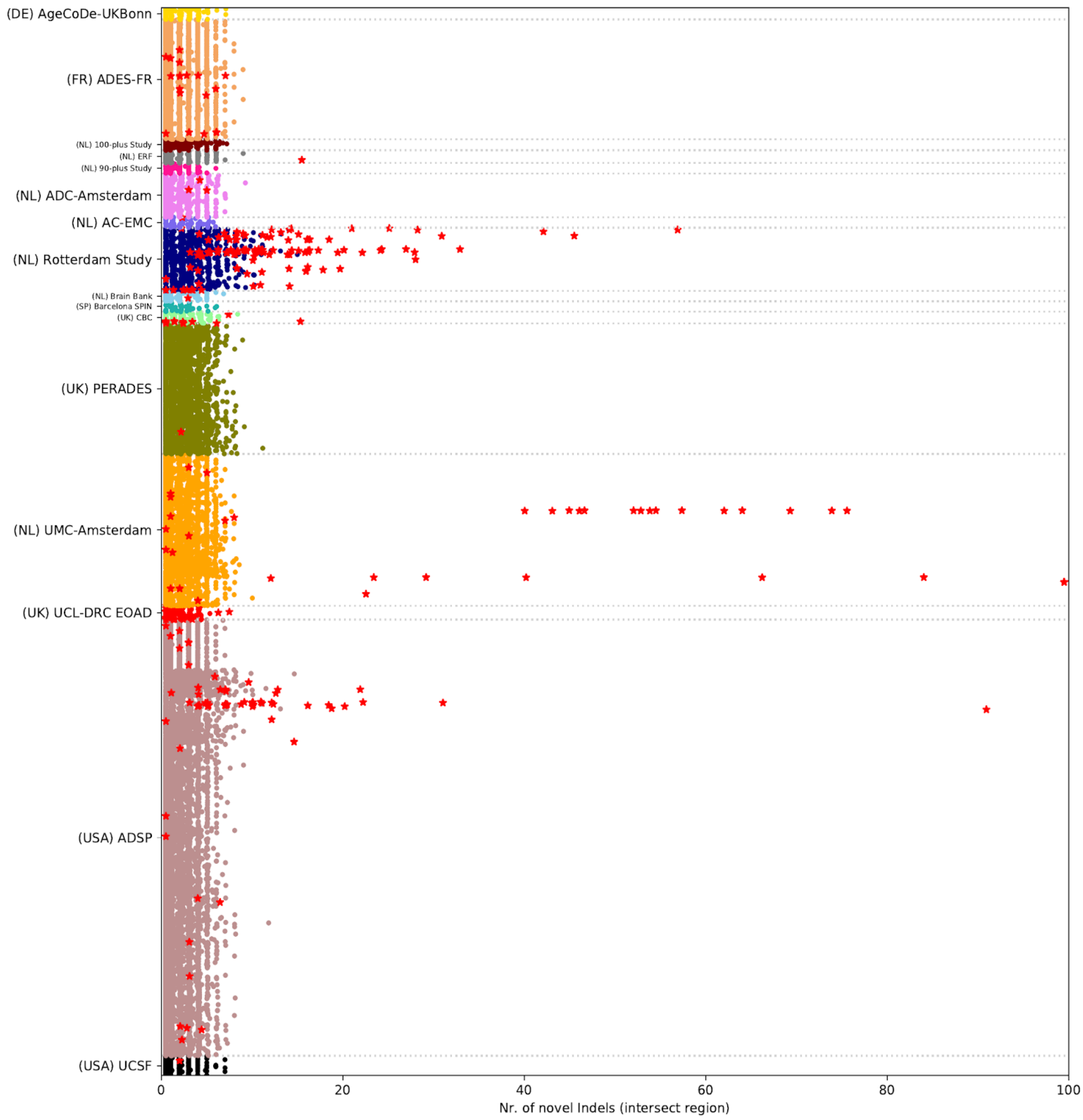

Nr. of novel indels per sample, in the intersection of all capture kits. Sample QC outliers are shown as red stars. Variants are classified as novel if they are not present in DBSNP v150. Per geographical region, the comprehensiveness of the annotation of local rare variants in DBSNP might vary.

#### 2.1.13 Figure S13: Ts/Tv ratio known variants (intersection capture kits)

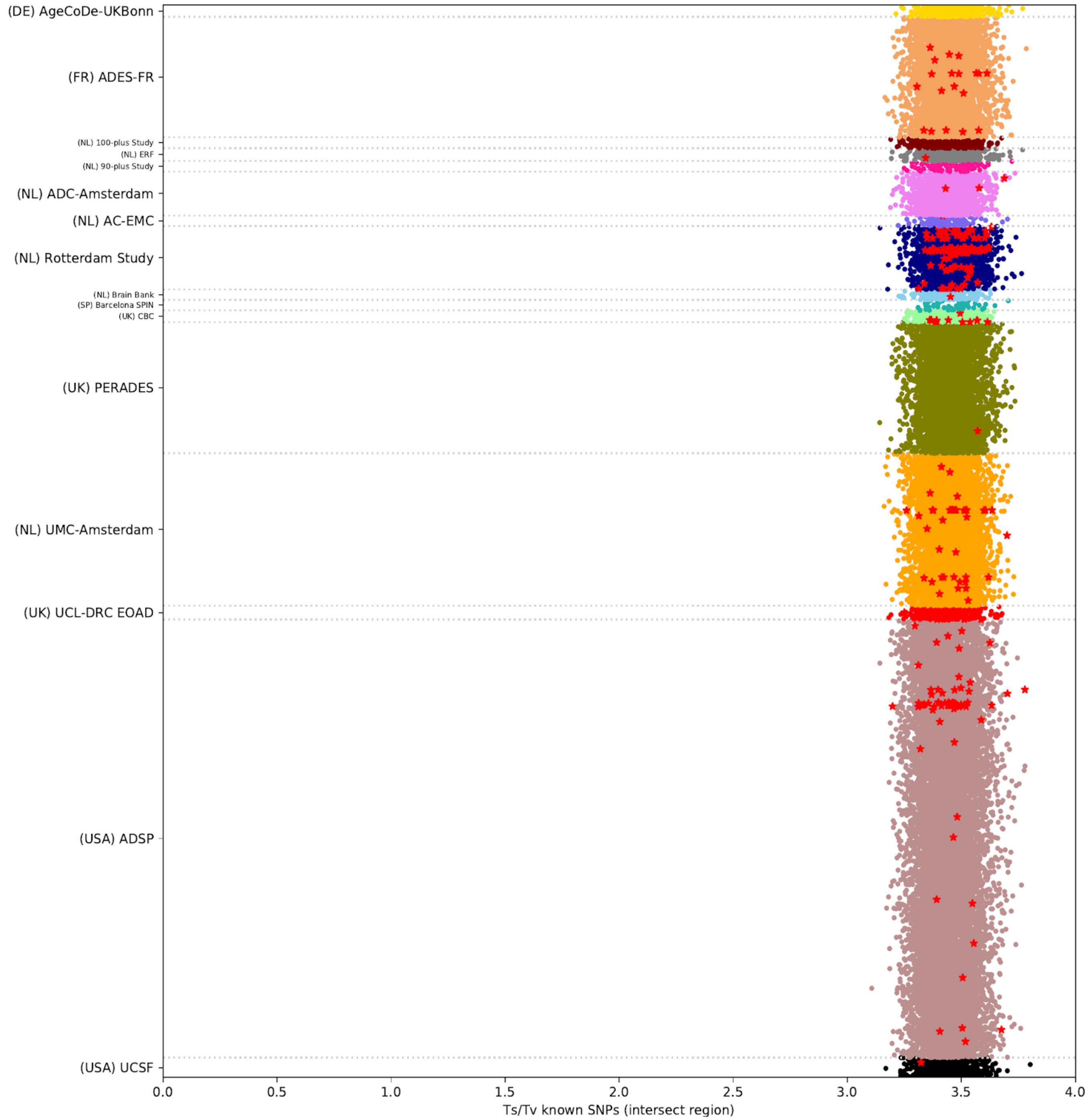

**Transition/Transversion ratio per sample,** of known variants in the region covered by all capture kits. QC outliers are shown as red stars. Variants are classified as known if they are present in DBSNP v150.

#### 2.1.14 Figure S14: Ts/Tv ratio novel variants (intersection of capture kits)

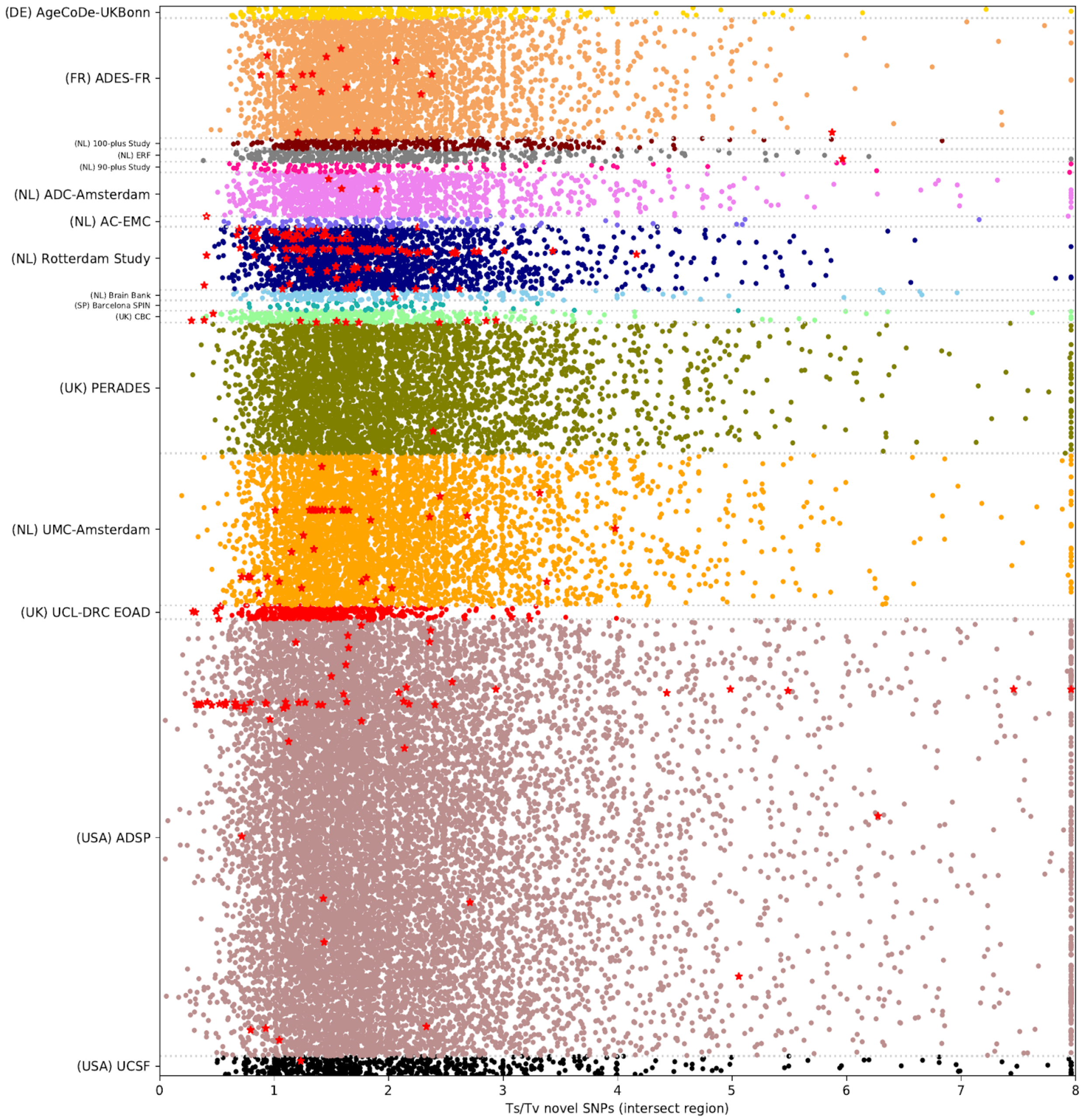

Transition/Transversion ratio per sample, of novel variants in the region covered by all capture kits. QC outliers are shown as red stars. The distribution is wide due to a low number of novel SNPs per sample (**Figure S11**). Ts/Tv values are for plotting purposes maximized at 8. Variants are classified as novel if they are not present in DBSNP v150.

##### 2.1.15 Figure S15: Het/Hom ratio known variants (intersection capture kits)

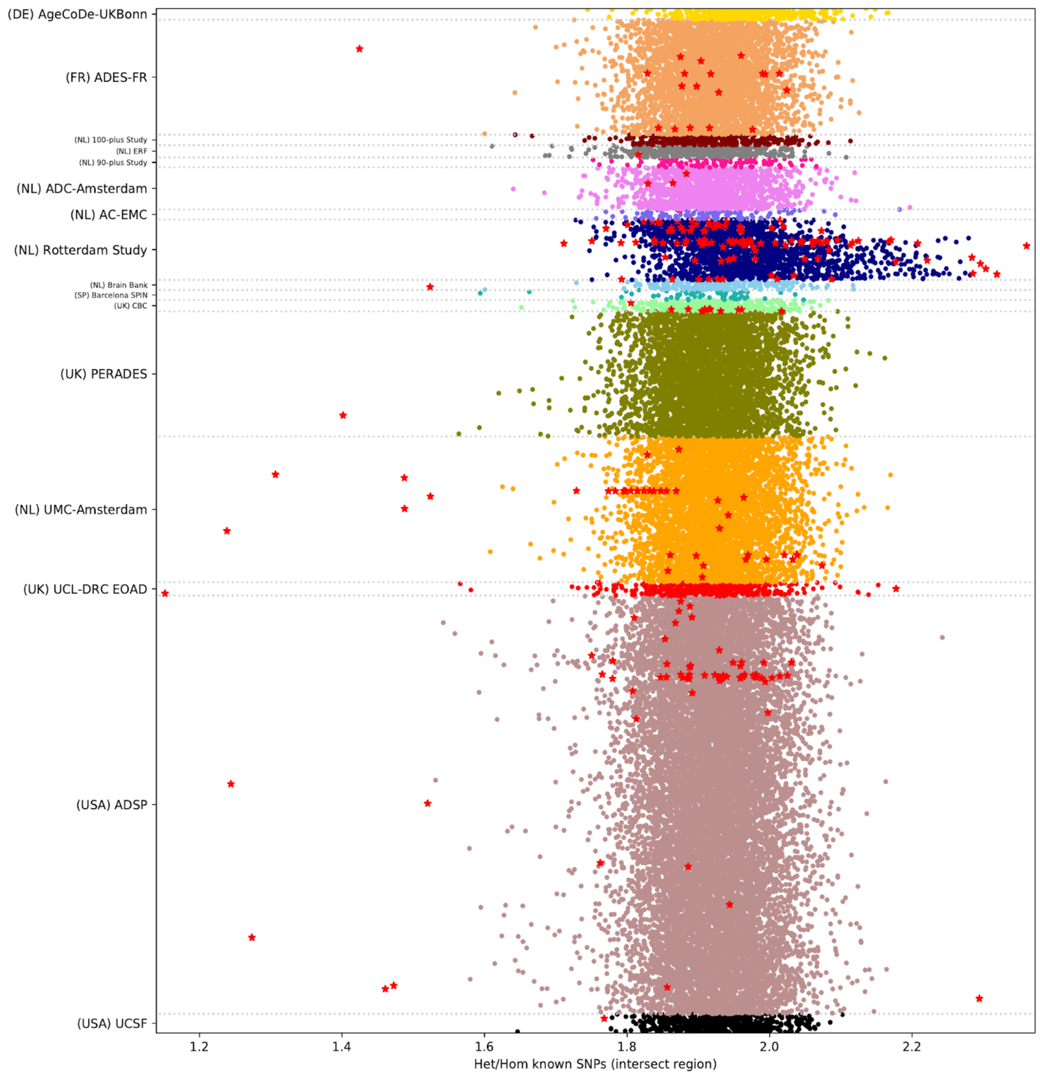

Heterozygous/Homozygous ratio per sample, of known variants in the region covered by all capture kits. Sample QC outliers are shown as red stars. Variants are classified as known if they are present in DBSNP v150. Low het/hom ratios can be an indication of inbreeding, while high het/hom ratios can be an indication of outbreeding or sequence contamination. The problem of contamination is mostly limited to more common variants, and not the rare variants that are the focus of this study

#### 2.1.16 Figure S16: First two PCA components per study, after sample QC.

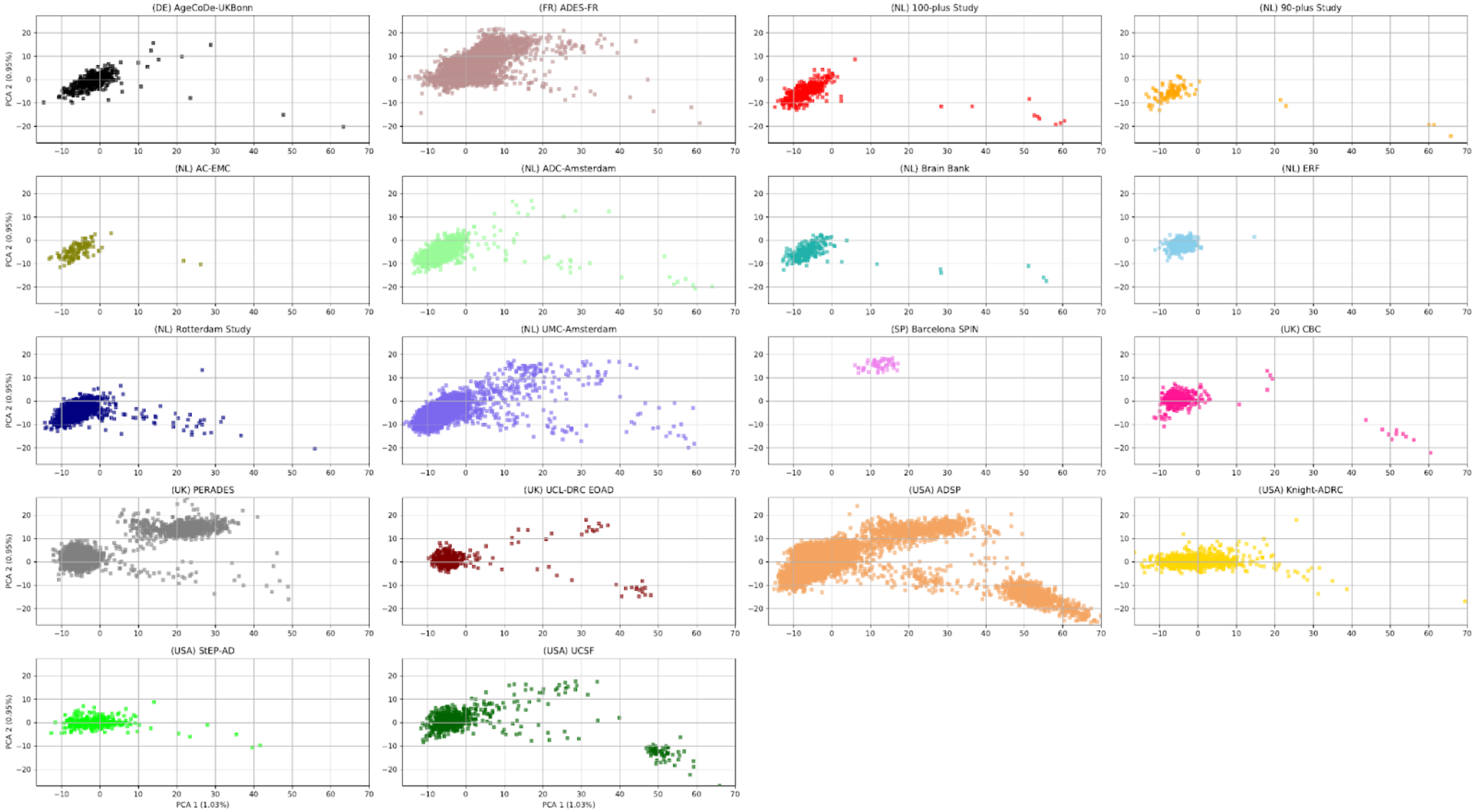

First two PCA covariates after sample QC. All analysis are corrected for the first 6 PCA components.

#### 2.1.17 Figure S17: Third and fourth PCA components per study, after sample QC.

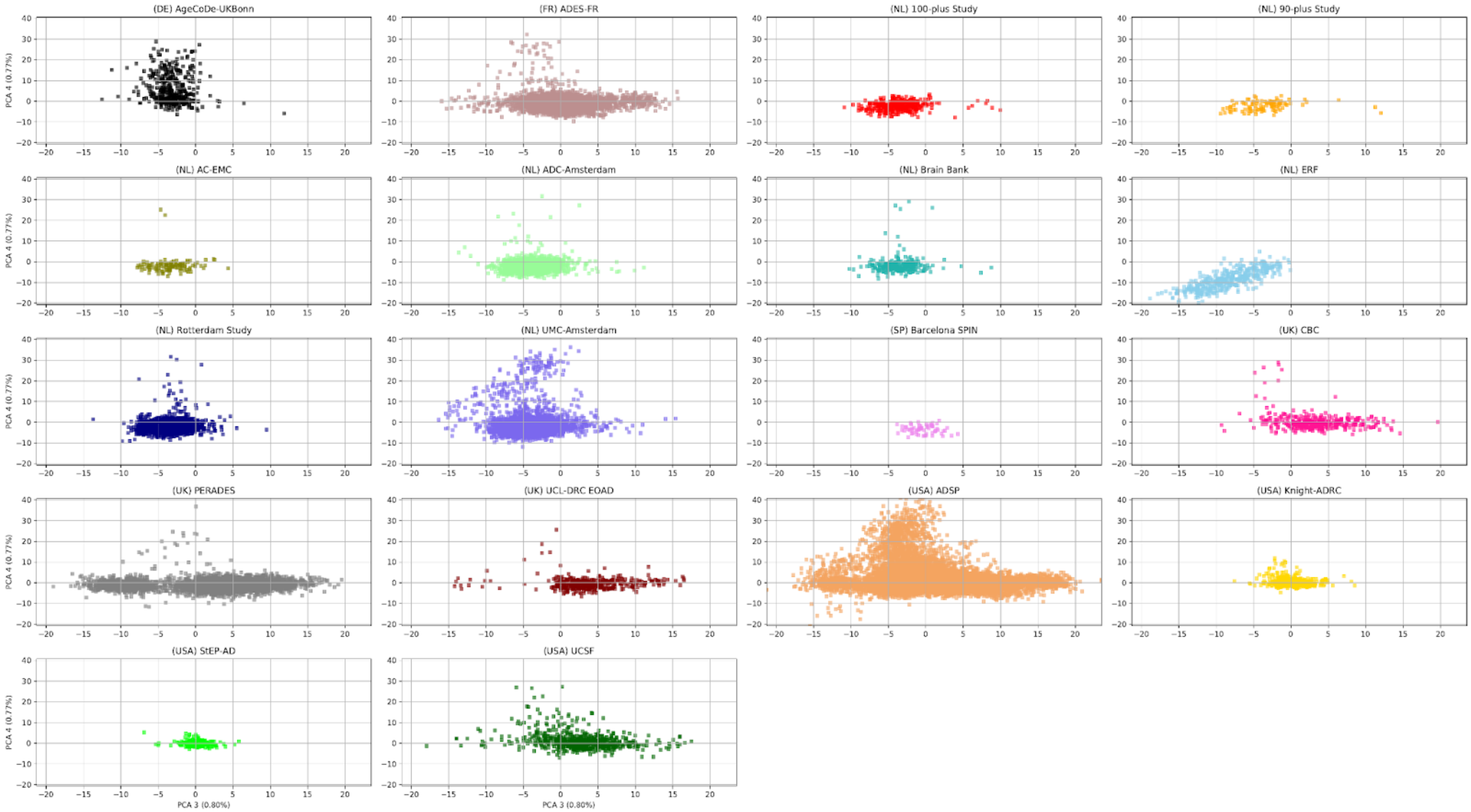

Third and fourth PCA covariates after sample QC. All analysis are corrected for the first 6 PCA components.

#### 2.1.18 Figure S18: Fifth and sixth PCA components per study, after sample QC.

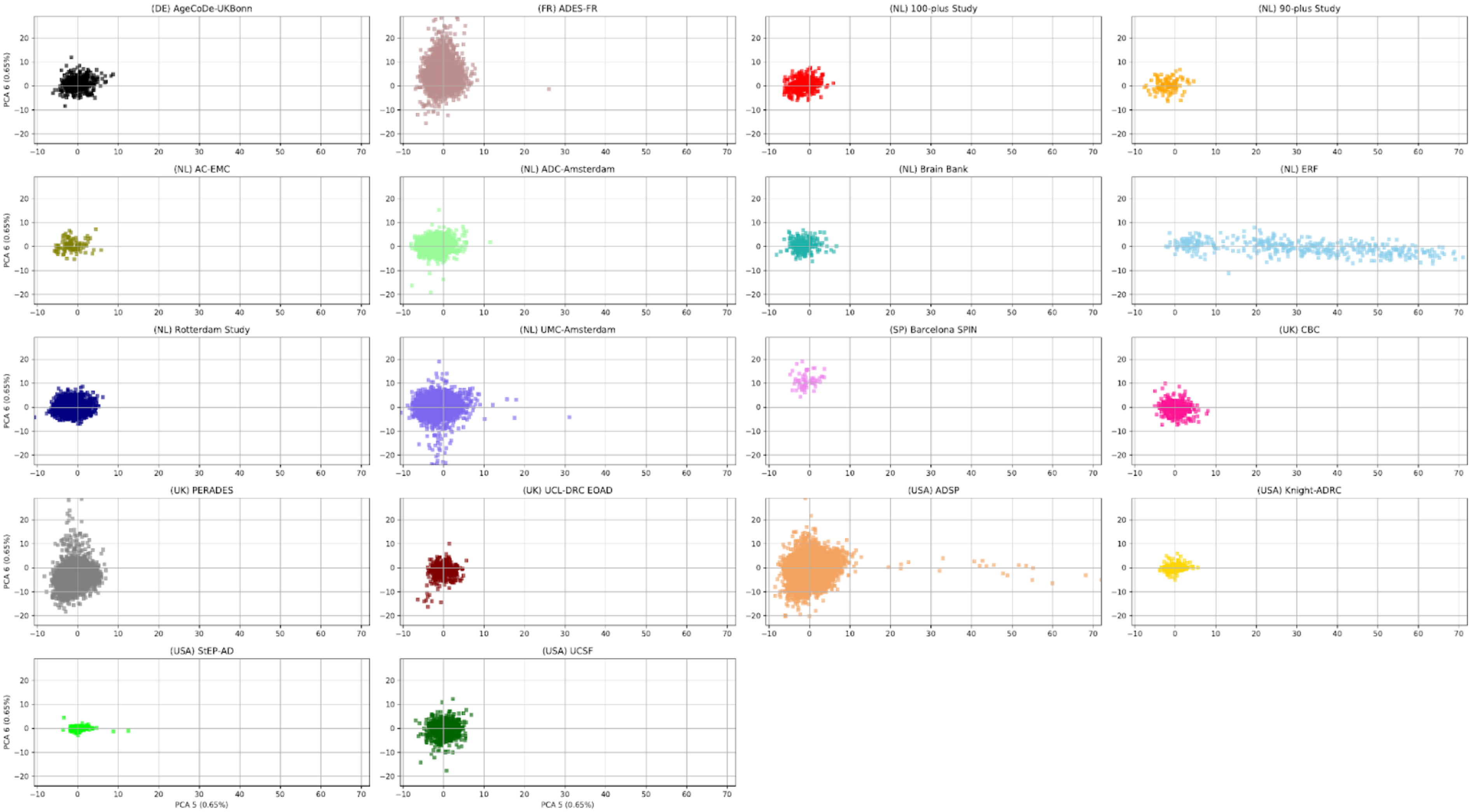

Fifth and sixth PCA covariates after sample QC. All analyses are corrected for the first 6 PCA components.

##### 2.1.19 Figure S19: Inflation in Stage-2

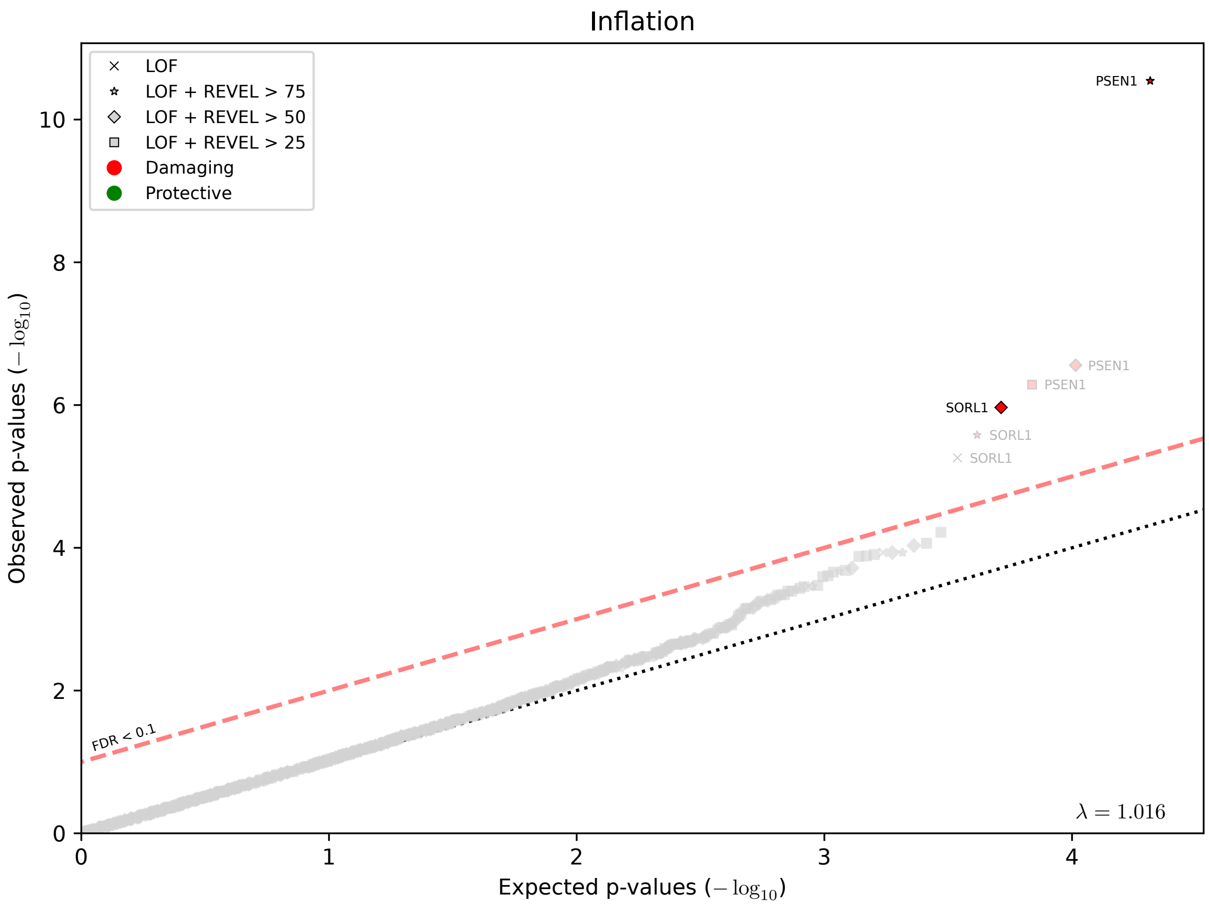

Inflation in Stage-2 (without exome-extract samples). Note that causative mutations were not separately removed in Stage-2, as we focused on a specific set of genes.

#### 2.1.20 Figure S20: Inflation in the mega-analysis dataset

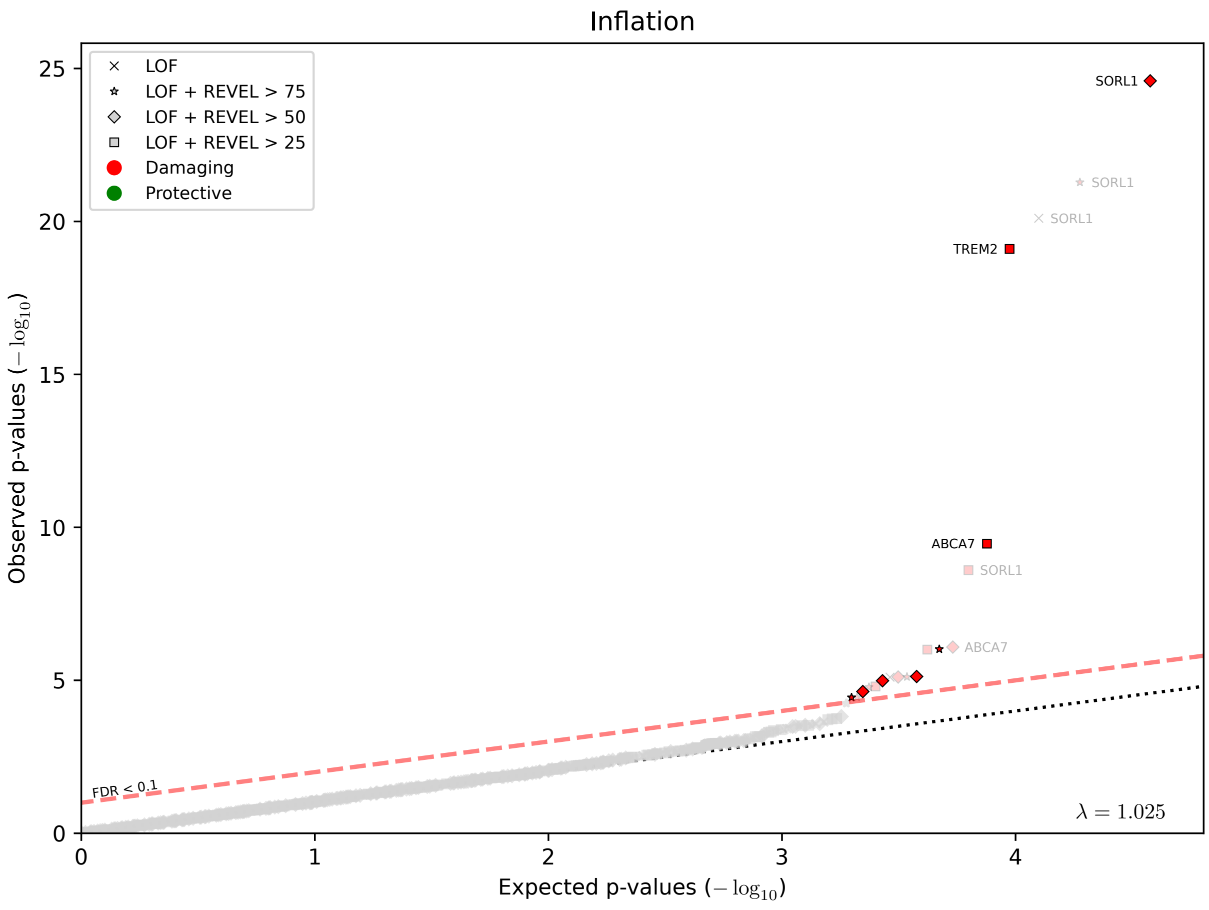

Inflation in the mega-analysis dataset (without exome-extract samples).

#### 2.1.21 Figure S21: Frequency in controls by age last seen

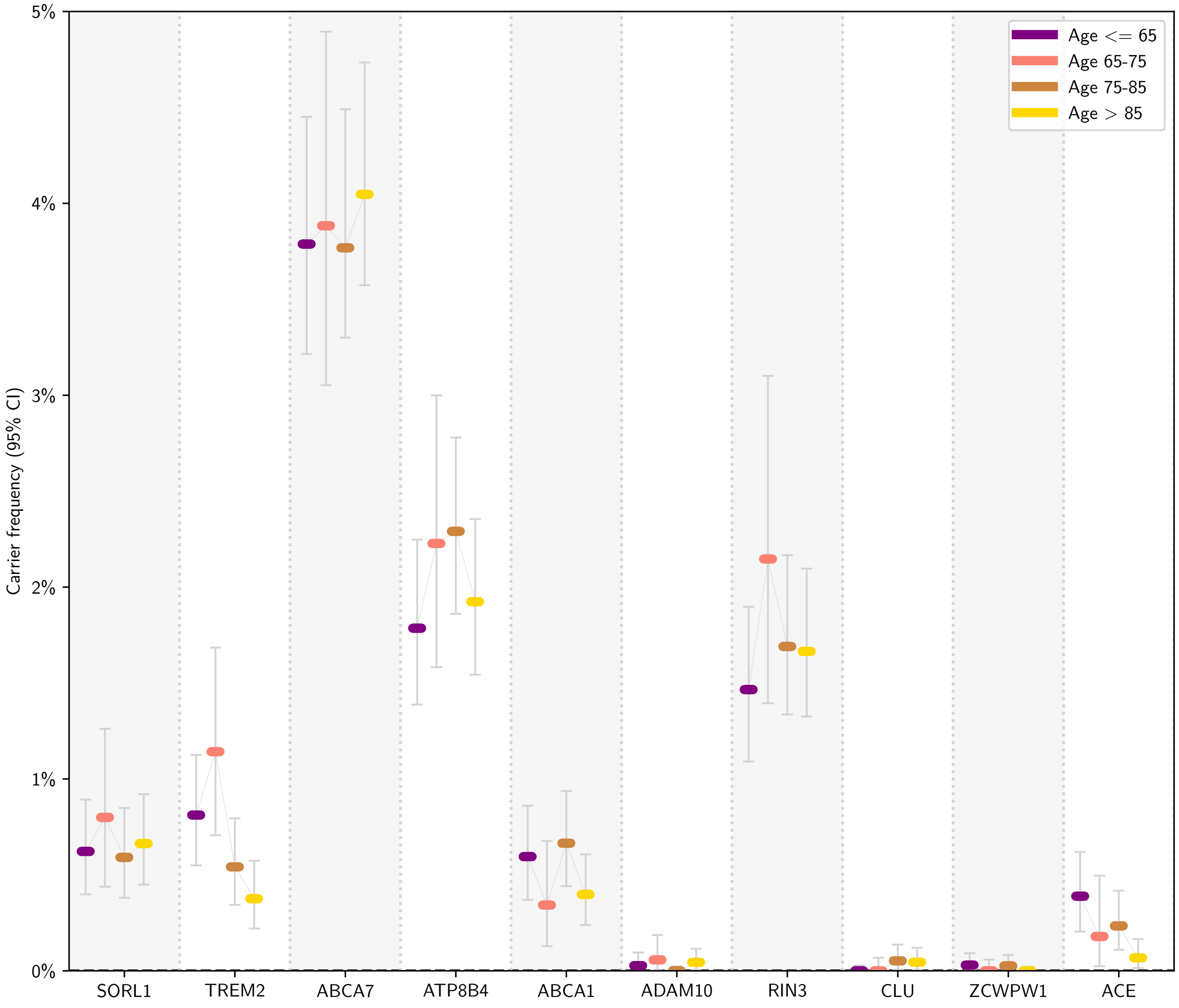

Carrier frequency in controls in the mega-analysis, by age last seen, for the variant selection threshold with the strongest association (**Table 3**, refined).

#### 2.1.22 Figure S22: Age-at-onset by variant category

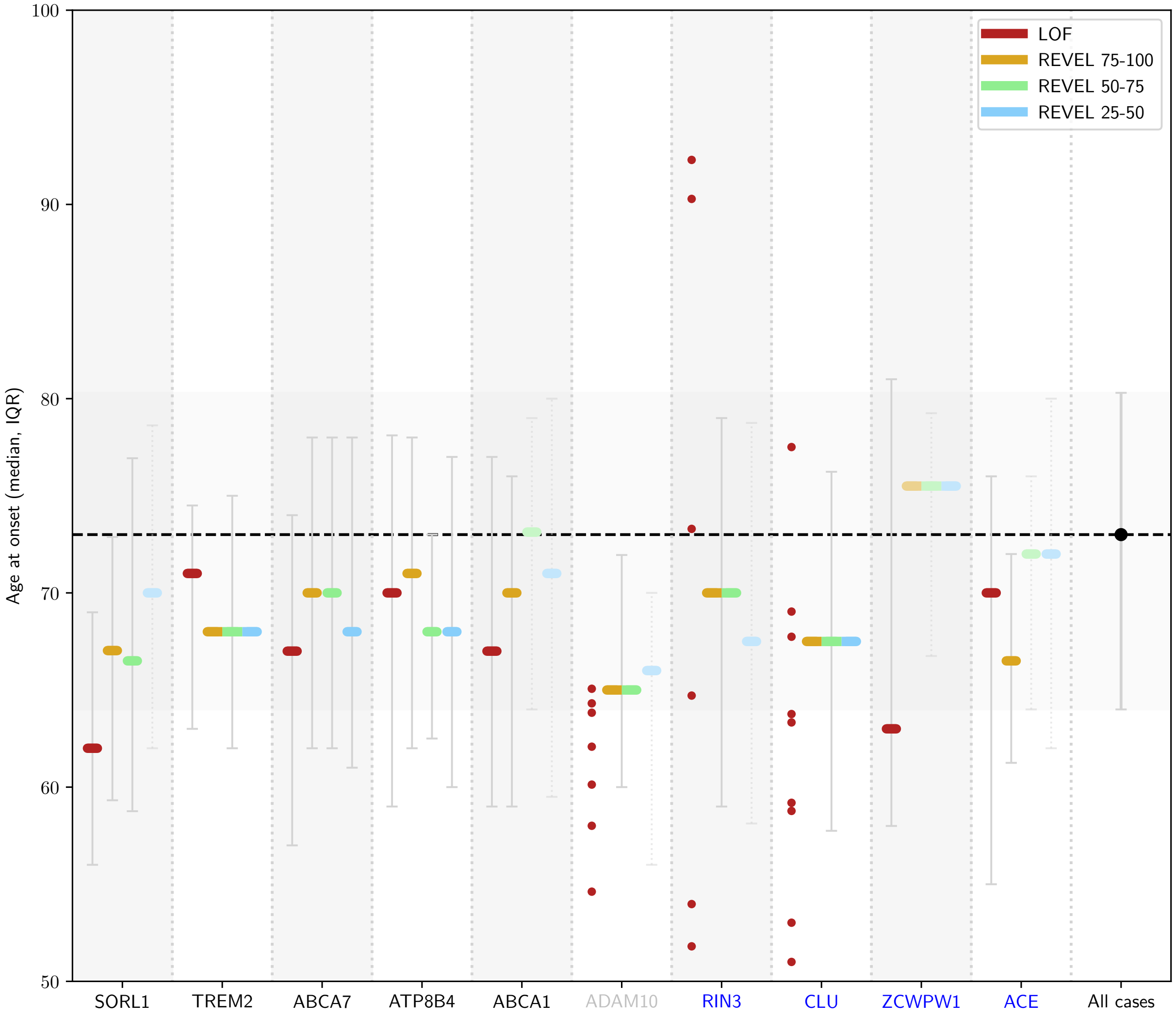

Age-at-onset (median and IQR). Samples in variant categories with < 10 samples are shown individually. The median age at onset and IQR for the complete dataset is shown on the right.

#### 2.1.23 Figure S23: Attributable fraction per gene and age-at-onset category

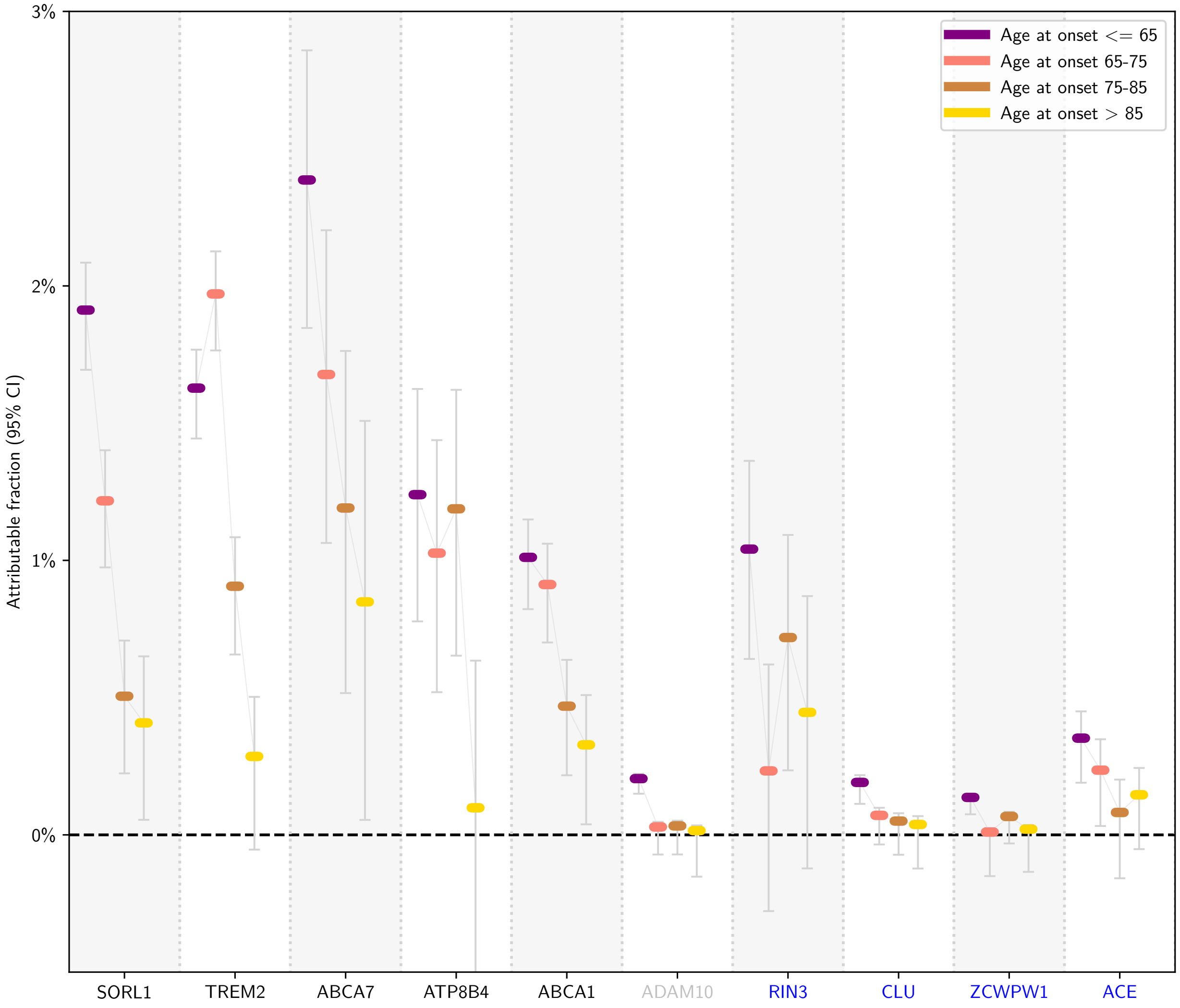

The attributable fraction of a gene is an estimate of the fraction of AD cases in a specific age group that have become part of this dataset due to carrying a rare damaging variant in that gene (Online methods). This estimate accounts only for variants in the burden selection.

#### 2.2 Supplemental Tables

#### 2.2.1 Table S1: Contributing Studies

| Study | Samples | | | | Gender (%female) | | APOE genotype (% E4) | | WGS (%) | | Case/Control Stage-1 | | | | | | Case/Control Stage-2 | | | | | | Diagnostic validation | | |
|---|---|---|---|---|---|---|---|---|---|---|---|---|---|---|---|---|---|---|---|---|---|---|---|---|---|
| | before QC (#) | after QC (#) | | | | | | | | | EOAD | | LOAD | | Controls | | EOAD | | LOAD | | Controls | | Neuro-patho-logy | CSF | Clinical |
| Study | Stage-1+2 | Stage-1+2 | Stage-1 | Stage-2 | Case | Control | Case | Control | Case | Control | # | AAO | # | AAO | # | ALS | # | AAO | # | AAO | # | ALS | | | |
| **France** | | | | | | | | | | | | | | | | | | | | | | | | | |
| ADES-FR | 4738 | 4645 | 3254 | 1391 | 62% | 48% | 53% | 22% | 30% | 9% | 1068 | 59.0 | 930 | 78.2 | 1256 | 75.5 | 477 | 57.1 | 52 | 69.1 | 862 | 61.9 | 16 | 1152 | 3477 |
| **Germany** | | | | | | | | | | | | | | | | | | | | | | | | | |
| AgeCoDe-UKBonn | 394 | 371 | 371 | 0 | 68% | - | 41% | - | 0% | 0% | 98 | 59.0 | 272 | 84.7 | 1 | - | | | | | | | 0 | 0 | 371 |
| **Spain** | | | | | | | | | | | | | | | | | | | | | | | | | |
| Barcelona SPIN | 60 | 59 | 59 | 0 | 44% | 44% | 6% | 33% | 0% | 0% | 50 | 56.4 | | | 9 | 72.8 | | | | | | | 37 | 13 | 9 |
| **The Netherlands** | | | | | | | | | | | | | | | | | | | | | | | | | |
| AC-EMC | 125 | 110 | 70 | 40 | 60% | - | 44% | - | 0% | - | 57 | 57.0 | 13 | 69.1 | | | 29 | 57.3 | 11 | 69.3 | | | 3 | 40 | 67 |
| ERF | 1325 | 400 | 400 | 0 | 50% | 57% | 75% | 31% | 0% | 0% | 1 | - | 3 | 76.0 | 396 | 48.1 | | | | | | | 0 | 0 | 400 |
| Rotterdam Study | 2699 | 1891 | 1891 | 0 | 69% | 55% | 44% | 26% | 0% | 0% | 1 | - | 366 | 83.5 | 1524 | 82.7 | | | | | | | 0 | 0 | 1891 |
| ADC-Amsterdam | 1564 | 1073 | 483 | 590 | 55% | 35% | 58% | 32% | 0% | 0% | 341 | 57.3 | 142 | 68.6 | | | 158 | 57.6 | 129 | 71.9 | 303 | 58.4 | 0 | 892 | 181 |
| Netherlands Brain Bank | 251 | 223 | 0 | 223 | 70% | 57% | 41% | 26% | 0% | 0% | | | | | | | 51 | 57.2 | 119 | 80.0 | 53 | 82.7 | 223 | 0 | 0 |
| UMC-Amsterdam | 6930 | 4299 | 0 | 4299 | 53% | 37% | - | - | 0% | 0% | | | | | | | 123 | 57.4 | 29 | 69.3 | 4147 | 45.3 | 0 | 0 | 4299 |
| 100-plus Study | 375 | 349 | 254 | 95 | 84% | 69% | 14% | 14% | 0% | 0% | | | 64 | 101.5 | 190 | 102.9 | | | | | 95 | 100.4 | 0 | 0 | 349 |
| 90-plus Study | 103 | 71 | 0 | 71 | - | 54% | - | 13% | - | 0% | | | | | | | | | | | 71 | 92.3 | 0 | 0 | 71 |
| **United Kingdom** | | | | | | | | | | | | | | | | | | | | | | | | | |
| CBC | 471 | 363 | 363 | 0 | 54% | 40% | 62% | 34% | 0% | 0% | 33 | 60.1 | 78 | 76.8 | 252 | 75.8 | | | | | | | 363 | 0 | 0 |
| PERADES | 4936 | 4140 | 4140 | 0 | 58% | 58% | 54% | 22% | 0% | 0% | 1265 | 58.1 | 2185 | 76.9 | 690 | 81.5 | | | | | | | 0 | 0 | 4140 |
| UCL-DRC EOAD | 539 | 409 | 409 | 0 | 55% | - | 47% | - | 0% | - | 389 | 54.9 | 20 | 76.6 | | | | | | | | | 7 | 35 | 367 |
| ***Europe total*** | | | | | | | | | | | | | | | | | | | | | | | | | |
| *ADES* | *24510* | *18403* | *11694* | *6709* | *60%* | *52%* | *53%* | *25%* | *9%* | *2%* | *3303* | *57.4* | *4073* | *79.0* | *4318* | *77.5* | *838* | *57.2* | *340* | *74.0* | *5531* | *53.5* | *649* | *2132* | *15622* |
| **USA** | | | | | | | | | | | | | | | | | | | | | | | | | |
| ADSP | 25798 | 12557 | 9651 | 2906 | 57% | 58% | 47% | 17% | 11% | 10% | 757 | 62.4 | 4519 | 77.2 | 4375 | 86.5 | 189 | 61.3 | 992 | 78.8 | 1725 | 78.0 | 0 | 0 | 12557 |
| StEP-AD | 278 | 278 | 0 | 278 | 50% | 58% | 12% | 89% | *exome-extract* | | | | | | | | 171 | 57.2 | 2 | 67.5 | 105 | 79.6 | 0 | 0 | 278 |
| Knight-ADRC | 1039 | 1038 | 0 | 1038 | 53% | 56% | 69% | 38% | *exome-extract* | | | | | | | | 275 | 59.3 | 383 | 75.8 | 380 | 77.5 | 0 | 0 | 1038 |
| UCSF | 736 | 282 | 0 | 282 | 52% | 63% | 58% | 24% | 100% | 100% | | | | | | | 154 | 50.6 | 40 | | 88 | 69.5 | 0 | 0 | 282 |
| ***USA total*** | | | | | | | | | | | | | | | | | | | | | | | | | |
| *USA* | *27851* | *14155* | *9651* | *4504* | *56%* | *58%* | *48%* | *20%* | *12%* | *11%* | *757* | *62.4* | *4519* | *77.2* | *4375* | *86.5* | *789* | *59.9* | *1417* | *78.0* | *2298* | *77.6* | *0* | *0* | *14155* |
| **Total** | **52361** | **32558** | **21345** | **11213** | **58%** | **55%** | **51%** | **22%** | **10%** | **5%** | **4060** | **58.8** | **8592** | **77.9** | **8693** | **82.1** | **1627** | **58.1** | **1757** | **77.2** | **7829** | **62.2** | **649** | **2132** | **29777** |

Characteristics of the samples contributed by each study, grouped by country. A.A.O: mean age at onset; A.L.S. mean age at last screening. EOAD: early onset cases, a.a.o. ≤65. LOAD, a.a.o >65. Study descriptions can be found in section 1 of the Supplement.

##### 2.2.2 Table S2: Capture Kits

| Study | Capture kits (#samples, after QC) |
|---|---|
| AgeCoDe-UKBonn | Nimblegen V2: 371 |
| ADES-FR | Agilent V1: 6, Agilent V3: 10, Agilent V4: 119, Agilent V4UTR: 14, Agilent V5: 1362, Agilent V5UTR: 849, Agilent V6UTR: 469, WGS: 954 |
| 100-plus Study | Agilent V6: 135, Nimblegen V3: 214 |
| 90-plus Study | Agilent V6: 71 |
| AC-EMC | Agilent V6: 40, Nimblegen v2: 70 |
| ADC-Amsterdam | Agilent V6: 770, Nimblegen v3: 303 |
| Brain Bank | Agilent V6: 223 |
| ERF | Agilent V4: 400 |
| Rotterdam Study | Nimblegen v2: 1891 |
| UMC-Amsterdam | MedExome: 4299 |
| Barcelona SPIN | Nimblegen v3: 59 |
| CBC | Nimblegen V2: 63, Multiplex Illumina TruSeq v2: 100, Multiplex Illumina TruSeq: 200 |
| PERADES | Nextera v1.2: 4140 |
| UCL-DRC EOAD | Sureselect: 5, Haloplex: 404 |
| ADSP | Illumina Rapid Capture Exome: 4211, Agilent V4: 16, Agilent V5: 208, Agilent V6: 6, Nimblegen VCRome V21: 6077, Nimblegen v2: 9, Nimblegen v3: 786, WGS: 1244 |
| UCSF/NYGC/UAB | WGS: 282 |

WGS: Whole Genome Sequencing

#### 2.2.3 Table S3 Sample QC and Variant QC

##### 2.2.3.1 Table S3a: Sample QC

| **Samples** | **Stage-1** | | **Stage-2** | | **Mega** | |
|---|---|---|---|---|---|---|
| QC-steps | Total | Removed | Total | Removed | Total | Removed |
| 0. Samples processed | 25,982 | | 26,379 | | 52,361 | |
| 1. Missingness | 25,857 | 125 | 26,374 | 5 | 52,231 | 130 |
| 2. Contamination | 25,430 | 427 | 26,345 | 29 | 51,775 | 456 |
| 3. Sex-check | 25,244 | 186 | 26,330 | 15 | 51,574 | 201 |
| 4. Population outliers | 24,405 | 839 | 15,563 | 10,767 | 39,968 | 11,606 |
| 5. excess novel SNPs | 24,248 | 157 | 15,469 | 94 | 39,717 | 251 |
| 6. excess novel Indels | 24,227 | 21 | 15,469 | 0 | 39,696 | 21 |
| 7. other QC | 24,212 | 15 | 15,462 | 7 | 39,674 | 22 |
| 8a. IBD | 22,334 | 1,878 | 13,859 | 1,603 | 35,689 | 3,985 |
| 8b. Duplicate w.r.t. Stage-1 | 22,334 | NA | 13,637 | 222 | 35,689 | NA |
| 9. Bad plates | 22,213 | 121 | 13,637 | NA | 35,568 | 121 |
| 10. Causative mutations | 22,047 | 166 | 13,637 | NA | 35,402 | 166 |
| 11. Braak mismatch/unlabeled | 21,345 | 702 | 11,213 | 2,424 | 31,905 | 3,497 |
| **Sample totals** | | | | | | |
| EOAD | 4,060 | | 1,627 | | 5,643 | |
| LOAD | 8,592 | | 1,757 | | 10,165 | |
| Controls | 8,693 | | 7,829 | | 16,097 | |
| **Totals excluding exome-extracts:** | | | | | | |
| EOAD | 4,060 | | 1,181 | | 5,197 | |
| LOAD | 8,592 | | 1,372 | | 9,780 | |
| Controls | 8,693 | | 6,482 | | 14,750 | |

##### 2.2.3.2 Table S3b: Variant QC (excluding exome-extract samples, all genes)

| Variants | Stage-1 | | Stage-2 | | Mega | |
|---|---|---|---|---|---|---|
| QC-steps | Total | Ratio | Total | Ratio | Total | Ratio |
| 0. Variants called | 11,752,148 | 100.0% | 7,673,870 | 100.0% | 15,172,697 | 100.0% |
| 1. Bi-allelic variants | 12,938,556 | 110.1% | 8,223,193 | 107.2% | 16,829,185 | 110.9% |
| 2. Variant merging (in-phase variants, multi-allelic overlap, low REF AF) | 12,309,375 | 104.7% | 7,555,341 | 98.5% | 15,688,759 | 103.4% |
| 3. Oxo-G mutations | 10,408,894 | 88.6% | 6,966,627 | 90.8% | 13,621,311 | 89.8% |
| 4. STR/LCR regions | 9,590,204 | 81.6% | 6,243,771 | 81.4% | 12,576,698 | 82.9% |
| 5. Allele balance (het. 0.25-0.75, hom. > 0.9) | 8,012,587 | 68.2% | 5,318,752 | 69.3% | 10,293,141 | 67.8% |
| 6. Depth fraction heterozygous calls > 0.2 | 7,814,724 | 66.5% | 5,227,137 | 68.1% | 9,909,839 | 65.3% |
| 7. Hardy-Weinberg (p < 5e-8) | 7,779,331 | 66.2% | 5,178,054 | 67.5% | 9,857,042 | 65.0% |
| 8. VQSR | 7,612,856 | 64.8% | 5,084,524 | 66.3% | 9,655,998 | 63.6% |
| 9. Variant Batch Detector | 7,543,193 | 64.2% | 4,908,915 | 64.0% | 9,454,876 | 62.3% |
| **Variant selection** | | | | | | |
| 10. In protein coding autosomal genes (Gencode V19/V29) | 6,883,630 | 58.6% | 4,358,607 | 56.8% | 8,618,616 | 56.8% |
| M.11. Missense variants | 1,486,559 | 12.6% | 764,058 | 10.0% | 1,789,034 | 11.8% |
| M.12. REVEL > 25 | 540,934 | 4.6% | 269,492 | 3.5% | 658,387 | 4.3% |
| M.13 (GnomAD) MAF < 1% / dosage > 0.5 | 530,072 | 4.5% | 250,029 | 3.3% | 643,920 | 4.2% |
| M.14. Missingness (< 20% + no differential missingness) | 353,913 | 3.0% | 182,533 | 2.4% | 470,563 | 3.1% |
| L.11. Loss-of-function variants (stop-gained,frameshift,splice acceptor/donor) | 144,429 | 1.2% | 65,358 | 0.9% | 165,516 | 1.1% |
| L.12. Loftee HC + VEP high impact | 109,550 | 0.9% | 49,425 | 0.6% | 125,766 | 0.8% |
| L.13. (GnomAD) MAF < 1% / dosage > 0.5 | 108,016 | 0.9% | 45,644 | 0.6% | 123,514 | 0.8% |
| L.14. Missingness(< 20% + no differential missingness) | 57,543 | 0.5% | 27,191 | 0.4% | 74,645 | 0.5% |
| **Categories** | | | | | | |
| REVEL 25-50 | 198,068 | 1.7% | 102,866 | 1.3% | 262,244 | 1.7% |
| REVEL 50-75 | 99,910 | 0.9% | 51,295 | 0.7% | 133,059 | 0.9% |
| REVEL 75-100 | 54,212 | 0.5% | 27,487 | 0.4% | 72,832 | 0.5% |
| LOF | 57,543 | 0.5% | 27,191 | 0.4% | 74,645 | 0.5% |
| **Thresholds** | | | | | | |
| LOF+REVEL≥25 | 409,733 | 3.5% | 208,839 | 2.7% | 542,780 | 3.6% |
| LOF+REVEL≥50 | 211,665 | 1.8% | 105,973 | 1.4% | 280,536 | 1.8% |
| LOF+REVEL≥75 | 111,755 | 1.0% | 54,678 | 0.7% | 147,477 | 1.0% |
| LOF | 57,543 | 0.5% | 27,191 | 0.4% | 74,645 | 0.5% |

##### 2.2.3.3 Table S3c: Variant QC: including exome-extract samples, only targeted genes

| Variants in targeted genes | Stage-1 | Stage-2 | Mega |
|---|---|---|---|

| QC-steps | Total | Ratio | Total | Ratio | Total | Ratio |
|---|---|---|---|---|---|---|
| 0. Variants called | 10,339 | 100.0% | 7,238 | 100.0% | 13,032 | 100.0% |
| 1. Bi-allelic variants | 11,384 | 110.1% | 7,803 | 107.8% | 14,531 | 111.5% |
| 2. Variant merging (in-phase variants, multi-allelic overlap, low REF AF) | 10,674 | 103.2% | 7,253 | 100.2% | 13,652 | 104.8% |
| 3. Oxo-G mutations | 9,075 | 87.8% | 6,705 | 92.6% | 11,948 | 91.7% |
| 4. STR/LCR regions | 8,443 | 81.7% | 6,034 | 83.4% | 11,115 | 85.3% |
| 5. Allele balance (het. 0.25-0.75, hom. > 0.9) | 7,207 | 69.7% | 5,282 | 73.0% | 9,453 | 72.5% |
| 6. Depth fraction heterozygous calls > 0.2 | 6,958 | 67.3% | 5,180 | 71.6% | 9,129 | 70.1% |
| 7. Hardy-Weinberg (p < 5e-8) | 6,941 | 67.1% | 5,164 | 71.3% | 9,112 | 69.9% |
| 8. VQSR | 6,851 | 66.3% | 5,075 | 71.3% | 8,966 | 68.8% |
| 9. Variant Batch Detector | 6,848 | 66.2% | 5,072 | 70.1% | 8,963 | 68.8% |
| **Variant selection** | | | | | | |
| 10. In protein coding autosomal genes (Gencode V19/V29) | 6,848 | 66.2% | 5,072 | 70.1% | 8,963 | 68.8% |
| M.11. Missense variants | 1,590 | 15.4% | 894 | 12.4% | 1,873 | 14.4% |
| M.12. REVEL > 25 | 1,066 | 10.3% | 591 | 8.2% | 1,263 | 9.7% |
| M.13 (gnomAD) MAF < 1% / dosage > 0.5 | 1,022 | 9.9% | 567 | 7.8% | 1,228 | 9.4% |
| M.14. Missingness (< 20% + no differential missingness) | 781 | 7.6% | 428 | 5.9% | 943 | 7.2% |
| L.11. Loss-of-function variants (stop-gained,frameshift,splice acceptor/donor) | 206 | 2.0% | 106 | 1.5% | 255 | 2.0% |
| L.12. Loftee HC + VEP high impact | 191 | 1.8% | 97 | 1.3% | 236 | 1.8% |
| L.13. (gnomAD) MAF < 1% / dosage > 0.5 | 189 | 1.8% | 96 | 1.3% | 234 | 1.8% |
| L.14. Missingness (< 20% + no differential missingness) | 136 | 1.3% | 64 | 0.9% | 168 | 1.3% |
| **Categories** | | | | | | |
| REVEL 25-50 | 296 | 2.9% | 168 | 2.3% | 360 | 2.8% |
| REVEL 50-75 | 266 | 2.6% | 140 | 1.9% | 320 | 2.5% |
| REVEL 75-100 | 214 | 2.1% | 119 | 1.6% | 258 | 2.0% |
| LOF | 136 | 1.3% | 64 | 0.9% | 168 | 1.3% |
| **Thresholds** | | | | | | |
| LOF+REVEL≥25 | 912 | 8.8% | 491 | 6.8% | 1,106 | 8.5% |
| LOF+REVEL≥50 | 616 | 6.0% | 323 | 4.5% | 746 | 5.7% |
| LOF+REVEL≥75 | 350 | 3.4% | 183 | 2.5% | 426 | 3.3% |
| LOF | 136 | 1.3% | 64 | 0.9% | 168 | 1.3% |

#### 2.2.4 Table S4: Power in stage-1

| | | ncarriers = 10 | | | | | | ncarriers = 25 | | | | | | ncarriers = 50 | | | | | |
|---|---|---|---|---|---|---|---|---|---|---|---|---|---|---|---|---|---|---|---|
| | | | | P < value | | | | | | P < value | | | | | | P < value | | | |
| **Odds ratio** | **Test** | **P-50%** | **P-80%** | **1E-04** | **1E-05** | **1E-06** | **1E-07** | **P-50%** | **P-80%** | **1E-04** | **1E-05** | **1E-06** | **1E-07** | **P-50%** | **P-80%** | **1E-04** | **1E-05** | **1E-06** | **1E-07** |
| EOAD: 1.5<br>LOAD: 1.3 | Case - Control | 0.495 | 0.751 | 0% | 0% | 0% | 0% | 0.426 | 0.760 | 0% | 0% | 0% | 0% | 0.285 | 0.699 | 0% | 0% | 0% | 0% |
| | EOAD vs. rest | 0.481 | 0.769 | 0% | 0% | 0% | 0% | 0.406 | 0.759 | 0% | 0% | 0% | 0% | 0.390 | 0.765 | 0% | 0% | 0% | 0% |
| | Ordinal | 0.488 | 0.792 | 0% | 0% | 0% | 0% | 0.435 | 0.807 | 0% | 0% | 0% | 0% | 0.264 | 0.654 | 0% | 0% | 0% | 0% |
| EOAD: 2.5<br>LOAD: 1.5 | Case - Control | 0.352 | 0.757 | 0% | 0% | 0% | 0% | 0.176 | 0.597 | 0% | 0% | 0% | 0% | 0.055 | 0.284 | 2% | 0% | 0% | 0% |
| | EOAD vs. rest | 0.310 | 0.689 | 0% | 0% | 0% | 0% | 0.120 | 0.435 | 2% | 1% | 0% | 0% | 0.045 | 0.284 | 4% | 0% | 0% | 0% |
| | Ordinal | 0.338 | 0.693 | 0% | 0% | 0% | 0% | 0.074 | 0.416 | 2% | 1% | 0% | 0% | 0.015 | 0.098 | 7% | 3% | 0% | 0% |
| EOAD: 3.5<br>LOAD: 2.0 | Case - Control | 0.264 | 0.672 | 0% | 0% | 0% | 0% | 0.065 | 0.309 | 1% | 0% | 0% | 0% | 0.008 | 0.049 | 10% | 3% | 1% | 0% |
| | EOAD vs. rest | 0.176 | 0.608 | 0% | 0% | 0% | 0% | 0.066 | 0.335 | 3% | 1% | 0% | 0% | 0.008 | 0.084 | 15% | 5% | 2% | 0% |
| | Ordinal | 0.142 | 0.503 | 1% | 0% | 0% | 0% | 0.020 | 0.175 | 5% | 1% | 0% | 0% | 0.001 | 0.020 | 29% | 12% | 3% | 1% |
| EOAD: 5.0<br>LOAD: 2.5 | Case - Control | 0.184 | 0.492 | 0% | 0% | 0% | 0% | 0.025 | 0.103 | 5% | 0% | 0% | 0% | 0.001 | 0.008 | 31% | 13% | 4% | 1% |
| | EOAD vs. rest | 0.113 | 0.372 | 1% | 0% | 0% | 0% | 0.026 | 0.135 | 6% | 2% | 1% | 1% | 0.001 | 0.014 | 32% | 16% | 7% | 5% |
| | Ordinal | 0.073 | 0.357 | 2% | 0% | 0% | 0% | 0.006 | 0.042 | 13% | 4% | 1% | 0% | 0.000 | 0.001 | 58% | 39% | 21% | 12% |
| EOAD: 10.0<br>LOAD: 3.33 | Case - Control | 0.085 | 0.223 | 0% | 0% | 0% | 0% | 0.001 | 0.017 | 18% | 4% | 0% | 0% | 0.000 | 0.000 | 73% | 48% | 26% | 13% |
| | EOAD vs. rest | 0.026 | 0.139 | 9% | 2% | 0% | 0% | 0.000 | 0.004 | 44% | 22% | 11% | 6% | 0.000 | 0.000 | **90%** | 72% | 53% | 40% |
| | Ordinal | 0.010 | 0.082 | 8% | 2% | 1% | 0% | 0.000 | 0.001 | 61% | 35% | 19% | 8% | 0.000 | 0.000 | **98%** | **95%** | **83%** | 67% |
| EOAD: 20.0<br>LOAD: 5.0 | Case - Control | 0.024 | 0.138 | 0% | 0% | 0% | 0% | 0.000 | 0.002 | 39% | 15% | 2% | 0% | 0.000 | 0.000 | **97%** | **85%** | 69% | 46% |
| | EOAD vs. rest | 0.005 | 0.044 | 19% | 7% | 1% | 1% | 0.000 | 0.000 | 72% | 55% | 36% | 23% | 0.000 | 0.000 | **100%** | **98%** | **91%** | **86%** |
| | Ordinal | 0.001 | 0.013 | 20% | 8% | 7% | 1% | 0.000 | 0.000 | **90%** | 73% | 56% | 35% | 0.000 | 0.000 | **100%** | **100%** | **99%** | **97%** |
| EOAD: 40.0<br>LOAD: 10.0 | Case - Control | 0.012 | 0.078 | 0% | 0% | 0% | 0% | 0.000 | 0.000 | 67% | 32% | 4% | 0% | 0.000 | 0.000 | **100%** | **99%** | **93%** | **84%** |
| | EOAD vs. rest | 0.005 | 0.032 | 22% | 10% | 1% | 1% | 0.000 | 0.000 | 74% | 61% | 46% | 35% | 0.000 | 0.000 | **99%** | **99%** | **95%** | **92%** |
| | Ordinal | 0.001 | 0.006 | 24% | 7% | 6% | 1% | 0.000 | 0.000 | **96%** | **81%** | 69% | 51% | 0.000 | 0.000 | **100%** | **100%** | **100%** | **100%** |

| | | genes - LOF | | | | genes - LOF+REVEL>=75 | | | | genes - LOF+REVEL>=50 | | | | genes - LOF+REVEL>=25 | | | |
|---|---|---|---|---|---|---|---|---|---|---|---|---|---|---|---|---|---|
| | | P < value | | | | P < value | | | | P < value | | | | P < value | | | |
| **Odds ratio** | **Test** | **1E-04** | **1E-05** | **1E-06** | **1E-07** | **1E-04** | **1E-05** | **1E-06** | **1E-07** | **1E-04** | **1E-05** | **1E-06** | **1E-07** | **1E-04** | **1E-05** | **1E-06** | **1E-07** |
| EOAD: 1.5<br>LOAD: 1.3 | Case - Control | 0% | 0% | 0% | 0% | 0% | 0% | 0% | 0% | 1% | 0% | 0% | 0% | 2% | 1% | 0% | 0% |
| | EOAD vs. rest | 0% | 0% | 0% | 0% | 0% | 0% | 0% | 0% | 0% | 0% | 0% | 0% | 1% | 0% | 0% | 0% |
| | Ordinal | 0% | 0% | 0% | 0% | 0% | 0% | 0% | 0% | 1% | 0% | 0% | 0% | 2% | 1% | 1% | 0% |
| EOAD: 2.5<br>LOAD: 1.5 | Case - Control | 1% | 0% | 0% | 0% | 2% | 1% | 1% | 0% | 5% | 3% | 2% | 2% | 11% | 8% | 6% | 4% |
| | EOAD vs. rest | 1% | 1% | 0% | 0% | 3% | 2% | 1% | 1% | 6% | 5% | 3% | 2% | 14% | 11% | 8% | 6% |
| | Ordinal | 1% | 1% | 1% | 0% | 3% | 2% | 2% | 1% | 8% | 6% | 5% | 4% | 17% | 14% | 11% | 9% |
| EOAD: 3.5<br>LOAD: 2.0 | Case - Control | 2% | 1% | 1% | 1% | 4% | 3% | 2% | 2% | 10% | 8% | 6% | 5% | 22% | 17% | 14% | 12% |
| | EOAD vs. rest | 2% | 1% | 1% | 1% | 4% | 3% | 2% | 2% | 10% | 7% | 6% | 5% | 21% | 16% | 13% | 11% |
| | Ordinal | 2% | 2% | 1% | 1% | 6% | 5% | 4% | 3% | 14% | 11% | 9% | 7% | 28% | 23% | 19% | 16% |
| EOAD: 5.0<br>LOAD: 2.5 | Case - Control | 2% | 2% | 1% | 1% | 6% | 5% | 4% | 3% | 14% | 12% | 9% | 8% | 29% | 24% | 21% | 18% |
| | EOAD vs. rest | 3% | 2% | 1% | 1% | 7% | 5% | 4% | 3% | 15% | 12% | 9% | 8% | 29% | 24% | 20% | 17% |
| EOAD: 10.0<br>LOAD: 3.33 | Case - Control | 4% | 3% | 2% | 2% | 10% | 8% | 6% | 5% | 21% | 17% | 14% | 13% | 39% | 34% | 30% | 26% |
| | EOAD vs. rest | 6% | 5% | 4% | 3% | 14% | 11% | 9% | 7% | 27% | 22% | 19% | 16% | 46% | 41% | 36% | 32% |
| | Ordinal | 7% | 6% | 4% | 4% | 16% | 13% | 11% | 9% | 30% | 25% | 22% | 19% | 50% | 45% | 41% | 37% |
| EOAD: 20.0<br>LOAD: 5.0 | Case - Control | 5% | 4% | 3% | 3% | 13% | 10% | 8% | 7% | 26% | 22% | 18% | 16% | 46% | 41% | 36% | 33% |
| | EOAD vs. rest | 9% | 7% | 6% | 5% | 19% | 16% | 13% | 11% | 34% | 30% | 26% | 23% | 55% | 50% | 46% | 42% |
| | Ordinal | 11% | 8% | 7% | 6% | 21% | 17% | 15% | 13% | 37% | 32% | 29% | 26% | 58% | 53% | 49% | 46% |
| EOAD: 40.0<br>LOAD: 10.0 | Case - Control | 7% | 5% | 4% | 3% | 15% | 12% | 10% | 9% | 29% | 25% | 21% | 19% | 50% | 45% | 41% | 37% |
| | EOAD vs. rest | 10% | 8% | 6% | 5% | 20% | 17% | 14% | 12% | 35% | 31% | 27% | 24% | 56% | 52% | 47% | 44% |
| | Ordinal | 12% | 9% | 8% | 6% | 23% | 19% | 16% | 14% | 39% | 34% | 31% | 28% | 60% | 55% | 52% | 48% |

Power calculations were performed for ordinal logistic regression and Firth logistic regression (case-control and EOAD vs. rest), **Figure 1B**. Given odds ratios for EOAD and LOAD cases, and the cumulative minor allele count (cMAC) per gene, we sampled the number of alleles in EOAD cases, LOAD cases and controls according to a multinomial distribution. We randomized these allele-carriers across the dataset, and performed the burden test (as described above). CMAC values per gene were set to x for the ‘ncarriers=x’ columns, while for the ‘genes’ columns values per gene were set according to the cMAC values observed in the Stage-1 dataset (**Figure 1B**) for the 4 different variant deleteriousness thresholds. Power for genes with cMAC <10 was set to 0, as these genes were not analyzed.

#### 2.2.5 Table S5: List of genes and tests performed for the targeted GWAS analysis

Only variant selection thresholds for which there were at least >= 10 damaging alleles in the dataset were considered for burden testing. **bold**=passed significance threshold; *italic*=not enough damaging alleles to perform burden testing *Note that for the CLU/PTK2B loci and the APP/ADAMTS1 loci, which are each near each other, the same genes were prioritized.*

| Tested gene: Ensemble id | gene name | description | gene prioritization source | GWAS locus name | GWAS sentinel SNPs : dbSNP id | EADB OR (95% CI) | EADB p-value | performed burden tests (allele count) |
|---|---|---|---|---|---|---|---|---|
| ENSG00000136717 | BIN1 | bridging integrator 1 [Source:HGNC Symbol;Acc:HGNC:1052] | schwarzentruber | BIN1 | rs6733839 | 1.17 (1.16-1.19) | 6.06E-118 | LOF+REVEL>=25 (147), LOF+REVEL>=50 (12) |
| ENSG00000073921 | PICALM | phosphatidylinositol binding clathrin assembly protein [Source:HGNC Symbol;Acc:HGNC:15514] | schwarzentruber | EED | rs3851179 | 0.9 (0.89-0.92) | 2.95E-48 | LOF+REVEL>=25 (49), LOF+REVEL>=50 (25) |
| ENSG00000203710 | CR1 | complement C3b/C4b receptor 1 (Knops blood group) [Source:HGNC Symbol;Acc:HGNC:2334] | schwarzentruber | CR1 | rs679515 | 1.13 (1.11-1.15) | 7.16E-46 | LOF+REVEL>=25 (322), LOF+REVEL>=50 (43), LOF+REVEL>=75 (26), LOF (22) |
| **ENSG00000120885** | **CLU** | **clusterin [Source:HGNC Symbol;Acc:HGNC:2095]** | **schwarzentruber** | CLU | rs11787077 | 0.91 (0.9-0.92) | 1.70E-44 | LOF+REVEL>=25 (26), LOF+REVEL>=50 (15), LOF+REVEL>=75 (12), LOF (10) |
| ENSG00000120899 | PTK2B | protein tyrosine kinase 2 beta [Source:HGNC Symbol;Acc:HGNC:9612] | schwarzentruber | | | | | LOF+REVEL>=25 (236), LOF+REVEL>=50 (98), LOF+REVEL>=75 (43), LOF (14) |
| *ENSG00000166926* | *MS4A6E* | *membrane spanning 4-domains A6E [Source:NCBI gene;Acc:245802]* | *schwarzentruber* | MS4A4A | rs1582763 | 0.91 (0.9-0.92) | 3.74E-42 | |
| **ENSG00000064687** | **ABCA7** | **ATP binding cassette subfamily A member 7 [Source:HGNC Symbol;Acc:HGNC:37]** | **schwarzentruber** | ABCA7 | rs12151021 | 1.1 (1.09-1.12) | 1.59E-37 | LOF+REVEL>=25 (1363), LOF+REVEL>=50 (1044), LOF+REVEL>=75 (400), LOF (115) |
| **ENSG00000124731** | **TREM1** | **triggering receptor expressed on myeloid cells 1 [Source:HGNC Symbol;Acc:HGNC:17760]** | **schwarzentruber** | TREM2 | rs10947943;rs143332484; rs75932628;rs60755019 | 0.94 (0.93-0.96);1.41 (1.32-1.5); 2.39 (2.09-2.73);1.55 (1.33-1.8) | 1.13e-09;2.78e-25; 2.53e-37;2.07e-08 | LOF+REVEL>=25 (15), LOF+REVEL>=75 (12), LOF+REVEL>=50 (12), LOF (12) |
| ENSG00000095970 | TREM2 | triggering receptor expressed on myeloid cells 2 [Source:HGNC Symbol;Acc:HGNC:17761] | schwarzentruber | | | | | LOF+REVEL>=25 (385), LOF+REVEL>=50 (53), LOF+REVEL>=75 (50), LOF (49) |
| ENSG00000085514 | PILRA | paired immunoglobin like type 2 receptor alpha | schwarzentruber | SPDYE3 | rs7384878 | 0.92 (0.91-0.94) | 1.06E-26 | LOF+REVEL>=25 (11) |

| | | | | | | | | |
|---|---|---|---|---|---|---|---|---|
| | | [Source:HGNC Symbol;Acc:HGNC:20396] | | | | | | |
| **ENSG00000078487** | **ZCWPW1** | **zinc finger CW-type and PWWP domain containing 1 [Source:HGNC Symbol;Acc:HGNC:23486]** | **schwarzentruber** | | | | | LOF+REVEL>=25 (79), LOF+REVEL>=50 (72), LOF+REVEL>=75 (16), LOF (16) |
| ENSG00000138613 | APH1B | aph-1 homolog B, gamma-secretase subunit [Source:HGNC Symbol;Acc:HGNC:24080] | schwarzentruber | APH1B | rs117618017 | 1.11 (1.09-1.13) | 2.15E-25 | LOF+REVEL>=25 (46), LOF+REVEL>=50 (36), LOF+REVEL>=75 (21), LOF (12) |
| ENSG00000120885 | CLU | clusterin [Source:HGNC Symbol;Acc:HGNC:2095] | schwarzentruber | PTK2B | rs73223431 | 1.07 (1.06-1.08) | 4.03E-22 | LOF+REVEL>=25 (26), LOF+REVEL>=50 (15), LOF+REVEL>=75 (12), LOF (10) |
| ENSG00000120899 | PTK2B | protein tyrosine kinase 2 beta [Source:HGNC Symbol;Acc:HGNC:9612] | schwarzentruber | | | | | LOF+REVEL>=25 (236), LOF+REVEL>=50 (98), LOF+REVEL>=75 (43), LOF (14) |
| ENSG00000198087 | CD2AP | CD2 associated protein [Source:HGNC Symbol;Acc:HGNC:14258] | schwarzentruber | CD2AP | rs7767350 | 1.08 (1.06-1.09) | 7.94E-22 | LOF+REVEL>=25 (51), LOF+REVEL>=50 (21), LOF+REVEL>=75 (16) |
| **ENSG00000137642** | **SORL1** | **sortilin related receptor 1 [Source:HGNC Symbol;Acc:HGNC:11185]** | **schwarzentruber** | SORL1 | rs74685827;rs11218343 | 1.19 (1.13-1.25);0.84 (0.81-0.87) | 2.81e-11;1.4e-21 | LOF+REVEL>=25 (1308), LOF+REVEL>=50 (380), LOF+REVEL>=75 (205), LOF (63) |
| ENSG00000087589 | CASS4 | Cas scaffold protein family member 4 [Source:HGNC Symbol;Acc:HGNC:15878] | schwarzentruber | CASS4 | rs6014724 | 0.89 (0.87-0.91) | 4.13E-21 | LOF+REVEL>=25 (111) |
| **ENSG00000100599** | **RIN3** | **Ras and Rab interactor 3 [Source:HGNC Symbol;Acc:HGNC:18751]** | **schwarzentruber** | SLC24A4 | rs7401792;rs12590654 | 1.04 (1.02-1.05);0.93 (0.92-0.95) | 4.83e-08;4.25e-21 | LOF+REVEL>=25 (629), LOF+REVEL>=50 (588), LOF+REVEL>=75 (10) |
| ENSG00000030582 | GRN | granulin precursor [Source:HGNC Symbol;Acc:HGNC:4601] | eadb | GRN | rs5848 | 1.07 (1.06-1.09) | 2.38E-20 | LOF+REVEL>=25 (110), LOF+REVEL>=50 (45), LOF+REVEL>=75 (24) |
| ENSG00000204287 | HLA-DRA | major histocompatibility complex, class II, DR alpha [Source:HGNC Symbol;Acc:HGNC:4947] | schwarzentruber | HLA-DQA1 | rs6605556 | 0.91 (0.9-0.93) | 7.07E-20 | LOF+REVEL>=25 (10) |
| **ENSG00000159640** | **ACE** | **angiotensin I converting enzyme [Source:HGNC Symbol;Acc:HGNC:2707]** | **schwarzentruber** | ACE | rs4277405 | 0.94 (0.93-0.95) | 8.80E-20 | LOF+REVEL>=25 (1113), LOF+REVEL>=50 (245), LOF+REVEL>=75 (101), LOF (52) |
| *ENSG00000136485* | *DCAF7* | *DDB1 and CUL4 associated factor 7 [Source:HGNC Symbol;Acc:HGNC:30915]* | *schwarzentruber* | | | | | |

| | | | | | | | | |
|---|---|---|---|---|---|---|---|---|
| ENSG00000108219 | TSPAN14 | tetraspanin 14 [Source:HGNC Symbol;Acc:HGNC:23303] | schwarzentruber | TSPAN14 | rs6586028 | 0.93 (0.91-0.94) | 1.97E-19 | LOF+REVEL>=25 (34), LOF+REVEL>=50 (12) |
| ENSG00000148429 | USP6NL | USP6 N-terminal like [Source:HGNC Symbol;Acc:HGNC:16858] | schwarzentruber | USP6NL | rs7912495 | 1.06 (1.05-1.08) | 9.74E-19 | LOF+REVEL>=25 (31) |
| ENSG00000168918 | INPP5D | inositol polyphosphate-5-phosphatase D [Source:HGNC Symbol;Acc:HGNC:6079] | schwarzentruber | INPP5D | rs10933431 | 0.93 (0.92-0.95) | 3.62E-18 | LOF+REVEL>=25 (386), LOF+REVEL>=50 (75), LOF+REVEL>=75 (20) |
| ENSG00000002587 | HS3ST1 | heparan sulfate-glucosamine 3-sulfotransferase 1 [Source:HGNC Symbol;Acc:HGNC:5194] | schwarzentruber | CLNK | rs6846529 | 1.07 (1.05-1.08) | 2.20E-17 | LOF+REVEL>=25 (31), LOF+REVEL>=50 (18) |
| ENSG00000179526 | SHARPIN | SHANK associated RH domain interactor [Source:HGNC Symbol;Acc:HGNC:25321] | eadb | SHARPIN | rs34173062 | 1.13 (1.09-1.16) | 1.72E-16 | LOF+REVEL>=25 (27), LOF+REVEL>=50 (16) |
| ENSG00000146090 | RASGEF1C | RasGEF domain family member 1C [Source:HGNC Symbol;Acc:HGNC:27400] | nearest | RASGEF1C | rs113706587 | 1.09 (1.07-1.12) | 2.22E-16 | LOF+REVEL>=25 (44) |
| ENSG00000073712 | FERMT2 | fermitin family member 2 [Source:HGNC Symbol;Acc:HGNC:15767] | schwarzentruber | FERMT2 | rs17125924 | 1.1 (1.07-1.12) | 8.32E-16 | LOF+REVEL>=25 (73), LOF+REVEL>=50 (14) |
| *ENSG00000127184* | *COX7C* | *cytochrome c oxidase subunit 7C [Source:HGNC Symbol;Acc:HGNC:2292]* | *nearest* | COX7C | rs62374257 | 1.07 (1.05-1.09) | 1.38E-15 | |
| **ENSG00000137845** | **ADAM10** | **ADAM metallopeptidase domain 10 [Source:HGNC Symbol;Acc:HGNC:188]** | **schwarzentruber** | MINDY2 | rs602602 | 0.94 (0.93-0.96) | 2.07E-15 | LOF+REVEL>=25 (45), LOF+REVEL>=50 (23), LOF+REVEL>=75 (15), LOF (10) |
| ENSG00000166035 | LIPC | lipase C, hepatic type [Source:HGNC Symbol;Acc:HGNC:6619] | schwarzentruber | | | | | LOF+REVEL>=25 (357), LOF+REVEL>=50 (156), LOF+REVEL>=75 (126), LOF (30) |
| ENSG00000126856 | PRDM7 | PR/SET domain 7 [Source:HGNC Symbol;Acc:HGNC:9351] | nearest | PRDM7 | rs56407236 | 1.11 (1.08-1.14) | 6.47E-15 | LOF+REVEL>=25 (79), LOF+REVEL>=50 (41), LOF+REVEL>=75 (41), LOF (41) |
| ENSG00000108798 | ABI3 | ABI family member 3 [Source:HGNC Symbol;Acc:HGNC:29859] | nearest | ABI3 | rs616338 | 1.32 (1.23-1.42) | 2.82E-14 | LOF+REVEL>=25 (49), LOF+REVEL>=50 (32) |
| ENSG00000146904 | EPHA1 | EPH receptor A1 [Source:HGNC Symbol;Acc:HGNC:3385] | schwarzentruber | EPHA1 | rs11771145 | 0.95 (0.93-0.96) | 3.30E-14 | LOF+REVEL>=25 (369), LOF+REVEL>=50 (180), LOF+REVEL>=75 (74), LOF (42) |

| | | | | | | | | |
|---|---|---|---|---|---|---|---|---|
| ENSG00000221855 | TAS2R41 | taste 2 receptor member 41 [Source:HGNC Symbol;Acc:HGNC:18883] | schwarzentruber | | | | | LOF+REVEL>=25 (20) |
| *ENSG00000185899* | *TAS2R60* | *taste 2 receptor member 60 [Source:HGNC Symbol;Acc:HGNC:20639]* | *schwarzentruber* | | | | | |
| ENSG00000159840 | ZYX | zyxin [Source:HGNC Symbol;Acc:HGNC:13200] | schwarzentruber | | | | | LOF+REVEL>=25 (99), LOF+REVEL>=50 (72), LOF+REVEL>=75 (19) |
| *ENSG00000066336* | *SPI1* | *Spi-1 proto-oncogene [Source:HGNC Symbol;Acc:HGNC:11241]* | *schwarzentruber* | SPI1 | rs10437655 | 1.06 (1.04-1.07) | 5.28E-14 | |
| ENSG00000197943 | PLCG2 | phospholipase C gamma 2 [Source:NCBI gene;Acc:5336] | schwarzentruber | PLCG2 | rs12446759;rs72824905 | 0.95 (0.94-0.96);0.74 (0.68-0.81) | 1.22e-13;8.48e-12 | LOF+REVEL>=25 (249), LOF+REVEL>=50 (69), LOF+REVEL>=75 (38), LOF (14) |
| ENSG00000071051 | NCK2 | NCK adaptor protein 2 [Source:HGNC Symbol;Acc:HGNC:7665] | schwarzentruber | NCK2 | rs143080277 | 1.47 (1.33-1.63) | 2.07E-13 | LOF+REVEL>=25 (24), LOF+REVEL>=50 (12) |
| ENSG00000149927 | DOC2A | double C2 domain alpha [Source:HGNC Symbol;Acc:HGNC:2985] | eadb | DOC2A | rs1140239 | 0.94 (0.93-0.96) | 2.59E-13 | LOF+REVEL>=25 (135), LOF+REVEL>=50 (56) |
| ENSG00000108091 | CCDC6 | coiled-coil domain containing 6 [Source:HGNC Symbol;Acc:HGNC:18782] | schwarzentruber | ANK3 | rs7068231 | 0.95 (0.94-0.96) | 3.32E-13 | LOF+REVEL>=25 (11) |
| ENSG00000161929 | SCIMP | SLP adaptor and CSK interacting membrane protein [Source:HGNC Symbol;Acc:HGNC:33504] | schwarzentruber | SCIMP | rs7225151 | 1.08 (1.05-1.1) | 4.13E-13 | |
| ENSG00000108379 | WNT3 | Wnt family member 3 [Source:HGNC Symbol;Acc:HGNC:12782] | nearest | WNT3 | rs199515 | 0.94 (0.93-0.96) | 9.34E-13 | LOF+REVEL>=25 (39), LOF+REVEL>=50 (28), LOF+REVEL>=75 (16) |
| ENSG00000142192 | APP | amyloid beta precursor protein [Source:HGNC Symbol;Acc:HGNC:620] | schwarzentruber | APP | rs2154481 | 0.95 (0.94-0.97) | 1.00E-12 | LOF+REVEL>=25 (189), LOF+REVEL>=50 (144), LOF+REVEL>=75 (19) |
| ENSG00000145214 | DGKQ | diacylglycerol kinase theta [Source:HGNC Symbol;Acc:HGNC:2856] | eadb | IDUA | rs3822030 | 0.95 (0.94-0.96) | 8.29E-12 | LOF+REVEL>=25 (105), LOF+REVEL>=50 (32), LOF+REVEL>=75 (15), LOF (12) |
| ENSG00000103510 | KAT8 | lysine acetyltransferase 8 [Source:HGNC Symbol;Acc:HGNC:17933] | schwarzentruber | BCKDK | rs889555 | 0.95 (0.94-0.97) | 1.96E-11 | LOF+REVEL>=25 (22), LOF+REVEL>=50 (18), LOF+REVEL>=75 (13) |
| ENSG00000178226 | PRSS36 | serine protease 36 [Source:HGNC Symbol;Acc:HGNC:26906] | schwarzentruber | | | | | LOF+REVEL>=25 (128), LOF+REVEL>=50 (58), LOF+REVEL>=75 (34), LOF (22) |

| | | | | | | | | |
|---|---|---|---|---|---|---|---|---|
| ENSG00000196549 | MME | membrane metalloendopeptidase [Source:HGNC Symbol;Acc:HGNC:7154] | eadb | MME | rs16824536;rs61762319 | 0.92 (0.89-0.95);1.16 (1.11-1.21) | 3.63e-08;2.16e-11 | LOF+REVEL>=25 (387), LOF+REVEL>=50 (317), LOF+REVEL>=75 (93), LOF (48) |
| *ENSG00000131042* | *LILRB2* | *leukocyte immunoglobulin like receptor B2 [Source:HGNC Symbol;Acc:HGNC:6606]* | *eadb* | LILRB2 | rs587709 | 1.05 (1.04-1.07) | 3.63E-11 | |
| *ENSG00000106460* | *TMEM106B* | *transmembrane protein 106B [Source:HGNC Symbol;Acc:HGNC:22407]* | *eadb* | TMEM106B | rs13237518 | 0.96 (0.94-0.97) | 4.88E-11 | |
| ENSG00000095585 | BLNK | B cell linker [Source:HGNC Symbol;Acc:HGNC:14211] | eadb | BLNK | rs6584063 | 0.89 (0.86-0.92) | 6.73E-11 | LOF+REVEL>=25 (22), LOF+REVEL>=50 (11) |
| ENSG00000167716 | WDR81 | WD repeat domain 81 [Source:HGNC Symbol;Acc:HGNC:26600] | eadb | WDR81 | rs35048651 | 1.06 (1.04-1.08) | 7.67E-11 | LOF+REVEL>=25 (573), LOF+REVEL>=50 (444), LOF+REVEL>=75 (37), LOF (17) |
| ENSG00000028528 | SNX1 | sorting nexin 1 [Source:HGNC Symbol;Acc:HGNC:11172] | nearest | SNX1 | rs3848143 | 1.05 (1.04-1.07) | 8.41E-11 | LOF+REVEL>=25 (175), LOF+REVEL>=50 (22), LOF (10), LOF+REVEL>=75 (10) |
| *ENSG00000219545* | *UMAD1* | *UBAP1-MVB12-associated (UMA) domain containing 1 [Source:HGNC Symbol;Acc:HGNC:48955]* | *nearest* | UMAD1 | rs6943429 | 1.05 (1.03-1.06) | 1.03E-10 | |
| ENSG00000146648 | EGFR | epidermal growth factor receptor [Source:HGNC Symbol;Acc:HGNC:3236] | eadb | SEC61G | rs76928645 | 0.93 (0.91-0.95) | 1.62E-10 | LOF+REVEL>=25 (302), LOF+REVEL>=50 (114), LOF+REVEL>=75 (21) |
| ENSG00000142192 | APP | amyloid beta precursor protein [Source:HGNC Symbol;Acc:HGNC:620] | schwarzentruber | ADAMTS1 | rs2830489 | 0.95 (0.94-0.97) | 1.69E-10 | LOF+REVEL>=25 (189), LOF+REVEL>=50 (144), LOF+REVEL>=75 (19) |
| ENSG00000184986 | TMEM121 | transmembrane protein 121 [Source:HGNC Symbol;Acc:HGNC:20511] | nearest | IGH gene cluster | rs7157106;rs10131280 | 1.05 (1.03-1.07);0.94 (0.92-0.96) | 1.99e-08;4.26e-10 | LOF+REVEL>=25 (12) |
| ENSG00000086289 | EPDR1 | ependymin related 1 [Source:HGNC Symbol;Acc:HGNC:17572] | nearest | EPDR1 | rs6966331 | 0.96 (0.94-0.97) | 4.64E-10 | LOF+REVEL>=25 (35), LOF+REVEL>=50 (17), LOF+REVEL>=75 (11) |
| *ENSG00000129911* | *KLF16* | *Kruppel like factor 16 [Source:HGNC Symbol;Acc:HGNC:16857]* | *nearest* | KLF16 | rs149080927 | 1.05 (1.04-1.07) | 5.09E-10 | |
| ENSG00000157368 | IL34 | interleukin 34 [Source:HGNC Symbol;Acc:HGNC:28529] | nearest | IL34 | rs4985556 | 1.07 (1.05-1.09) | 5.98E-10 | LOF+REVEL>=25 (167) |
| ENSG00000091536 | MYO15A | myosin XVA [Source:HGNC Symbol;Acc:HGNC:7594] | eadb | MYO15A | rs2242595 | 0.94 (0.92-0.96) | 1.11E-09 | LOF+REVEL>=25 (2797), LOF+REVEL>=50 (1683), LOF+REVEL>=75 (632), LOF (112) |

| | | | | | | | | |
|---|---|---|---|---|---|---|---|---|
| *ENSG00000168421* | *RHOH* | *ras homolog family member H [Source:HGNC Symbol;Acc:HGNC:686]* | *eadb* | RHOH | rs2245466 | 1.05 (1.03-1.06) | 1.22E-09 | |
| **ENSG00000165029** | **ABCA1** | **ATP binding cassette subfamily A member 1 [Source:HGNC Symbol;Acc:HGNC:29]** | **eadb** | ABCA1 | rs1800978 | 1.06 (1.04-1.08) | 1.59E-09 | LOF+REVEL>=25 (824), LOF+REVEL>=50 (614), LOF+REVEL>=75 (353), LOF (29) |
| *ENSG00000139405* | *RITA1* | *RBPJ interacting and tubulin associated 1 [Source:HGNC Symbol;Acc:HGNC:25925]* | *eadb* | TPCN1 | rs6489896 | 1.08 (1.05-1.1) | 1.80E-09 | |
| ENSG00000164733 | CTSB | cathepsin B [Source:HGNC Symbol;Acc:HGNC:2527] | eadb | CTSB | rs1065712 | 1.09 (1.06-1.12) | 1.94E-09 | LOF+REVEL>=25 (412), LOF+REVEL>=50 (336), LOF+REVEL>=75 (312), LOF (20) |
| *ENSG00000154124* | *OTULIN* | *OTU deubiquitinase with linear linkage specificity [Source:HGNC Symbol;Acc:HGNC:25118]* | *eadb* | ANKH | rs112403360 | 1.09 (1.06-1.12) | 2.27E-09 | |
| ENSG00000249853 | HS3ST5 | heparan sulfate-glucosamine 3-sulfotransferase 5 [Source:HGNC Symbol;Acc:HGNC:19419] | nearest | HS3ST5 | rs785129 | 1.04 (1.03-1.06) | 2.40E-09 | LOF+REVEL>=25 (214), LOF+REVEL>=50 (165) |
| ENSG00000203896 | LIME1 | Lck interacting transmembrane adaptor 1 [Source:HGNC Symbol;Acc:HGNC:26016] | eadb | SLC2A4RG | rs6742 | 0.95 (0.93-0.97) | 2.58E-09 | LOF+REVEL>=25 (14), LOF (14), LOF+REVEL>=50 (14), LOF+REVEL>=75 (14) |
| ENSG00000107679 | PLEKHA1 | pleckstrin homology domain containing A1 [Source:HGNC Symbol;Acc:HGNC:14335] | eadb | PLEKHA1 | rs7908662 | 0.96 (0.95-0.97) | 2.59E-09 | LOF+REVEL>=25 (21) |
| ENSG00000115825 | PRKD3 | protein kinase D3 [Source:HGNC Symbol;Acc:HGNC:9408] | nearest | PRKD3 | rs17020490 | 1.06 (1.04-1.08) | 3.29E-09 | LOF+REVEL>=25 (95), LOF+REVEL>=50 (63), LOF+REVEL>=75 (16) |
| ENSG00000103811 | CTSH | cathepsin H [Source:HGNC Symbol;Acc:HGNC:2535] | eadb | CTSH | rs12592898 | 0.94 (0.92-0.96) | 4.18E-09 | LOF+REVEL>=25 (63), LOF+REVEL>=50 (39), LOF+REVEL>=75 (31), LOF (11) |
| ENSG00000161640 | SIGLEC11 | sialic acid binding Ig like lectin 11 [Source:HGNC Symbol;Acc:HGNC:15622] | eadb | SIGLEC11 | rs9304690 | 1.05 (1.03-1.07) | 4.74E-09 | LOF+REVEL>=25 (25), LOF+REVEL>=75 (10), LOF+REVEL>=50 (10), LOF (10) |
| ENSG00000003147 | ICA1 | islet cell autoantigen 1 [Source:HGNC Symbol;Acc:HGNC:5343] | eadb | ICA1 | rs10952097 | 1.07 (1.05-1.1) | 6.81E-09 | LOF+REVEL>=25 (110), LOF+REVEL>=50 (33), LOF+REVEL>=75 (19) |
| ENSG00000134243 | SORT1 | sortilin 1 [Source:HGNC Symbol;Acc:HGNC:11186] | eadb | SORT1 | rs141749679 | 1.38 (1.24-1.54) | 7.54E-09 | LOF+REVEL>=25 (42), LOF+REVEL>=50 (17) |

| | | | | | | | | |
|---|---|---|---|---|---|---|---|---|
| ENSG00000145901 | TNIP1 | TNFAIP3 interacting protein 1 [Source:HGNC Symbol;Acc:HGNC:16903] | eadb | TNIP1 | rs871269 | 0.96 (0.95-0.97) | 8.67E-09 | LOF+REVEL>=25 (33) |
| *ENSG00000153814* | *JAZF1* | *JAZF zinc finger 1 [Source:HGNC Symbol;Acc:HGNC:28917]* | *eadb* | JAZF1 | rs1160871 | 0.95 (0.93-0.97) | 9.83E-09 | |
| ENSG00000163596 | ICA1L | islet cell autoantigen 1 like [Source:HGNC Symbol;Acc:HGNC:14442] | eadb | WDR12 | rs139643391 | 0.94 (0.92-0.96) | 1.08E-08 | LOF+REVEL>=25 (61), LOF (52), LOF+REVEL>=50 (29), LOF+REVEL>=75 (17) |
| ENSG00000103241 | FOXF1 | forkhead box F1 [Source:HGNC Symbol;Acc:HGNC:3809] | nearest | FOXF1 | rs16941239 | 1.13 (1.08-1.17) | 1.29E-08 | LOF+REVEL>=25 (26) |
| ENSG00000138600 | SPPL2A | signal peptide peptidase like 2A [Source:HGNC Symbol;Acc:HGNC:30227] | schwarzentruber | SPPL2A | rs8025980 | 0.96 (0.94-0.97) | 1.32E-08 | LOF+REVEL>=25 (226), LOF+REVEL>=50 (213) |
| ENSG00000125826 | RBCK1 | RANBP2-type and C3HC4-type zinc finger containing 1 [Source:HGNC Symbol;Acc:HGNC:15864] | eadb | RBCK1 | rs1358782 | 0.95 (0.94-0.97) | 1.55E-08 | LOF+REVEL>=25 (52), LOF+REVEL>=50 (18) |
| ENSG00000151694 | ADAM17 | ADAM metallopeptidase domain 17 [Source:HGNC Symbol;Acc:HGNC:195] | eadb | ADAM17 | rs72777026 | 1.06 (1.04-1.08) | 2.72E-08 | LOF+REVEL>=25 (75), LOF+REVEL>=50 (15) |
| ENSG00000178573 | MAF | MAF bZIP transcription factor [Source:HGNC Symbol;Acc:HGNC:6776] | eadb | MAF | rs450674 | 0.96 (0.95-0.98) | 3.16E-08 | LOF+REVEL>=25 (21) |

#### 2.2.6 Table S6: Burden testing of prioritized genes in GWAS loci.

| GWAS-targeted analysis | | | Burden test (variant MAF <1%) | | | | | | Burden test (variant MAF < 0.1%) | | | |
|---|---|---|---|---|---|---|---|---|---|---|---|---|
| **locus** | **sentinel GWAS SNPs** | **locus prio source** | **gene** | **group** | **pvalue** | **FDR** | **#variant / #carriers** | **case / control OR (95% CI)** | **pvalue** | **#variant / #carriers** | **fraction very rare** | **case / control OR (95% CI)** |
| SORL1 | rs74685827 rs11218343 | schwarzentruber | SORL1 | LOF+REVEL≥25 | 2.5E-09 | <<0.01% | 298 / 1265 | 1.4 (1.2-1.5) | 1.4E-18 | 293 / 582 | 47% | 1.9 (1.7-2.3) |
| | | | | LOF+REVEL≥50 | 2.5E-25 | <<0.01% | 201 / 356 | 2.7 (2.2-3.3) | 2.3E-25 | 200 / 354 | 99% | 2.6 (2.2-3.2) |
| | | | | LOF+REVEL≥75 | 5.3E-22 | <<0.01% | 117 / 204 | 4.0 (3.0-5.4) | 4.3E-22 | 117 / 204 | 100% | 4.0 (3.0-5.4) |
| | | | | LOF | 7.9E-21 | <<0.01% | 49 / 63 | 20.0 (11.8-34.0) | 8.2E-21 | 49 / 63 | 100% | 20.0 (11.8-34.0) |
| TREM2 | rs10947943 rs60755019 rs143332484 rs75932628 | schwarzentruber | TREM2 | LOF+REVEL≥25 | 7.9E-20 | <<0.01% | 20 / 379 | 3.0 (2.4-3.7) | 3.6E-04 | 19 / 77 | 20% | 2.2 (1.4-3.6) |
| ABCA7 | rs12151021 | schwarzentruber | ABCA7 | LOF+REVEL≥25 | 3.4E-10 | <<0.01% | 338 / 1319 | 1.4 (1.3-1.6) | 1.1E-07 | 316 / 762 | 57% | 1.5 (1.3-1.8) |
| | | | | LOF+REVEL≥50 | 8.2E-07 | <<0.01% | 216 / 1016 | 1.4 (1.2-1.5) | 2.9E-04 | 202 / 493 | 48% | 1.5 (1.2-1.8) |
| | | | | LOF+REVEL≥75 | 1.7E-05 | 0.03% | 104 / 395 | 1.5 (1.2-1.8) | 1.6E-02 | 98 / 234 | 59% | 1.4 (1.0-1.8) |
| | | | | LOF | 1.9E-03 | 1.5% | 50 / 112 | 1.6 (1.1-2.4) | 9.2E-03 | 48 / 92 | 82% | 1.6 (1.0-2.4) |
| SLC24A4/ RIN3 | rs7401792 rs12590654 | schwarzentruber | RIN3 | LOF+REVEL≥25 | 1.6E-05 | 0.03% | 44 / 622 | 1.4 (1.2-1.6) | 3.4E-02 | 42 / 129 | 21% | 1.4 (1.0-2.1) |
| | | | | LOF+REVEL≥50 | 1.0E-05 | 0.02% | 23 / 583 | 1.4 (1.2-1.7) | 1.5E-02 | 21 / 89 | 15% | 1.8 (1.2-2.8) |
| MINDY2 | rs602602 | schwarzentruber | ADAM10 | LOF+REVEL≥25 | 4.1E-04 | 0.48% | 34 / 44 | 2.0 (1.1-3.7) | 4.5E-04 | 34 / 44 | 100% | 2.0 (1.1-3.7) |
| | | | | LOF+REVEL≥50 | 2.3E-05 | 0.04% | 20 / 22 | 3.3 (1.4-7.6) | 2.3E-05 | 20 / 22 | 100% | 3.3 (1.4-7.6) |
| | | | | LOF+REVEL≥75 | 3.6E-05 | 0.06% | 13 / 14 | 3.6 (1.3-9.9) | 3.6E-05 | 13 / 14 | 100% | 3.6 (1.3-9.9) |
| | | | | LOF | 3.8E-04 | 0.47% | 9 / 9 | 3.6 (0.9-21.8) | 3.8E-04 | 9 / 9 | 100% | 3.6 (0.9-23.5) |
| ABCA1 | rs1800978 | eadb | ABCA1 | LOF+REVEL≥25 | 1.6E-03 | 1.3% | 274 / 796 | 1.3 (1.1-1.5) | 6.2E-07 | 268 / 509 | 62% | 1.6 (1.4-1.9) |
| | | | | LOF+REVEL≥50 | 6.8E-03 | 4.5% | 187 / 607 | 1.3 (1.1-1.5) | 6.8E-07 | 184 / 337 | 55% | 1.9 (1.5-2.4) |
| | | | | LOF+REVEL≥75 | 5.6E-05 | 0.08% | 115 / 352 | 1.5 (1.2-1.9) | 1.4E-08 | 113 / 202 | 58% | 2.4 (1.8-3.3) |
| | | | | LOF | 3.7E-04 | 0.47% | 24 / 29 | 5.0 (2.3-10.9) | 3.7E-04 | 24 / 29 | 100% | 5.0 (2.3-10.9) |
| PTK2B/ CLU | rs73223431 rs11787077 | schwarzentruber | CLU | LOF+REVEL≥25 | 5.0E-04 | 0.52% | 24 / 26 | 3.6 (1.6-8.3) | 5.0E-04 | 24 / 26 | 100% | 3.6 (1.6-8.3) |
| | | | | LOF+REVEL≥50 | 1.1E-03 | 0.97% | 14 / 15 | 5.4 (1.6-28.6) | 1.1E-03 | 14 / 15 | 100% | 5.3 (1.6-30.1) |
| | | | | LOF+REVEL≥75 | 5.0E-04 | 0.52% | 12 / 12 | 9.9 (1.6-44.0) | 5.0E-04 | 12 / 12 | 100% | 9.8 (1.6-43.0) |
| | | | | LOF | 2.6E-03 | 2.0% | 10 / 10 | 7.3 (1.3-43.5) | 2.6E-03 | 10 / 10 | 100% | 7.3 (1.3-44.1) |
| SPDYE3 | rs7384878 | schwarzentruber | ZCWPW1 | LOF+REVEL≥25 | 6.1E-03 | 4.2% | 22 / 77 | 1.8 (1.2-2.9) | 5.0E-03 | 21 / 76 | 99% | 1.8 (1.2-2.9) |
| | | | | LOF+REVEL≥50 | 3.1E-03 | 2.2% | 16 / 70 | 1.9 (1.2-3.1) | 3.1E-03 | 16 / 70 | 100% | 1.9 (1.2-3.1) |
| | | | | LOF+REVEL≥75 | 1.1E-03 | 0.97% | 11 / 15 | 5.0 (1.7-30.3) | 7.7E-04 | 11 / 15 | 100% | 5.0 (1.7-29.8) |
| | | | | LOF | 7.8E-04 | 0.76% | 11 / 15 | 5.0 (1.7-28.7) | 7.9E-04 | 11 / 15 | 100% | 5.0 (1.7-30.5) |
| ACE | rs4277405 | schwarzentruber | ACE | LOF+REVEL≥75 | 9.0E-04 | 0.84% | 38 / 99 | 2.0 (1.3-2.9) | 9.3E-04 | 38 / 99 | 100% | 2.0 (1.3-2.9) |

Performed on the mega-analysis dataset (excluding exome-extracts). Complete list of performed tests in **Table S5**. Grey; no difference with the gnomAD MAF<1%.

#### 2.2.7 Table S7 Age burden trends in cases and controls separately

| | | *for reference* | **Case age-at-onset trends (i.e. ordinal logistic burden test without controls, by age-at-onset)** | | | | | **Control age trend (i.e. ordinal logistic burden test without cases, by age-last-seen)** | | |
|---|---|---|---|---|---|---|---|---|---|---|
| **gene** | **group** | **Case/control OR (95% CI)** | **AD-age (ord-OR, 95%CI)** | **pvalue AD-age 65-,65-75,75-85,85+** | **LOAD-age (ord-OR, 95%CI)** | **pvalue LOAD-age 65-75,75-85,85+** | **Carrier frequency by age-at-onset 65- / 65-75 / 75-85 / 85+ [controls]** | **pvalue 65-,65-75,75-85,85+** | **(ord-OR, 95% CI)** | **Carrier frequency by age-last-seen 65- / 65-75 / 75-85 / 85+** |
| SORL1 | LOF+REVEL≥50 | 2.1 (1.7-2.5) | 1.8 (1.4-2.2) | **1.9E-08*** | 1.6 (1.2-2.1) | **2.5E-03*** | 2.75% / 1.98% / 1.23% / 1.18% [0.68%] | 9.9E-01 | 1.0 (0.7-1.4) | 0.62% / 0.80% / 0.59% / 0.66% |
| | LOF | 19.8 (11.9-32.7) | 3.9 (2.3-6.4) | **2.8E-08*** | 2.8 (1.2-6.7) | **1.5E-02** | 0.78% / 0.33% / 0.14% / 0.11% [0.02%] | 1.6E-01 | 5.4 (0.5-60.9) | 0.03% / 0.06% / 0.00% / 0.00% |
| TREM2 | LOF+REVEL≥25 | 2.8 (2.3-3.5) | 1.5 (1.2-1.8) | **8.2E-05*** | 1.9 (1.5-2.5) | **1.6E-06*** | 2.27% / 2.70% / 1.62% / 1.11% [0.75%] | **3.3E-04*** | 1.9 (1.3-2.7) | 0.92% / 1.14% / 0.62% / 0.51% |
| | LOF | 2.1 (1.2-3.4) | 0.9 (0.5-1.5) | 6.4E-01 | 1.1 (0.6-2.3) | 7.3E-01 | 0.21% / 0.36% / 0.14% / 0.37% [0.16%] | 3.3E-01 | 0.6 (0.2-1.6) | 0.14% / 0.00% / 0.10% / 0.15% |
| ABCA7 | LOF+REVEL≥25 | 1.4 (1.3-1.6) | 1.2 (1.1-1.4) | **7.4E-04*** | 1.1 (1.0-1.3) | *7.9E-02* | 6.18% / 5.33% / 4.95% / 4.73% [3.90%] | 3.4E-01 | 0.9 (0.8-1.1) | 3.79% / 3.88% / 3.77% / 4.05% |
| | LOF | 1.7 (1.1-2.4) | 1.9 (1.3-2.8) | **1.4E-03*** | 2.4 (1.3-4.3) | **2.4E-03** | 0.62% / 0.59% / 0.39% / 0.11% [0.27%] | 8.4E-01 | 0.9 (0.6-1.6) | 0.16% / 0.46% / 0.34% / 0.31% |
| ATP8B4 | LOF+REVEL≥25 | 1.4 (1.2-1.6) | 1.4 (1.0-1.8) | **2.8E-02** | 1.4 (0.9-2.0) | 1.0E-01 | 3.56% / 3.22% / 3.47% / 2.36% [2.09%] | 6.4E-01 | 1.1 (0.9-1.3) | 1.79% / 2.23% / 2.29% / 1.92% |
| | LOF | 1.1 (0.6-1.9) | 1.0 (0.5-2.1) | 9.3E-01 | 0.7 (0.3-1.6) | 3.8E-01 | 0.21% / 0.10% / 0.20% / 0.18% [0.16%] | 9.8E-01 | 1.0 (0.4-2.3) | 0.14% / 0.11% / 0.15% / 0.11% |
| ABCA1 | LOF+REVEL≥75 | 1.6 (1.3-2.0) | 1.4 (1.2-1.8) | **9.9E-04*** | 1.7 (1.2-2.3) | **6.9E-04*** | 1.91% / 2.01% / 1.26% / 1.07% [1.13%] | 2.2E-01 | 1.2 (0.9-1.6) | 1.30% / 1.14% / 1.16% / 0.97% |
| | LOF | 3.5 (1.9-6.4) | 1.7 (0.9-3.2) | *8.0E-02* | 1.5 (0.7-3.6) | 3.3E-01 | 0.28% / 0.21% / 0.20% / 0.11% [0.08%] | 5.0E-01 | 1.4 (0.5-4.3) | 0.14% / 0.11% / 0.07% / 0.04% |
| ADAM10 | LOF+REVEL≥50 | 4.7 (2.0-10.8) | 4.0 (1.5-11.0) | **2.6E-03*** | 1.6 (0.4-7.2) | 5.3E-01 | 0.23% / 0.05% / 0.06% / 0.04% [0.02%] | 6.6E-01 | 0.7 (0.1-4.3) | 0.03% / 0.06% / 0.00% / 0.04% |
| RIN3 | LOF+REVEL≥50 | 1.4 (1.2-1.7) | 1.2 (1.0-1.4) | 1.2E-01 | 0.9 (0.7-1.2) | 5.1E-01 | 2.67% / 1.84% / 2.36% / 2.11% [1.62%] | 9.6E-01 | 1.0 (0.8-1.3) | 1.47% / 2.15% / 1.69% / 1.66% |
| | LOF | 2.1 (0.5-9.3) | 1.2 (0.2-5.9) | 8.3E-01 | 0.3 (0.0-3.2) | 2.8E-01 | 0.06% / 0.03% / 0.00% / 0.08% [0.01%] | 2.2E-01 | 4.5 (0.4-54.5) | 0.03% / 0.00% / 0.03% / 0.00% |
| CLU | LOF+REVEL≥25 | 3.6 (1.6-8.3) | 2.2 (1.0-5.1) | *5.5E-02* | 1.2 (0.4-4.1) | 7.4E-01 | 0.23% / 0.11% / 0.09% / 0.08% [0.03%] | 2.7E-01 | 0.4 (0.1-2.3) | 0.00% / 0.00% / 0.05% / 0.04% |
| | LOF | 7.3 (1.9-27.2) | 3.9 (1.0-15.0) | **3.2E-02** | 3.0 (0.3-31.2) | 3.3E-01 | 0.12% / 0.05% / 0.03% / 0.00% [0.01%] | 5.0E-01 | | -- / -- / -- / -- |
| ZCWPW1 | LOF | 5.0 (1.9-13.5) | 2.6 (0.9-7.7) | *6.0E-02* | 0.7 (0.1-3.2) | 6.2E-01 | 0.15% / 0.03% / 0.09% / 0.04% [0.01%] | 3.3E-01 | 3.7 (0.3-54.5) | 0.03% / 0.00% / 0.03% / 0.00% |
| ACE | LOF+REVEL≥75 | 2.0 (1.3-2.9) | 1.4 (0.9-2.2) | 1.0E-01 | 1.1 (0.6-2.0) | 6.7E-01 | 0.60% / 0.49% / 0.32% / 0.34% [0.20%] | **2.0E-03*** | 3.0 (1.5-6.0) | 0.39% / 0.18% / 0.23% / 0.07% |
| | LOF | 1.4 (0.8-2.4) | 1.4 (0.7-2.6) | 3.5E-01 | 0.8 (0.4-1.9) | 6.5E-01 | 0.27% / 0.16% / 0.17% / 0.15% [0.14%] | **1.6E-03*** | 3.9 (1.6-9.3) | 0.33% / 0.09% / 0.16% / 0.02% |

ord-OR: OR based on ordinal logistic regression. Effect sizes (odds ratios, ORs) indicate the increased enrichment of carriers in the direction of the younger categories. *: significant after holm-bonferoni multiple testing correction. Note that the trend test 65-,65-75,75-85,85+ incorporates a difference between EOAD and LOAD samples. This difference was also used in our primary test to select these genes. In column F and G we therefore include an analysis which only considers the trend 65-75,75-85,85+. *Spearman rank correlation between case/control OR and AD-age ordinal-OR: cor=0.78, p=0.0001*

#### 2.2.8 Table S8 Contribution of extremely rare variants to the burden test

| | | Burden MAF < 0.01 | | | Burden MAF < 0.0001 | | | | |
|---|---|---|---|---|---|---|---|---|---|
| **gene** | **group** | **variants / carriers** | **carriers per variant** | **p-value** | **variants / carriers** | **carriers per variant** | **p-value** | **z-score ratio p-values** | **Case/control OR (MAF < 0.01)** |
| SORL1 | LOF+REVEL≥50 | 212 / 418 | 1.97 | 2.0E-25 | 187 / 274 | 0.46 | 6.2E-31 | **1.11** | 2.1 |
| | LOF | 51 / 68 | 1.33 | 8.8E-22 | 51 / 68 | 1.34 | 8.9E-22 | **1.00** | 19.8 |
| TREM2 | LOF+REVEL≥25 | 26 / 441 | 16.96 | 1.4E-21 | 18 / 26 | 1.47 | 1.7E-03 | 0.31 | 2.8 |
| | LOF | 12 / 66 | 5.50 | 2.4E-02 | 10 / 15 | 1.54 | 8.7E-02 | 0.69 | 2.1 |
| ABCA7 | LOF+REVEL≥25 | 351 / 1489 | 4.24 | 6.0E-13 | 257 / 428 | 1.66 | 6.1E-05 | 0.54 | 1.4 |
| | LOF | 49 / 119 | 2.43 | 8.8E-04 | 43 / 77 | 1.79 | 1.7E-02 | 0.67 | 1.7 |
| ATP8B4 | LOF+REVEL≥25 | 94 / 850 | 9.04 | 7.4E-07 | 73 / 99 | 1.36 | 6.8E-01 | -0.10 | 1.4 |
| ABCA1 | LOF+REVEL≥75 | 122 / 442 | 3.62 | 5.1E-07 | 108 / 165 | 1.53 | 1.2E-09 | **1.22** | 1.6 |
| | LOF | 27 / 47 | 1.74 | 5.5E-05 | 25 / 32 | 1.28 | 5.6E-04 | **0.84** | 3.5 |
| ADAM10 | LOF+REVEL≥50 | 19 / 22 | 1.16 | 5.1E-06 | 19 / 22 | 1.16 | 5.2E-06 | **1.00** | 4.7 |
| RIN3 | LOF+REVEL≥50 | 23 / 583 | 25.35 | 1.0E-05 | 17 / 23 | 1.36 | 5.9E-01 | -0.06 | 1.4 |
| CLU | LOF+REVEL≥25 | 24 / 26 | 1.08 | 5.0E-04 | 23 / 25 | 1.09 | 9.2E-04 | **0.95** | 3.6 |
| | LOF | 10 / 10 | 1.00 | 2.7E-03 | 10 / 10 | 1.00 | 2.7E-03 | **1.00** | 7.3 |
| ZCWPW1 | LOF | 11 / 15 | 1.36 | 7.8E-04 | 11 / 15 | 1.44 | 7.8E-04 | **1.00** | 5 |
| ACE | LOF+REVEL≥75 | 38 / 99 | 2.61 | 9.0E-04 | 33 / 55 | 1.68 | 1.4E-01 | 0.35 | 2 |

Burden tests were performed on the mega dataset for the categories shown in **Table 3**, both for a MAF > 0.01 and a MAF > 0.0001 threshold. Burden tests with p>0.05 for MAF < 0.01 were excluded. Numbers of variants and carriers are shown for both burden tests. P-values for both tests were compared by calculating a z-score ratio. Z-scores above 0.75 are shown in bold. SORL1 and ABCA1 increased in significance due to the strict MAF threshold for the variant category that includes missense variants, scoring a z-score ratio above 1. To determine if associations with high odds ratios can be linked to a higher contribution of extremely rare variants to the burden significance, the spearman rank correlation was calculated between z-score ratios and the case/control odds ratio of the Burden MAF < 0.01 test. This gave a positive spearman rank correlation of 0.56, p=0.03.

#### 2.2.9 Table S9 Variant features

| Gene | Test | P<br>effect size ~ rareness<br>(ordinal logistic) | P<br>LOF ≥ missense<br>(ordinal OR) | P<br>LOF ≥ missense<br>(case/control OR) |
|---|---|---|---|---|
| SORL1 | LOF+REVEL≥50 | **<5.0e-06*** | **<5.0e-09*** | **<5.0e-09*** |
| TREM2 | LOF+REVEL≥25 | 5.5E-01 | 9.5E-01 | 9.2E-01 |
| TREM2 | LOF+REVEL≥25 [refined] | 5.4E-01 | 4.1E-01 | *6.8E-02* |
| ABCA7 | LOF+REVEL≥25 | 1.0E+00 | *8.3E-02* | 2.0E-01 |
| ATP8B4 | LOF+REVEL≥25 | 1.0E+00 | 8.1E-01 | 8.3E-01 |
| ABCA1 | LOF+REVEL≥75 | **1.3e-04*** | **5.6e-03*** | **5.3e-03*** |
| ABCA1 | LOF+REVEL≥75 [refined] | 3.2E-01 | *7.8E-02* | 1.2E-01 |
| ADAM10 | LOF+REVEL≥50 | - | 1.7E-01 | 6.5E-01 |
| RIN3 | LOF+REVEL≥50 | 1.0E+00 | 2.7E-01 | 2.8E-01 |
| CLU | LOF+REVEL≥25 | - | 1.3E-01 | 1.1E-01 |
| ZCWPW1 | LOF | **4.5E-02** | NA | NA |
| ACE | LOF+REVEL≥75 | 1.0E+00 | 9.2E-01 | 9.7E-01 |

*:significant after holm-bonferoni multiple testing correction.

- effect size ~ variant rareness: association between variant effect size and variant rareness (allele count 1, 2, 3-5, 6-10, 10+), See section 1.10.4.
- LOF >= missense: indicates if the burden of LOF variants has a larger effect size as the burden of missense variants. Note that in **Figure 2C** we report only the results for the refined burden, such that only SORL1 has a significant association after multiple testing correction.

#### 2.2.10 Table S10 Carriers of multiple variants in identified genes

| Carrier type | All | Controls | Cases | EOAD | LOAD |
|---|---|---|---|---|---|
| ≥1 affected gene | 11.9% | 8.8% | 15.0% | 17.7% | 13.5% |
| ≥1 affected gene (incl. APOE with ε4/ε4) | 16.9% | 9.7% | 22.0% | 30.2% | 17.6% |
| ≥1 affected gene (incl. APOE with ε4) | 45.0% | 27.5% | 57.3% | 64.6% | 53.4% |
| ≥2 affected genes | 0.59% | 0.31% | 0.87% | 1.17% | 0.71% |
| - expected | 0.59% (0.53%-0.66%) | 0.30% (0.22%-0.37%) | 0.95% (0.81%-1.09%) | 1.35% (1.08%-1.58%) | 0.77% (0.62%-0.93%) |
| ≥2 affected genes (incl. APOE with ε4/ε4) | 1.34% | 0.42% | 1.99% | 3.42% | 1.24% |
| - expected | 1.25% (1.12%-1.38%) | 0.36% (0.27%-0.53%) | 2.07% (1.90%-2.23%) | 3.61% (3.26%-3.96%) | 1.34% (1.14%-1.53%) |
| ≥2 affected genes (incl. APOE with ε4) | **5.31%** | 1.99% | 7.66% | 10.06% | 6.39% |
| - expected | 4.90% (4.71%-5.10%) | 2.03% (1.84%-2.26%) | 7.55% (7.28%-7.82%) | 9.92% (9.43%-10.42%) | 6.40% (6.06%-6.75%) |

Genes considered: *SORL1, TREM2, ABCA7, ATP8B4, ABCA1, ADAM10, RIN3, CLU, ZCWPW1, ACE*. Affected gene: Carries at least one variant in the gene with an impact above the most significant variant threshold for that gene (**Table 3).** Expected: under a model in which affected gene alleles in the dataset are randomly distributed across all/Control/Case/EOAD/LOA samples respectively. Values and confidence intervals are generated by sampling 1000 times. "≥2 affected genes (incl. APOE with ε4/ε4)" means that a person has a total of at least 2 affected genes, e.g. *SORL1 and TREM2, or ABCA7 and APOE* (only ε4/ε4 considered damaging)

##### 2.2.11 Table S11: Testing for interaction with APOE - E4 genotype

| Gene | Test | APOE-interaction pvalue |
|---|---|---|
| SORL1 | LOF+REVEL≥50 | 6.0E-01 |
| TREM2 | LOF+REVEL≥25 | 5.9E-01 |
| ABCA7 | LOF+REVEL≥25 | 4.8E-01 |
| ATP8B4 | LOF+REVEL≥25 | 7.5E-01 |
| ABCA1 | LOF+REVEL≥75 | 4.4E-01 |
| ADAM10 | LOF+REVEL≥50 | 4.0E-01 |
| RIN3 | LOF+REVEL≥50 | 8.5E-01 |
| CLU | LOF+REVEL≥25 | 2.3E-01 |
| ZCWPW1 | LOF | 6.4E-02 |
| ACE | LOF+REVEL≥75 | 3.3E-01 |

APOE - E4 dosage was used to test for an interaction effect. The following model was used: burden_score + apoe_e4_dosage + burden_score * apoe_e4_dosage. A p-value was calculated based on a likelihood ratio test between a model with the interaction effect, and one without. Studies in which the APOE genotype was used as part of the selection (ADSP, Barcelona, StEP-AD) were excluded.

#### 2.2.12 Table S12 Somatic Mutation Check

| Gene | Test | Average allele balance |
|---|---|---|
| SORL1 | LOF+REVEL≥50 | 0.514 |
| TREM2 | LOF+REVEL≥25 | 0.505 |
| ABCA7 | LOF+REVEL≥25 | 0.520 |
| ATP8B4 | LOF+REVEL≥25 | 0.530 |
| ABCA1 | LOF+REVEL≥75 | 0.522 |
| ADAM10 | LOF+REVEL≥50 | 0.529 |
| RIN3 | LOF+REVEL≥50 | 0.529 |
| CLU | LOF+REVEL≥25 | 0.518 |
| ZCWPW1 | LOF | 0.521 |
| ACE | LOF+REVEL≥75 | 0.514 |
| Reference | | |
| All genes | (cMAC >= 10) | 0.524<br>(IQR: 0.512-0.538)<br>(95% CI: 0.482-0.581) |
| TET2 | LOF | 0.669 |
| DNTM3A | LOF | 0.664 |

Average allele balance of damaging heterozygous genotypes in the burden analysis (mega-analysis dataset). An allele balance of 0.5 indicates similar number of reads covering the reference and alternate allele, while an allele balance of 1.0 indicates that only the reference allele is covered. A value slightly above 0.5 is normal in exomes due to a slight reference read bias. TET2 and DNTM3A LOF variants are known to be involved in age-related clonal hematopoiesis (ARCH)[58].

#### 2.2.13 Table S13: Validation of variant selection

| Gene + transcripts (canonical=bold) | | Stage 1 | OR (95% CI) | Protein change (per transcript) (bold: name in text) | Impact prediction | | | MAF | Gnomad non-neuro (max. freq. pop) | | Stage 2 (dir. S1) | OR (95% CI) | |
|---|---|---|---|---|---|---|---|---|---|---|---|---|---|
| **SNP** | **Type** | **FDR** | **Stage 1** | | **REVEL** | **CADD** | **Clinvar** | **Mega** | **Freq.** | **Pop.** | **h-bonf** | **Stage 2** | **Mega** |
| **SORL1** | **ENST00000260197: A**, ENST00000525532: B, ENST00000534286: C, ENST00000532694: D, ENST00000527934: E | | | | | | | | | | | | |
| rs140384365 | addition | 3.8% | 2.49 (1.22-5.07) | **A: V1459I,** B: V403I, C: V369I, D: V305I, E: V74I | 0.09 | 12.3 | Lik. benign | 0.10% | 0.05% | nfe | 6.3E-01 | 0.86 (0.37-2.01) | 1.33 (0.79-2.23) |
| rs143536682 | outlier | 3.8% | 0.53 (0.19-1.47) | **A: S2175R,** B: S1119R, C: S1085R, D: S1021R, E: S790R | 0.81 | 25.0 | | 0.03% | 0.03% | nfe | <=5 car. | <=5 car. | 0.73 (0.29-1.85) |
| **TREM2** | **ENST00000373113: A**, ENST00000373122: B, ENST00000338469: C | | | | | | | | | | | | |
| rs142232675 | addition | 0.05% | 2.63 (1.56-4.45) | A,B,C: **D87N** | 0.20 | 19.8 | Conflict. int. of path. | 0.14% | 0.18% | nfe | 7.4E-01 | 0.74 (0.30-1.83) | 1.71 (1.09-2.66) |
| rs538447052 | outlier | 4.1% | 1.91 (0.71-5.08) | B: splice acceptor variant | LOF | 5.1 | | 0.06% | 0.03% | nfe | **1.0E-03** | 0.53 (0.18-1.57) | 1.23 (0.62-2.43) |
| **ABCA7** | **ENST00000263094: A**, ENST00000433129: B, ENST00000435683: C | | | | | | | | | | | | |
| rs546173555 | outlier | 1.1% | 1.09 (0.37-3.20) | A,B: **R19W** | 0.54 | 23.9 | | 0.02% | 0.01% | nfe | <=5 car. | <=5 car. | 1.49 (0.51-4.36) |
| rs117187003 | outlier | 0.1% | 0.84 (0.61-1.15) | A,B: **V1599M,** C: V1461M | 0.58 | 25.5 | Lik. benign | 0.40% | 0.43% | nfe | 2.9E-01 | 1.62 (0.99-2.67) | 1.00 (0.77-1.30) |
| rs143614132 | outlier | 20% | 0.67 (0.28-1.60) | A,B: **G1820S,** C: G1682S | 0.91 | 32.0 | | 0.07% | 0.06% | nfe | 2.9E-01 | 1.01 (0.35-2.87) | 0.71 (0.37-1.35) |
| **ATP8B4** | **ENST00000284509: A**, ENST00000559829: B | | | | | | | | | | | | |
| rs201949459 | outlier | 19% | 0.81 (0.28-2.38) | A,B: **P83A** | 0.84 | 25.2 | | 0.03% | 0.09% | sas | 6.3E-01 | 0.85 (0.10-6.98) | 0.84 (0.33-2.19) |
| rs74811880 | addition | 0.3% | 3.14 (1.55-6.34) | A,B: **H987R** | 0.26 | 15.0 | | 0.08% | 0.08% | nfe | 4.8E-01 | 1.58 (0.45-5.53) | 2.30 (1.26-4.19) |
| **ABCA1** | **ENST00000374736: A**, ENST00000423487: B, ENST00003074733: C | | | | | | | | | | | | |
| rs145183203 | outlier | 1.9% | 0.92 (0.56-1.51) | A,B: P85L, C: P25L | 0.84 | 24.7 | Lik. benign | 0.20% | 0.14% | nfe | **4.1E-02** | 0.85 (0.46-1.56) | 0.84 (0.58-1.21) |
| rs140365800 | outlier | 13% | 0.81 (0.29-2.22) | A: D1018G | 0.84 | 32.0 | Conflict. int. of path. | 0.05% | 0.11% | amr | **4.8E-02** | 0.42 (0.13-1.34) | 0.85 (0.41-1.77) |

The variant-selection approach was validated for variants that i) were a missense or LOF variant without QC issues, ii) had at least 15 carriers and iii) a MAF < 1% both in our dataset and in the gnomAD non-neuro populations. Variants that were in the burden test were considered for removal when their effect size significantly deviated from other LOF variants or missense in their pathogenicity category (fisher exact test). Variants that were not in the burden test were considered for addition if they significantly associated with AD in the same direction as the most significant burden test (logistic regression). All considered variants are shown in the gene-specific variant **Tables SG1-SG10** (i.e. across all genes in refinement columns AP-AU). Multiple testing correction was performed per gene, with FDR used for Stage-1 and Holm-Bonferoni for Stage-2. Variants indicated in green were identified as outliers in Stage-1 (column entitled:Type), and confirmed to be significant outliers in Stage-2, these three variants were removed to refine the burden.

#### 2.2.14 Table S14

| | Mega-analysis | | |
|---|---|---|---|
| | **gene** | **group** | **pvalue** |
| **Table 3a: primary analysis** | **SORL1** | LOF+REVEL≥50 | 2.0E-25 |
| | | - REVEL 50-100 | 1.0E-13 |
| | | - LOF | 8.8E-22 |
| | **TREM2** | LOF+REVEL≥25 | 1.4E-21 |
| | | LOF+REVEL≥25 [refined] | NA |
| | | - REVEL 25-100 | 4.4E-21 |
| | | - LOF | 2.4E-02 |
| | | - LOF [refined] | NA |
| | **ABCA7** | LOF+REVEL≥25 | 6.0E-13 |
| | | - REVEL 25-100 | 7.3E-11 |
| | | - LOF | 8.8E-04 |
| | **ATP8B4** | LOF+REVEL≥25 | 7.4E-07 |
| | | - REVEL 25-100 | 4.9E-07 |
| | | - LOF | 7.3E-01 |
| | **ABCA1** | LOF+REVEL≥75 | 5.1E-07 |
| | | LOF+REVEL≥75 [refined] | NA |
| | | - REVEL 75-100 | 9.7E-05 |
| | | - REVEL 75-100 [refined] | NA |
| | | - LOF | 5.5E-05 |
| | ADAM10 | LOF+REVEL≥50 | 5.1E-06 |
| **Table 3b: GWAS-targeted analysis** | RIN3 | LOF+REVEL≥50 | 1.0E-05 |
| | | - REVEL 50-100 | 1.5E-05 |
| | | - LOF | 2.7E-01 |
| | CLU | LOF+REVEL≥25 | 5.0E-04 |
| | | - REVEL 25-100 | 3.7E-02 |
| | | - LOF | 2.7E-03 |
| | ZCWPW1 | LOF | 7.8E-04 |
| | ACE | LOF+REVEL≥75 | 9.0E-04 |
| | | - REVEL 75-100 | 7.5E-04 |
| | | - LOF | 1.8E-01 |

As a sensitivity analysis we calculated significance on the mega-analysis dataset for the most significant association, for the missense and LOF variants separately.

#### 2.2.15 Tables SG1-10

Tables SG1-S10 will be made available for download as spreadsheets in the online supplemental data upon formal publication of this manuscript.

### 3 ACKNOWLEDGMENTS

#### 3.1 Study participants and personnel involved in sample collection

The authors are grateful to the study participants, their family members, and the participating general practitioners, pharmacists and all laboratory personnel involved in blood collection, DNA isolation, and DNA biobanking.

#### 3.2 SURF supercomputer facility

The work in this manuscript was carried out on the Cartesius supercomputer, which is embedded in the Dutch national e-infrastructure with the support of SURF Cooperative. Computing hours were granted in 2016, 2017, 2018 and 2019 to H. Holstege by the Dutch Research Council (project name: ‘100plus’; project numbers 15318 and 17232).

#### 3.3 Study Cohorts

##### 3.3.1 ADES-FR

This study was funded by grants from the Clinical Research Hospital Program from the French Ministry of Health (GMAJ, PHRC, 2008/067), the CNR-MAJ, the JPND PERADES, Equipe FRM DEQ20170336711, and Fondation Alzheimer (ECASCAD study). This research was supported by the Laboratory of Excellence GENMED (Medical Genomics) grant no. ANR-10-LABX-0013 managed by the National Research Agency (ANR) part of the Investment for the Future program. This work was also supported by Foundation Alzheimer, the Institut Pasteur de Lille, Inserm, the Haut-de-France and Lille Métropole Communauté Urbaine council, and the French government's LABEX (laboratory of excellence program investment for the future) DISTALZ grant (Development of Innovative Strategies for a Transdisciplinary approach to Alzheimer's disease). The 3C Study supports are listed on the Study Website (www.three-city-study.com).

##### 3.3.2 AgeCoDe-UKBonn

The AgeCoDe cohort was funded in part by the German Federal Ministry of Education and Research (BMBF) (grants KNDD 01GI0710, 01GI0711, 01GI0712, 01GI0713, 01GI0714, 01GI0715, 01GI0716, 01ET1006B). Sequencing of AgeCoDe sample was in part funded by the German Research Foundation (DFG) grant RA 1971/6-1 to Alfredo Ramirez.

##### 3.3.3 Barcelona- SPIN

Support for Jordi Clarimon provided by Maratón RTVE (Spain).Support for Oriol Dols provided by the Association for Frontotemporal Degeneration (Clinical Research Postdoctoral Fellowship, AFTD).

##### 3.3.4 AC-EMC

Exome sequencing was funded by Alzheimer Nederland.

##### 3.3.5 ERF

The ERF study as a part of EUROSPAN (European Special Populations Research Network) was supported by European Commission FP6 STRP grant number 018947 (LSHG-CT-2006-01947) and also received funding from the European Community's Seventh Framework Programme (FP7/2007-2013)/grant agreement HEALTH-F4- 2007-201413 by the European Commission under the programme "Quality of Life and Management of the Living Resources" of 5th Framework Programme (no. QLG2-CT-2002-01254). High-throughput analysis of the ERF data was supported by a joint grant from the Netherlands Organization for Scientific Research and the Russian Foundation for Basic Research (NWO-RFBR 047.017.043).

##### 3.3.6 Rotterdam Study

The generation and management of the exome sequencing data for the Rotterdam Study was executed by the Human Genotyping Facility of the Genetic Laboratory of the Department of Internal Medicine, Erasmus MC, the Netherlands. The Rotterdam Study is funded by Erasmus Medical Center and Erasmus University, Rotterdam, the Netherlands Organization for Health Research and Development (ZonMw), the Research Institute for Diseases in the Elderly (RIDE) (014-93-015; RIDE2), the Ministry of Education, Culture and Science, the Ministry for Health, Welfare and Sports, the European Commission (DG XII), and the municipality of Rotterdam. Genetic data sets are also supported by the Netherlands Organization of Scientific Research NWO Investments (175.010.2005.011, 911-03-012), the Genetic Laboratory of the Department of Internal Medicine, Erasmus MC, and the Netherlands Genomics Initiative (NGI)/Netherlands Organization for Scientific Research (NWO), the Netherlands Consortium for Healthy Aging (NCHA), project 050-060-810, and by a Complementation Project of the Biobanking and Biomolecular Research Infrastructure Netherlands (BBMRI-NL; www.bbmri.nl ; project number CP2010-41). We thank Mr. Pascal Arp, Ms. Mila Jhamai, Mr. and Marijn Verkerk, for their help in creating the RS-Exome Sequencing database.

##### 3.3.7 ADC-Amsterdam

We thank all study participants and all personnel involved in data collection for the contributing studies. Research of Alzheimer center Amsterdam is part of the neurodegeneration research program of Amsterdam Neuroscience. Alzheimer Center Amsterdam is supported by Stichting Alzheimer Nederland and Stichting VUmc fonds. The clinical database structure was developed with funding from Stichting Dioraphte. This work was supported by Stichting Alzheimer Nederland (WE.09-2014-06, WE.05-2010-06); Stichting Dioraphte; Internationale Stichting Alzheimer Onderzoek (#11519); JPND-PERADES (ZonMw 733051022): Centralized Facility for Sequence to Phenotype analyses (ZonMW 9111025); Netherlands Consortium for Healthy Aging (NCHA 050-060-810); Biobanking and Biomolecular Research Infrastructure Netherlands (BBMRI-NL CP2010-41); Netherlands Genomics Initiative (NGI)/NWO. This study is further supported by ABOARD, a public-private partnership receiving funding from ZonMW (#73305095007) and Health~Holland, Topsector Life Sciences & Health (PPP-allowance; #LSHM20106). This research is performed by using data from the Parelsnoer Institute an initiative of the Dutch Federation of University Medical Centres (www.parelsnoer.org).

##### 3.3.8 100-plus Study

Cohort collection and exome sequencing of the 100-plus Study cohort was supported by Stichting Alzheimer Nederland (WE.09-2014-03); HorstingStuit Foundation, VUmc Foundation, and the Dioraphte Foundation (Project 17020403), Memorabel (ZonMW project number #733050814, #733050512) and Stichting VUmcFonds. Additional support is from ABOARD, a public-private partnership receiving funding from ZonMW (#73305095007) and Health~Holland, Topsector Life Sciences & Health (PPP-allowance; #LSHM20106).

##### 3.3.9 EMIF-90+

—

##### 3.3.10 CBC: Control Brain Consortium

This work was supported by the UK Dementia Research Institute which receives its funding from DRI Ltd, funded by the UK Medical Research Council, Alzheimer's Society and Alzheimer's Research UK, Medical Research Council (award number MR/N026004/1). Wellcome Trust Hardy (award number 202903/Z/16/Z), Dolby Family Fund; National Institute for Health Research University College London Hospitals Biomedical Research Centre; BRCNIHR Biomedical Research Centre at University College London Hospitals NHS Foundation Trust and University College London.

J. Hardy was supported by the Dolby Foundation and the JPND PERADES. J.B. and R.G were supported by the National Institute on Aging of the National Institutes of Health under Award Number R01AG067426. The content is solely the responsibility of the authors and does not necessarily represent the official views of the National Institutes of Health.

##### 3.3.11 PERADES

We thank all individuals who participated in the study. We also want to express our gratitude to the MRC Centre Core Team for the laboratory support and the Advanced

Research Computing at Cardiff University (ARCCA) for the computational support. Cardiff University was supported by the Medical Research Council. Cardiff University was also supported by the European Joint Programme for Neurodegenerative Disease, Alzheimer's Research UK, the Welsh Assembly Government, and a donation from the Moondance Charitable Foundation. Cardiff University acknowledges the support of the UK Dementia Research Institute, which receives its funding from UK DRI Ltd, funded by the UK Medical Research Council, Alzheimer's Society and Alzheimer's Research UK. Cambridge University acknowledges support from the MRC. The University of Southampton acknowledges support from the Alzheimer’s Society. ARUK provided support to Nottingham University. Join Dementia Research (JDR) is funded by the Department of Health and delivered by the National Institute for Health Research in partnership with Alzheimer Scotland, Alzheimer's Research UK and Alzheimer's Society. IRCCS Santa Lucia Foundation acknowledges the Italian Ministry of Health for financial support (IMH_RC) of this study. The Centro de Biologia de Molecular Severo Ochoa (CSIS-UAM), CIBERNED, Instituto de Investigacion Sanitaria la Paz, University Hospital La Paz and the Universidad Autonoma de Madrid were supported by grants from the Ministerio de Educacion y Ciencia and the Ministerio de Sanidad y Consumo (Instituto de Salud Carlos III), and an institutional grant of the Fundacion Ramon Areces to the CMBSO. Thanks to I. Sastre and Dr A Martinez-Garcia for DNA preparation, and Drs P Gil and P Coria for their recruitment efforts. Department of Neurology, University Hospital Mutua de Terrassa, Terrassa, Barcelona, Spain was supported by CIBERNED, Centro de Investigacion Biomedica en Red de Enfermedades Neurodegenerativas, Instituto de Salud Carlos III, Madrid Spain and acknowledges Maria A Pastor (Department of Neurology, University of Navarra Medical School and Neuroimaging Laboratory, Center for Applied Medical Research, Pamplona, Spain), Manuel Seijo-Martinez (Department of Neurology, Hospital do Salnes, Pontevedra, Spain), Ramon Rene, Jordi Gascon and Jaume Campdelacreu (Department of Neurology, Hospital de Bellvitage, Barcelona, Spain) for providing DNA samples. Hospital de la Sant Pau, Universitat Autonoma de Spain acknowledges support from the Spanish Ministry of Economy and Competitiveness (grant number PI12/01311), and from Generalitat de Catalunya (2014SGR-235). The Santa Lucia Foundation and the

Fondazione Ca' Granda IRCCS Ospedale Policlinico, Italy, acknowledge the Italian Ministry of Health (grant RC 10.11.12.13/A)

##### 3.3.12 StEP-AD cohort

The Stanford Extreme Phenotypes in Alzheimer's Disease (StEP AD) Study is funded by the National Institutes of Health: R01AG060747 and AG047366

##### 3.3.13 Knight-ADRC

NIH P50 AG05681, P01 AG03991, P01 AG026276, NIA U01 AG058922;

##### 3.3.14 UCSF/NYGC/UAB

—

##### 3.3.15 UCL-DRC EOAD

This work was supported by the Medical Research Council (UK), the Biomedical Research Centre at University College London Hospitals NHS Foundation Trust and charitable donations to the UCL Dementia Research Centre.

##### 3.3.16 ADSP

The Alzheimer's Disease Sequencing Project (ADSP) is comprised of two Alzheimer's Disease (AD) genetics consortia and three National Human Genome Research Institute (NHGRI) funded Large Scale Sequencing and Analysis Centers (LSAC). The two AD genetics consortia are the Alzheimer's Disease Genetics Consortium (ADGC) funded by NIA (U01 AG032984), and the Cohorts for Heart and Aging Research in Genomic Epidemiology (CHARGE) funded by NIA (R01 AG033193), the National Heart, Lung, and Blood Institute (NHLBI), other National Institute of Health (NIH) institutes and other foreign governmental and non-governmental organizations. The Discovery Phase analysis of sequence data is supported through UF1AG047133 (to Drs. Schellenberg, Farrer, Pericak-Vance, Mayeux, and Haines); U01AG049505 to Dr. Seshadri; U01AG049506 to Dr. Boerwinkle; U01AG049507 to Dr. Wijsman; and U01AG049508 to Dr. Goate and the

Discovery Extension Phase analysis is supported through U01AG052411 to Dr. Goate, U01AG052410 to Dr. Pericak-Vance and U01 AG052409 to Drs. Seshadri and Fornage, U54 AG052427 to Drs. Schellenberg and Wang, and R01 AG054060 to Dr Naj. The ADGC cohorts include: Adult Changes in Thought (ACT) (UO1 AG006781, UO1 HG004610, UO1 HG006375, U01 HG008657), the Alzheimer's Disease Centers (ADC) ( P30 AG019610, P30 AG013846, P50 AG008702, P50 AG025688, P50 AG047266, P30 AG010133, P50 AG005146, P50 AG005134, P50 AG016574, P50 AG005138, P30 AG008051, P30 AG013854, P30 AG008017, P30 AG010161, P50 AG047366, P30 AG010129, P50 AG016573, P50 AG016570, P50 AG005131, P50 AG023501, P30 AG035982, P30 AG028383, P30 AG010124, P50 AG005133, P50 AG005142, P30 AG012300, P50 AG005136, P50 AG033514, P50 AG005681, and P50 AG047270), the Chicago Health and Aging Project (CHAP) (R01 AG11101, RC4 AG039085, K23 AG030944), Indianapolis Ibadan (R01 AG009956, P30 AG010133), the Memory and Aging Project (MAP) ( R01 AG17917), Mayo Clinic (MAYO) (R01 AG032990, U01 AG046139, R01 NS080820, RF1 AG051504, P50 AG016574), Mayo Parkinson's Disease controls (NS039764, NS071674, 5RC2HG005605), University of Miami (R01 AG027944, R01 AG028786, R01 AG019085, IIRG09133827, A2011048), the Multi-Institutional Research in Alzheimer's Genetic Epidemiology Study (MIRAGE) (R01 AG09029, R01 AG025259), the National Cell Repository for Alzheimer's Disease (NCRAD) (U24 AG21886), the National Institute on Aging Late Onset Alzheimer's Disease Family Study (NIA- LOAD) (R01 AG041797), the Religious Orders Study (ROS) (P30 AG10161, R01 AG15819), the Texas Alzheimer's Research and Care Consortium (TARCC) (funded by the Darrell K Royal Texas Alzheimer's Initiative), Vanderbilt University/Case Western Reserve University (VAN/CWRU) (R01 AG019757, R01 AG021547, R01 AG027944, R01 AG028786, P01 NS026630, and Alzheimer's Association), the Washington Heights-Inwood Columbia Aging Project (WHICAP) (RF1 AG054023), the University of Washington Families (VA Research Merit Grant, NIA: P50AG005136, R01AG041797, NINDS: R01NS069719), the Columbia University HispanicEstudio Familiar de Influencia Genetica de Alzheimer (EFIGA) (RF1 AG015473), the University of Toronto (UT) (funded by Wellcome Trust, Medical Research Council, Canadian Institutes of Health Research), and Genetic Differences (GD) (R01 AG007584). The CHARGE cohorts are supported in part by

National Heart, Lung, and Blood Institute (NHLBI) infrastructure grant HL105756 (Psaty), RC2HL102419 (Boerwinkle) and the neurology working group is supported by the National Institute on Aging (NIA) R01 grant AG033193.

This work was also supported by National Institute on Aging grants R01 AG048927 to Dr. Farrer, RF1 AG054080 to Dr. Beecham, U24 AG056270 to Dr. Mayeux, RF1 AG057519 to Dr. Farrer, U01 AG062602 to Dr. Farrer, R01 AG067501 to Dr. Mayeux, and U19 AG068753 to Dr. Farrer.

The CHARGE cohorts participating in the ADSP include the following: Austrian Stroke Prevention Study (ASPS), ASPS-Family study, and the Prospective Dementia Registry-Austria (ASPS/PRODEM-Aus), the Atherosclerosis Risk in Communities (ARIC) Study, the Cardiovascular Health Study (CHS), the Erasmus Rucphen Family Study (ERF), the Framingham Heart Study (FHS), and the Rotterdam Study (RS). ASPS is funded by the Austrian Science Fond (FWF) grant number P20545-P05 and P13180 and the Medical University of Graz. The ASPS-Fam is funded by the Austrian Science Fund (FWF) project I904),the EU Joint Programme - Neurodegenerative Disease Research (JPND) in frame of the BRIDGET project (Austria, Ministry of Science) and the Medical University of Graz and the Steiermärkische Krankenanstalten Gesellschaft. PRODEM-Austria is supported by the Austrian Research Promotion agency (FFG) (Project No. 827462) and by the Austrian National Bank (Anniversary Fund, project 15435. ARIC research is carried out as a collaborative study supported by NHLBI contracts (HHSN268201100005C, HHSN268201100006C, HHSN268201100007C, HHSN268201100008C, HHSN268201100009C, HHSN268201100010C, HHSN268201100011C, and HHSN268201100012C). Neurocognitive data in ARIC is collected by U01 2U01HL096812, 2U01HL096814, 2U01HL096899, 2U01HL096902, 2U01HL096917 from the NIH (NHLBI, NINDS, NIA and NIDCD), and with previous brain MRI examinations funded by R01-HL70825 from the NHLBI. CHS research was supported by contracts HHSN268201200036C, HHSN268200800007C, N01HC55222, N01HC85079, N01HC85080, N01HC85081, N01HC85082, N01HC85083, N01HC85086, and grants U01HL080295 and U01HL130114 from the NHLBI with additional contribution from the National Institute of Neurological Disorders and Stroke (NINDS). Additional support was provided by R01AG023629, R01AG15928, and R01AG20098 from the NIA. FHS research is supported by NHLBI contracts N01-HC-25195 and HHSN268201500001I. This study was also supported by additional grants from the NIA (R01s AG054076, AG049607 and AG033040 and NINDS (R01 NS017950). The ERF study as a part of EUROSPAN (European Special Populations Research Network) was supported by European Commission FP6 STRP grant number 018947 (LSHG-CT-2006-01947) and also received funding from the European Community's Seventh Framework Programme (FP7/2007-2013)/grant agreement HEALTH-F4- 2007-201413 by the European Commission under the programme "Quality of Life and Management of the Living Resources" of 5th Framework Programme (no. QLG2-CT-2002- 01254). High-throughput analysis of the ERF data was supported by a joint grant from the Netherlands Organization for Scientific Research and the Russian Foundation for Basic Research (NWO-RFBR 047.017.043). The Rotterdam Study is funded by Erasmus Medical Center and Erasmus University, Rotterdam, the Netherlands Organization for Health Research and Development (ZonMw), the Research Institute for Diseases in the Elderly (RIDE), the Ministry of Education, Culture and Science, the Ministry for Health, Welfare and Sports, the European Commission (DG XII), and the municipality of Rotterdam. Genetic data sets are also supported by the Netherlands Organization of Scientific Research NWO Investments (175.010.2005.011, 911-03-012), the Genetic Laboratory of the Department of Internal Medicine, Erasmus MC, the Research Institute for Diseases in the Elderly (014-93-015; RIDE2), and the Netherlands Genomics Initiative (NGI)/Netherlands Organization for Scientific Research (NWO) Netherlands Consortium for Healthy Aging (NCHA), project 050-060-810. All studies are grateful to their participants, faculty and staff. The content of these manuscripts is solely the responsibility of the authors and does not necessarily represent the official views of the National Institutes of Health or the U.S. Department of Health and Human Services.

The four LSACs are: the Human Genome Sequencing Center at the Baylor College of Medicine (U54 HG003273), the Broad Institute Genome Center (U54HG003067), The American Genome Center at the Uniformed Services University of the Health Sciences (U01AG057659), and the Washington University Genome Institute (U54HG003079).

Biological samples and associated phenotypic data used in primary data analyses were stored at Study Investigators institutions, and at the National Cell Repository for Alzheimer's Disease (NCRAD, U24AG021886) at Indiana University funded by NIA. Associated Phenotypic Data used in primary and secondary data analyses were provided by Study Investigators, the NIA funded Alzheimer's Disease Centers (ADCs), and the National Alzheimer's Coordinating Center (NACC, U01AG016976) and the National Institute on Aging Genetics of Alzheimer's Disease Data Storage Site (NIAGADS, U24AG041689) at the University of Pennsylvania, funded by NIA This research was supported in part by the Intramural Research Program of the National Institutes of health, National Library of Medicine. Contributors to the Genetic Analysis Data included Study Investigators on projects that were individually funded by NIA, and other NIH institutes, and by private U.S. organizations, or foreign governmental or nongovernmental organizations.

Data collection and sharing for this project was funded by the Alzheimer's Disease Neuroimaging Initiative (ADNI) (National Institutes of Health Grant U01 AG024904) and DOD ADNI (Department of Defense award number W81XWH-12-2-0012). ADNI is funded by the National Institute on Aging, the National Institute of Biomedical Imaging and Bioengineering, and through generous contributions from the following: AbbVie, Alzheimer's Association; Alzheimer's Drug Discovery Foundation; Araclon Biotech; BioClinica, Inc.; Biogen; Bristol-Myers Squibb Company; CereSpir, Inc.; Cogstate; Eisai Inc.; Elan Pharmaceuticals, Inc.; Eli Lilly and Company; EuroImmun; F. Hoffmann-La Roche Ltd and its affiliated company Genentech, Inc.; Fujirebio; GE Healthcare; IXICO Ltd.; Janssen Alzheimer Immunotherapy Research & Development, LLC.; Johnson & Johnson Pharmaceutical Research & Development LLC.; Lumosity; Lundbeck; Merck & Co., Inc.; Meso Scale Diagnostics, LLC.; NeuroRx Research; Neurotrack Technologies; Novartis Pharmaceuticals Corporation; Pfizer Inc.; Piramal Imaging; Servier; Takeda Pharmaceutical Company; and Transition Therapeutics. The Canadian Institutes of Health Research is providing funds to support ADNI clinical sites in Canada. Private sector contributions are facilitated by the Foundation for the National Institutes of Health (www.fnih.org). The grantee organization is the Northern California Institute for Research and Education, and the study is coordinated by the Alzheimer's Therapeutic Research

Institute at the University of Southern California. ADNI data are disseminated by the Laboratory for Neuro Imaging at the University of Southern California.

### 4 Supplemental Authors

##### 4.1.1 PERADES Cohort:

Keeley Brookes[1], Tamar Guetta-Baranes[2], Elisa Toppi[3], Francesca Salani[3], Marina Arcaro[4], Chiara Fenoglio[5], Roberta Cecchetti[6], Elio Scarpini[7], Sandro Sorbi[8], Monica Diez-Fairen[9], Ignacio Alvarez[9], Miquel Aguilar[9], MRC: Simon Lovestone[10], John Powell[11], Carol Brayne[12], David Rubinsztein[12], Nandini Badarinarayan[13], Eloy Rodriguez-Rodriguez[14], Carmen Lage[14], Sara Lopez-Garcia[14], Emanuele Costantini[15], Michela Orsini[15], Francesco Panza[16], Nerisa Banaj[17], Federica Piras[17] and Daniela Vecchio[17]

**(1)**Nottingham Trent; **(2 )**Human Genetics. UoN; (**3**) IRCCS Fondazione Santa Lucia, Department of Clinical and Behavioral Neurology, Experimental Neuro-psychobiology Lab Via Ardeatina, 306, I-00179 Roma, Italy; **(4)** Fondazione IRCCS Ca' Granda, Ospedale Policlinico; **(5)** University of Milan, Dino Ferrari Center, Milan, Italy; 6Institute of Gerontology and Geriatrics, Department of Medicine and Surgery, University of Perugia, Italy; **(7)** University of Milan, Centro Dino Ferrari, CRC Molecular basis of Neuro-Psycho-Geriatrics diseases, Milan, Italy; **(8)** Department of Neuroscience, Psychology, Drug Research and Child Health , University of Florence, Italy; **(9)** Memory Disorders Unit, Department of Neurology, Hospital Universitari Mutua de Terrassa, Terrassa, Barcelona, Spain; **(10)** Department of Psychiatry, Medical Sciences Division, University of Oxford, Oxford, UK.; **(11)** Kings College London, Institute of Psychiatry, Department of Neuroscience, De Crespigny Park, Denmark Hill, London, UK; **(12)** Institute of Public Health, University of Cambridge, Cambridge, UK.; (**13)** Division of Psychological Medicine and Clinical Neuroscience, School of Medicine, Cardiff University, Cardiff, UK; **(14)** Neurology Service and Centro de Investigación en Red de Enfermedades Neurodegenerativas (CIBERNED), Marques de Valdecilla University Hospital (University of Cantabria and IDIVAL), Santander, Spain **(15)** Department of Neuroscience, Catholic University of Sacred Heart, Fondazione Policlinico Universitario A. Gemelli IRCCS, Rome, Italy **(16)** National Institute of Gastroenterology and Research Hospital IRCCS "S. De Bellis" Castellana Grotte, Bari Italy **(17)** Laboratory of Neuropsychiatry,IRCCS Santa Lucia Foundation, Rome, Italy

#### 4.1.2 StEP AD investigators

Clifton L. Dalgard, PhD[1], William J. Jagust, MD[2], Sterling C. Johnson, PhD[3], David A. Wolk, MD[4], Joel H. Kramer, PsyD[5], Bradford C. Dickerson, MD[6], David A. Bennett, MD[7], Sofiya Milman, MD[8], Bruno Dubois, MD, PhD[9], Ruth O'Hara, PhD[10], Sherry A. Beaudreau, PhD[11]

[1]Uniformed Services University of the Health Sciences;[2]University of California, Berkeley; [3]University of Wisconsin; [4]University of Pennsylvania; [5]University of California, San Francisco; [6]Harvard University; [7]Rush University; [8]Albert Einstein College of Medicine; [9]Brain and Spine Institute (ICM), France; [10]Stanford University
[11]VA Palo Alto Health Care System

#### 4.1.3 Knight ADRC investigators

Achal Neupane[1,3,4], John P Budde[1,3,4], Fengxian Wang[1,3,4], Joanne Norton[1,3,4], Gen Gentsch[1,3,4], John C Morris[2,3]

Departments of [1]Psychiatry, [2]Neurology, [3]Hope Center for Neurological Disorders, [4]NeuroGenomics and Informatics Center, Washington University School of Medicine, St. Louis, Missouri, USA.

#### 4.1.4 ADNI database

Data used in preparation of this article were obtained from the Alzheimer's Disease Neuroimaging Initiative (ADNI) database (adni.loni.usc.edu). As such, the investigators within the ADNI contributed to the design and implementation of ADNI and/or provided data but did not participate in analysis or writing of this report. A complete listing of ADNI investigators can be found at:
http://adni.loni.usc.edu/wp-content/uploads/how_to_apply/ADNI_Acknowledgement_List.pdf
